# Air temperature and thrombotic cardiovascular disease in England: case time series studies using whole-population electronic health records during 2020-2022

**DOI:** 10.64898/2026.09.02.26358673

**Authors:** Isabel Johanna Walter, Alexia Sampri, Thomas Bolton, Stelios Boulitsakis Logothetis, Shivang Pandey, Leonardo Olivetti, Manuel Martellini O Nocentini, Nathalie Conrad, Yueying Li, Fionna Chalmers, James Farrell, Rachel Denholm, Samantha Ip, Luigi Filippo Brizzi, Gabriele Messori, Arturo De La Cruz Libardi, Antonio Gasparrini, Angela Wood, Elena Raffetti, the CVD-COVID-UK/COVID-IMPACT Consortium

**Author notes:** Corresponding author Isabel Johanna Walter, MD Department of Global Public Health Karolinska Institutet, SE-171 77 Stockholm, Sweden. Conflicts of interest: None declared.

## Abstract

**Background:** Hot and cold air temperatures have been associated with fatal thrombotic cardiovascular disease, but evidence for non-fatal outcomes and drivers of susceptibility remains limited. Given the lasting cardiovascular effects of COVID-19 and its potential interaction with temperature exposure, we examined the impact of air temperature on thrombotic cardiovascular disease, as well as effect modification by COVID-19 and person-level characteristics.

**Methods:** We conducted case time series studies using whole-population electronic health records in England (Jan 2020-Dec 2022), linked with small-area daily mean air temperature. We estimated cumulative short-term effects of temperature on arterial thrombotic events (AEs), venous thrombotic events (VEs), acute myocardial infarction (AMI), and ischaemic stroke (IS), using conditional Poisson regression with distributed lag non-linear models, including interactions with COVID-19 status and person-level characteristics.

**Results:** Among 49,080,575 individuals, we identified 604,655 AEs, 215,090 VEs, 275,390 AMIs, and 231,485 ISs. Hot temperatures (99th percentile 23.1°C vs median 10.8°C) were associated with higher VE risk over 21 days (incidence rate ratio [IRR] 1.33, 95% CI 1.21-1.47), particularly among older adults, those with pre-existing conditions, and individuals of Black ethnic group. Cold temperatures (1st percentile -0.7°C vs median) were associated with AMI over 14 days (IRR 1.13, 95% CI 1.05-1.21), especially among those aged ≥65 years. No associations were found for AEs or IS. Cold temperatures increased VE risk among individuals with COVID-19 in the prior 6 months (IRR 1.40, 95% CI 1.05- 1.86), but not others.

**Conclusions:** Susceptibility to temperature-related thrombotic events varies across population groups and may change following recent infectious disease, such as COVID-19.

## Introduction

Hot and cold air temperatures are key drivers of communicable and non-communicable diseases^1–4^. Global warming has surpassed 1.5 °C above pre-industrial levels^5^, and Europe is warming at twice the global average rate^6^. As temperatures continue to rise, so do the frequency and intensity of extreme weather^7^. In England, temperature already exerts a measurable health burden: between 2000 and 2019, standardised excess mortality rates were 1.57 (95% CI 1.21-1.90) per 100,000 person-years associated with hot temperatures and 122.34 (112.90-131.52) due to cold temperatures^8^.

Both hot and cold air temperatures are independently associated with elevated cardiovascular mortality, plausibly through thermoregulatory and inflammatory pathways^9–13^. In Europe, cold temperatures, and less consistently hot temperatures, have been associated with an increased risk of myocardial infarction^9,13,14^, whereas studies on air temperature and ischaemic stroke or venous thrombotic events are conflicting or limited^10,15,16^. In addition, the timing of temperature-related thrombotic effects and whether associations differ between arterial and venous events remain incompletely characterised.

Infectious pathogens, such as SARS-CoV-2, present additional challenges when examining the impact of temperatures on cardiovascular health. Acute infections are well-established triggers of arterial and venous thrombotic events through systemic inflammation, endothelial dysfunction, and activation of coagulation pathways^17,18^. Moreover, the seasonal patterning of many infections, particularly in colder periods, may confound or mediate observed associations between temperature and cardiovascular outcomes, complicating causal interpretation^19^. However, previous temperature- health studies have rarely accounted for infection dynamics or examined whether susceptibility to temperature-related thrombotic events varies according to recent infection status, including COVID-19.

More broadly, studies examining air temperature exposure in relation to thrombotic cardiovascular outcomes beyond mortality remain scarce^9,13^. Additionally, most evidence relies on spatially aggregated data, limiting the ability to evaluate risks based on individual factors^12,14–16^. Assessing the health risk associated with moderate and extreme temperatures and identifying who is most vulnerable is essential to inform targeted climate adaptation measures.

To address these gaps, we aimed to characterise the association between daily mean air temperature and thrombotic cardiovascular disease in England. For the first time, we leverage novel methodologies to link environmental exposure data with whole-population electronic health records at the individual level, enabling the assessment of effect modification by prior COVID-19 diagnosis and person-level characteristics influencing susceptibility.

## Methods

### Study design and population

We conducted population-based case time series studies within a retrospective cohort of adults in England, using linked electronic health record datasets with near-complete population coverage. This study made use of anonymised data held in NHS England’s Secure Data Environment service for England and made available through the British Heart Foundation Data Science Centre’s <u>CVD-</u> <u>COVID-UK/COVID-IMPACT Consortium</u>. We followed a pre-specified protocol and proposal (Supplement). Data included primary care (the General Practice Extraction Service Data for Pandemic Planning and Research [GDPPR]), secondary care (Hospital Episode Statistics [HES]), and the Office for National Statistics Civil Registration of Deaths (Table S1). This study is reported in accordance with the STROBE guidelines (Table S2).

Health data was linked to daily mean air temperature using each individual’s Lower Super Output Area (LSOA) on the corresponding date (Supp. methods). The LSOA was based on the residential address, and identified from GDPPR, COVID-19 vaccinations, or HES data; individual-level addresses were not accessible to researchers. England has 32,844 LSOAs (2011 census), which are census-based aggregation units, each covering 1,000-3,000 residents.

The study period was Jan 1, 2020, to Dec 31, 2022. The cohort comprised individuals aged ≥18 years who were alive and registered in primary care (GDPPR) on Jan 1, 2020, with a recorded LSOA within the preceding two years before the study start date or during the study period. Individuals with an LSOA outside England during follow-up were excluded.

The North East-Newcastle and North Tyneside 2 research ethics committee provided ethical approval for the CVD-COVID-UK/COVID-IMPACT research programme (REC No 20/NE/0161) to access, within secure trusted research environments, unconsented, whole-population, de-identified data from electronic health records collected as part of patients’ routine healthcare.

### Environmental exposure

Daily mean air temperature data was obtained from a previous published study^8^, providing 1 × 1 km resolution daily mean surface air temperature across England from the HadUK-Grid database, developed by the UK Meteorological Office^20^. Particulate matter (PM2.5) concentrations were derived from a 1 x 1 km resolution dataset that integrated ground monitoring observations, satellite-derived data, climate reanalysis, chemical transport model datasets, traffic and land-use data^21^, covering the study period until Dec 31, 2021. LSOA-level temperature and PM2.5 series were derived by computing the area-weighted average of the values of all the grid cells intersecting the LSOA boundaries, with weights proportional to the intersection areas^8^.

### Baseline characteristics

Sociodemographic variables were defined at baseline (Jan 1, 2020) for all individuals in the cohort and included age, sex, ethnic group (Asian or Asian British; Black, Black British, Caribbean or African; mixed or multiple ethnic groups; other ethnic group; missing), region of residence, urban-rural classification, and area-based socioeconomic deprivation. Deprivation was measured using the 2019 English Index of Multiple Deprivation (IMD), linked from each individual’s residential LSOA, and grouped as more deprived (quintiles 1-3) versus less deprived (quintiles 4-5). Pre-existing conditions were identified from primary (GDPPR) and secondary (HES) care records using SNOMED CT and ICD-10 codes, based on established CVD-COVID-UK/COVID-IMPACT phenotypes^22^ or codelists manually curated by two clinicians (IJW; LFB). These included hypertension, diabetes, chronic kidney disease, depression, heart disease, stroke, excess weight, smoking status, and a composite of major pre-existing conditions (diabetes, chronic obstructive pulmonary disease [COPD], asthma, liver disease, chronic kidney disease, or cancer). Look-back periods and full phenotype definitions, including codelists, are provided in the Supplementary methods and Table S3.

### Outcomes

The primary study outcomes were defined as venous and arterial thrombotic events, capturing distinct thrombotic processes with different pathophysiological mechanisms and clinical presentations. Venous thrombotic events included deep vein and superficial venous thrombosis, pulmonary embolism, and intracranial venous thrombosis. Arterial thrombotic event was a composite of acute myocardial infarction, stroke (unspecified or ischaemic), other arterial embolism, arterial dissection or ruptured aneurysm, and retinal infarction. Acute myocardial infarction and ischaemic stroke were further analysed separately as they are the most common and clinically significant arterial outcomes. Outcomes were defined by the ICD-10 code in the primary hospital admission position or primary cause of death, based on codelists curated by two clinicians (IJW; LFB) (Table S4). For each outcome, all events during the study period were identified, allowing multiple events per individual provided they were at least 30 days apart.

### Statistical analysis

We estimated cumulative incidence rate ratios (IRRs) and 95% confidence intervals (CIs) to assess the association between daily mean air temperature and thrombotic cardiovascular events using a case time series design^23^, implemented with conditional Poisson regression. This design supports causal interpretation of the IRRs by comparing different time periods within the same individual, thereby controlling by design for all time-invariant personal characteristics^23^. The resulting IRRs represent the relative change in the rate of thrombotic cardiovascular events in the days following exposure to a given daily temperature, compared with the median temperature across all LSOAs in England during the study period.

A case time series dataset was constructed by appending the series for all individuals with at least one thrombotic cardiovascular event, with one record per day of follow-up from baseline to death or study end per individual. Each record included daily mean air temperature and an indicator of outcome occurrence. Four datasets were constructed, corresponding to one case time series study per outcome (Table S5). Temperature-outcome associations were fitted using distributed lag non-linear models (DLNMs) with the median daily mean air temperature in England as the reference value. Delayed effects were modelled over lags 0-21 days, allowing air temperature exposure to influence risk on the same day and up to 21 subsequent days, consistent with established literature^8^. A cross-basis matrix of daily mean air temperature, grouped by individual, was defined using natural splines for the temperature dimension (knots: 10th, 75th, 90th percentiles; boundaries 1st and 99th) and for the lag dimension over 0-21 days, with three internal knots placed at equally spaced values on the log scale^8^. Alternative spline specifications were assessed for model fit as well (Table S6). Models included the cross-basis of daily mean air temperature as exposure and day of week as covariate, with strata defined by individual, calendar year, and calendar month to control for within-person seasonality and long-term temporal trends^8,23^ (Figure S1). Due to larger counts of arterial thrombotic events, analyses were run in two random halves of the dataset for computational feasibility; coefficients were pooled using fixed-effect meta-analysis^24^.

Subgroup analyses were performed by COVID-19 diagnosis within 6 months prior to the outcome event and by baseline characteristics. Effect modification was assessed by including an interaction term between the cross-basis matrix and the covariate. Significance of the interaction was assessed through the Likelihood Ratio Test.

We quantified the absolute burden attributable to temperature as annual excess event rates per 100,000 person-years, defining heat and cold as daily mean temperatures above and below the median, respectively^25^.

To assess the robustness of findings to various modelling assumptions we performed a number of sensitivity analyses (Rationale in Table S7): (i) restricting follow-up to start from 1 May 2020 to account for reduced healthcare use in March-April 2020 (Figure S2); (ii) use of alternative lag windows of 0-14, 0-7, and 0-4 days; (iii) excluding of fatal events defined as death within 7, 14, or 30 days following an outcome event; (iv) restricting to fatal events; (v) censoring at the exact date of death; and (vi) restricting to the first event per individual. Further analyses adjusted for COVID-19 diagnosis, national lockdown periods, and PM2.5 as time-varying covariates, to assess the robustness of the temperature-outcome associations to co-exposures and other time-varying contextual factors.

When adjusting for PM2.5, follow-up was limited to 1 January 2020-31 December 2021 due to data availability. Full phenotype definitions of COVID-19 diagnosis and all time-varying covariates are provided in the Supplementary methods.

All statistical analyses were performed using Spark SQL (version 3.3.0), Python (version 3.9.21), and R (version 4.3.1). All code and codelists are publicly available on <u>GitHub</u> and the <u>HDRUK Phenotype</u> <u>library</u>(PH4045-PH4061).

## Results

The cohort included 49,080,575 individuals (Figure S3), with 50.3% women, 79.1% aged 18-64 years, 78.9% of White ethnic group, 83.8% residing in urban areas, and 20.1% living in the most deprived quintile (Table 1). The highest proportions lived in London (17.8%) and the South East (16.0%). At baseline, 43.4% had never smoked, 27.1% had excess weight, and common pre-existing conditions included hypertension (15.7%), depression (9.8%), and heart disease (9.4%) (Table 1). During follow-up, 2,171,910 individuals (4.5%) died, and 28.1% had a recorded COVID-19 diagnosis (Table 1).

**Table 1:** Baseline characteristics of the overall study population and individuals included in each case time series analysis. Data are presented as N (%) unless otherwise indicated. Age is presented as mean (standard deviation, SD). The “Total” column includes all individuals eligible for the study cohort. Outcome-specific columns include all individuals contributing to the corresponding case time series analysis, defined as those who experienced at least one event of the respective outcome during follow-up. Sample sizes are given in the column headings.

|  | **Total <sup>a</sup>**<br>**(n = 49,080,575)**<br>*N (%)* | **Venous thrombotic event <sup>a</sup>**<br>**(n = 202,790)**<br>*N (%)* | **Arterial thrombotic event <sup>a</sup>**<br>**(n = 546,185)**<br>*N (%)* | **Acute myocardial infarction <sup>a</sup>**<br>**(n = 260,915)**<br>*N (%)* | **Ischaemic stroke <sup>a</sup>**<br>**(n=208,965)**<br>*N (%)* |
| --- | --- | --- | --- | --- | --- |
| **Demographics** |  |  |  |  |  |
| **Sex** |  |  |  |  |  |
| Female | 24,687,730 (50.3%) | 104,715 (51.6%) | 216,485 (39.6%) | 87,265 (33.4%) | 98,555 (47.2%) |
| Male | 24,392,845 (49.7%) | 98,075 (48.4%) | 329,700 (60.4%) | 173,650 (66.6%) | 110,410 (52.8%) |
| **Age at study start**<br>mean (SD) | 47.9 (18.8) | 63.2 (17.5) | 70.9 (14.1) | 68.6 (13.8) | 73.0 (13.8) |
| **Age group** |  |  |  |  |  |
| 18-64 years | 38,815,780 (79.1%) | 98,600 (48.6%) | 177,830 (32.6%) | 103,800 (39.8%) | 54,890 (26.3%) |
| 65-74 years | 5,537,920 (11.3%) | 45,675 (22.5%) | 130,305 (23.9%) | 63,630 (24.4%) | 48,425 (23.2%) |
| 75-84 years | 3,358,645 (6.8%) | 40,115 (19.8%) | 146,720 (26.9%) | 60,540 (23.2%) | 63,670 (30.5%) |
| 85 years or older | 1,368,235 (2.8%) | 18,400 (9.1%) | 91,335 (16.7%) | 32,950 (12.6%) | 41,980 (20.1%) |
| **Ethnic group** |  |  |  |  |  |
| White | 38,721,535 (78.9%) | 185,875 (91.7%) | 488,915 (89.5%) | 229,360 (87.9%) | 188,550 (90.2%) |
| Asian or Asian British | 5,099,510 (10.4%) | 6,305 (3.1%) | 34,530 (6.3%) | 21,710 (8.3%) | 10,665 (5.1%) |
| Black, Black British, Caribbean or African | 1,922,355 (3.9%) | 6,090 (3%) | 11,340 (2.1%) | 4,035 (1.5%) | 5,745 (2.7%) |
| Mixed or multiple ethnic groups | 820,875 (1.7%) | 1,970 (1%) | 3,775 (0.7%) | 1,760 (0.7%) | 1,480 (0.7%) |
| Other ethnic group | 1,161,040 (2.4%) | 1,720 (0.8%) | 5,135 (0.9%) | 2,715 (1%) | 1,795 (0.9%) |
| Missing | 1,355,260 (2.8%) | 830 (0.4%) | 2,495 (0.5%) | 1,335 (0.5%) | 730 (0.3%) |
| **Deprivation index quintiles** |  |  |  |  |  |
| 1 (most deprived) | 9,852,765 (20.1%) | 43,465 (21.4%) | 116,760 (21.4%) | 58,450 (22.4%) | 42,980 (20.6%) |
| 2 | 10,445,475 (21.3%) | 40,755 (20.1%) | 110,400 (20.2%) | 53,275 (20.4%) | 41,895 (20.0%) |
| 3 | 10,018,145 (20.4%) | 40,345 (19.9%) | 111,455 (20.4%) | 52,820 (20.2%) | 42,770 (20.5%) |
| 4 | 9,581,705 (19.5%) | 40,190 (19.8%) | 107,560 (19.7%) | 50,165 (19.2%) | 42,035 (20.1%) |
| 5 (least deprived) | 9,182,480 (18.7%) | 38,040 (18.8%) | 100,015 (18.3%) | 46,205 (17.7%) | 39,285 (18.8%) |
| **Geographical location** |  |  |  |  |  |
| **Region** |  |  |  |  |  |
| London | 8,751,175 (17.8%) | 2,780 (10.7%) | 59,500 (10.9%) | 27,635 (10.6%) | 22,925 (11%) |
| West midlands | 5,017,760 (10.2%) | 23,495 (11.6%) | 59,540 (10.9%) | 29,335 (11.2%) | 22,295 (10.7%) |
| East midlands | 4,140,795 (8.4%) | 18,275 (9.0%) | 48,850 (8.9%) | 23,650 (9.1%) | 19,250 (9.2%) |
| North west | 6,271,990 (12.8%) | 29,835 (14.7%) | 80,750 (14.8%) | 39,850 (15.3%) | 29,765 (14.2%) |
| North east | 2,276,150 (4.6%) | 12,355 (6.1%) | 32,115 (5.9%) | 15,125 (5.8%) | 13,130 (6.3%) |
| South west | 4,769,400 (9.7%) | 20,640 (10.2%) | 62,185 (11.4%) | 29,390 (11.3%) | 23,395 (11.2%) |
| South east | 7,868,425 (16%) | 34,900 (17.2%) | 84,635 (15.5%) | 38,735 (14.8%) | 33,095 (15.8%) |
| East of England | 5,297,230 (10.8%) | 20,775 (10.2%) | 58,380 (10.7%) | 27,505 (10.5%) | 22,130 (10.6%) |
| Yorkshire and the Humber | 4,687,645 (9.6%) | 20,735 (10.2%) | 60,235 (11%) | 29,690 (11.4%) | 22,975 (11%) |
| **Rural urban class** |  |  |  |  |  |
| Urban | 41,118,955 (83.8%) | 165,285 (81.5%) | 439,035 (80.4%) | 210,915 (80.8%) | 167,290 (80.1%) |
| Rural | 7,961,620 (16.2%) | 37,505 (18.5%) | 107,150 (19.6%) | 50,000 (19.2%) | 41,675 (19.9%) |
| **Lifestyle factors** |  |  |  |  |  |
| **Smoking status** |  |  |  |  |  |
| Never smoked | 21,298,225 (43.4%) | 74,250 (36.6%) | 162,350 (29.7%) | 74,165 (28.4%) | 69,505 (33.3%) |
| Ex-smoker | 15,287,815 (31.1%) | 91,345 (45.0%) | 261,510 (47.9%) | 123,155 (47.2%) | 99,630 (47.7%) |
| Current smoker | 7,954,440 (16.2%) | 32,760 (16.2%) | 112,220 (20.5%) | 58,880 (22.6%) | 35,765 (17.1%) |
| Missing | 4,540,095 (9.3%) | 4,435 (2.2%) | 10,110 (1.9%) | 4,715 (1.8%) | 4,070 (1.9%) |
| Excess weight (BMI ≥ 25) | 13,309,335 (27.1%) | 91,580 (45.2%) | 243,100 (44.5%) | 121,485 (46.6%) | 91,945 (44.0%) |
| **Medical history** |  |  |  |  |  |
| Hypertension | 7,713,155 (15.7%) | 72,430 (35.7%) | 287,230 (52.6%) | 129,700 (49.7%) | 114,485 (54.8%) |
| Diabetes | 3,693,195 (7.5%) | 30,825 (15.2%) | 147,995 (27.1%) | 75,000 (28.7%) | 57,020 (27.3%) |
| Chronic kidney disease | 2,502,305 (5.1%) | 31,030 (15.3%) | 128,345 (23.5%) | 56,675 (21.7%) | 51,790 (24.8%) |
| Depression | 4,815,140 (9.8%) | 31,110 (15.3%) | 63,935 (11.7%) | 30,330 (11.6%) | 23,900 (11.4%) |
| Heart disease | 4,623,365 (9.4%) | 45,330 (22.4%) | 227,945 (41.7%) | 107,505 (41.2%) | 85,410 (40.9%) |
| Pre-existing conditions<sup>b</sup> | 10,161,715 (20.7%) | 88,745 (43.8%) | 283,895 (52.0%) | 133,810 (51.3%) | 138,975 (66.5%) |
| **COVID-19 status** |  |  |  |  |  |
| COVID-19 diagnosis during follow-up | 13,771,005 (28.1%) | 22,675 (11.2%) | 49,225 (9.01%) | 20,200 (7.7%) | 20,915 (10.0%) |

a. Numbers are rounded to fives to comply with privacy regulations. b. Composite of COPD, diabetes, cancer, liver disease, chronic kidney disease, asthma

A total of 604,655 arterial thrombotic events, 215,090 venous thrombotic events, 275,390 acute myocardial infarctions, and 231,485 ischaemic strokes were recorded during follow-up (Table S8). These corresponded to 546,185, 202,790, 260,915, and 208,965 individuals, respectively, as some participants experienced multiple events (Table 1, Table S9). Daily mean air temperature in England ranged from -6.5 °C to 31.4 °C over the study period. The 1st, 5th, 25th, 50th, 75th, 95th and 99th percentiles were -0.7 °C, 2.8 °C, 7.0 °C, 10.8 °C, 15.4 °C, 19.6 °C , and 23.1 °C (Figure 1, Table S10).

**Figure 1:**
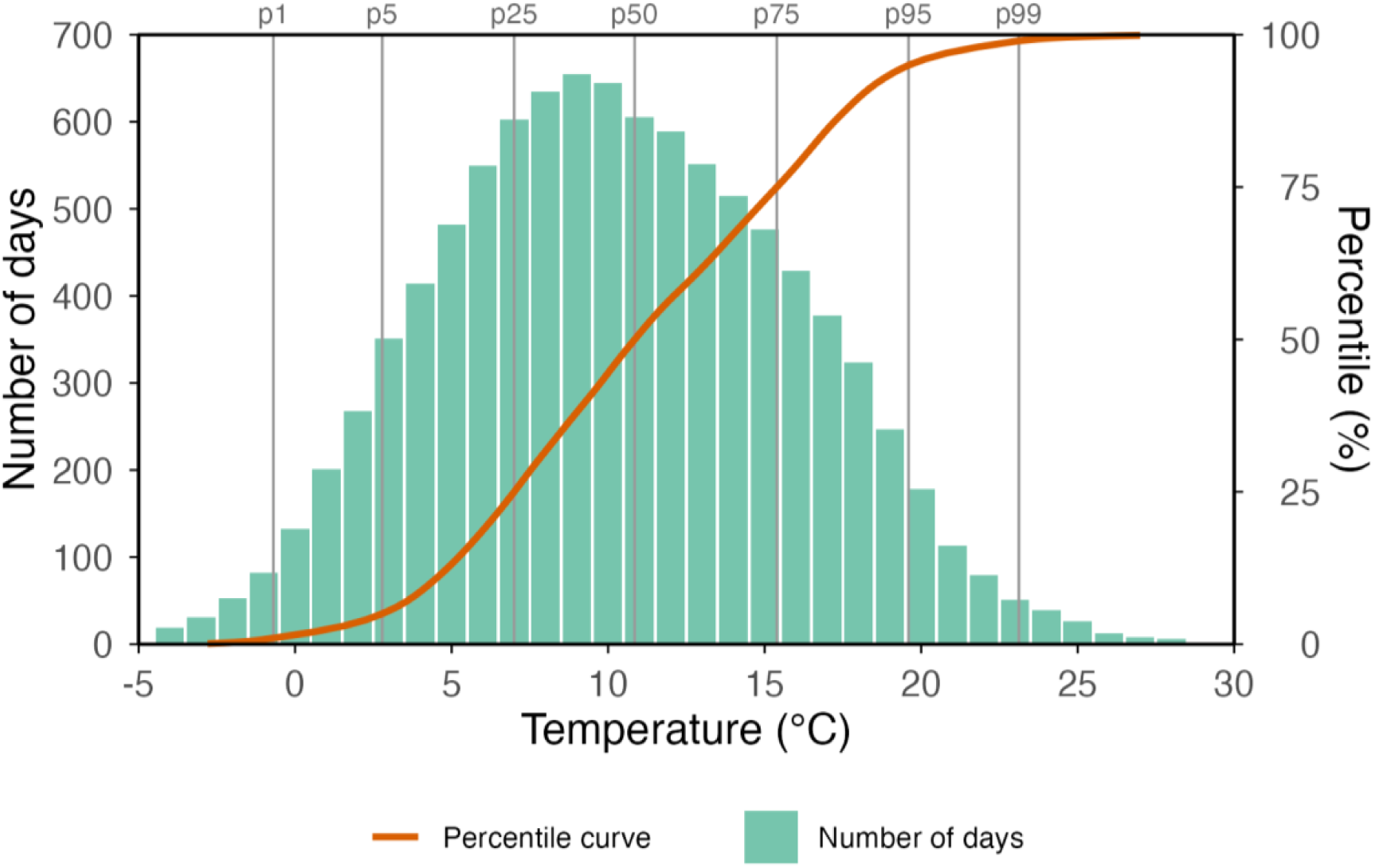
Daily mean air temperature distribution across England, 2020-2022. Bars show the number of calendar days on which each rounded daily mean temperature was recorded in at least one Lower Layer Super Output Area (LSOA) in England during the study period. The orange line shows the cumulative percentile distribution of daily mean temperatures across all LSOA- day observations. Vertical lines indicate the 1st, 5th, 25th, 50th, 75th, 95th and 99th percentiles of the national temperature distribution.

### Main analysis

Figure 2 displays cumulative IRRs for four thrombotic cardiovascular outcomes across daily mean air temperature, representing the relative risk over the 21 days following exposure compared with the median temperature. Corresponding model coefficients, events, and IRR values are provided in Table S11-15. A positive association between venous thrombotic events and high temperatures was found. Specifically, compared with days at the median temperature (10.8 °C), the risk of venous thrombotic events increased after days at higher temperatures, rising by 11% (IRR 1.11, 95% CI 1.04-1.18) at 19.6 °C (95th percentile) and by 33% (IRR 1.33, 95% CI 1.21-1.47) at 23.1 °C (99th percentile) (Table S12). Adjusting this analysis for time-varying covariates PM2.5 air pollution levels, COVID-19, or national lockdown, we found similar shapes of associations (Figure S4-6). No evidence of an association between hot or cold temperatures and the other outcomes was found (Figure 2, Tables S13-15).

**Figure 2:**
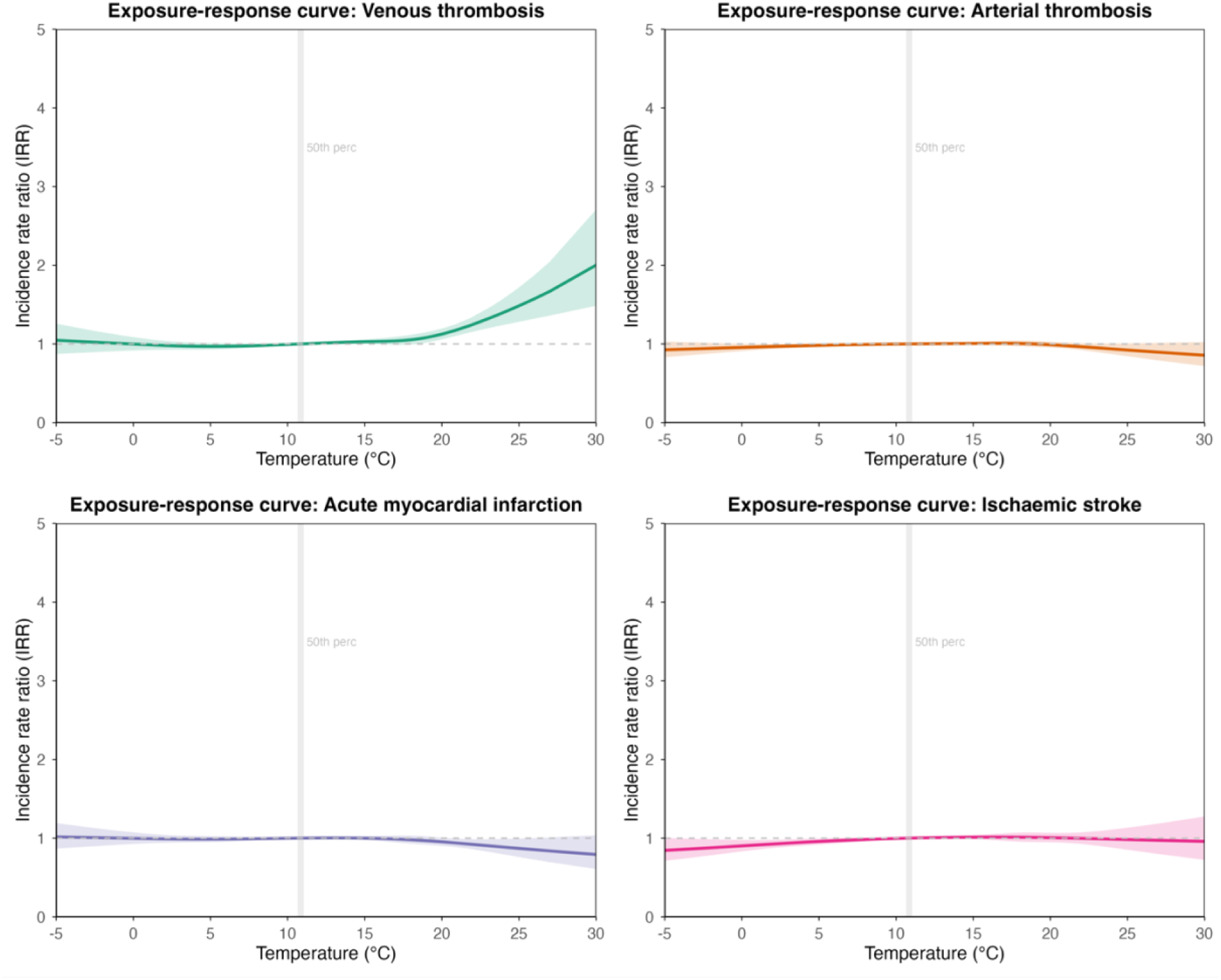
Exposure-response associations between daily mean air temperature and the cumulative risk of venous thrombotic event, arterial thrombotic event, acute myocardial infarction, and ischaemic stroke over lag days 0-21. Exposure-response curves show cumulative incidence rate ratios (IRRs) and 95% confidence intervals for the association between daily mean air temperature and each thrombotic outcome, estimated over lag days 0-21 using distributed lag non-linear models. Estimates are adjusted for seasonality and long-term temporal trends. The vertical grey line indicates the 50th percentile (10.8°C) of the national temperature distribution, which was used as the reference temperature (IRR = 1.0). The horizontal dashed line indicates an IRR of 1.0.

### Subgroup analysis

Stratified subgroup analysis revealed that recent COVID-19 (a diagnosis in the last six months), age, ethnic group, and pre-existing health conditions were significant effect modifiers (Figures 3, S7-19, Tables S12-15). Formal interaction tests are presented in Figure 3. Cold temperatures were associated with higher risk of venous thrombotic events over the following 21 days among individuals with a COVID-19 diagnosis in the prior six months (IRR 1.40, 95% CI 1.05-1.86; 1st percentile -0.7°C vs median 10.8°C) but not among individuals without recent COVID-19 (IRR 0.96, 95% CI 0.86-1.07; 1st percentile -0.7°C vs median 10.8°C) (Figure 3, S7, Table S12). Moreover, hot temperatures were associated with higher risk of venous thrombotic events over the following 21 days among individuals without recent COVID-19 (IRR 1.35, 95% CI 1.21-1.50; 99th percentile 23.1°C vs median 10.8°C) compared to individuals with a COVID-19 diagnosis in the prior six months (IRR 1.21, 95% CI 0.87- 1.67; 99th percentile 23.1°C vs median 10.8°C) (Figure 3, S7, Table S12).

**Figure 3:**
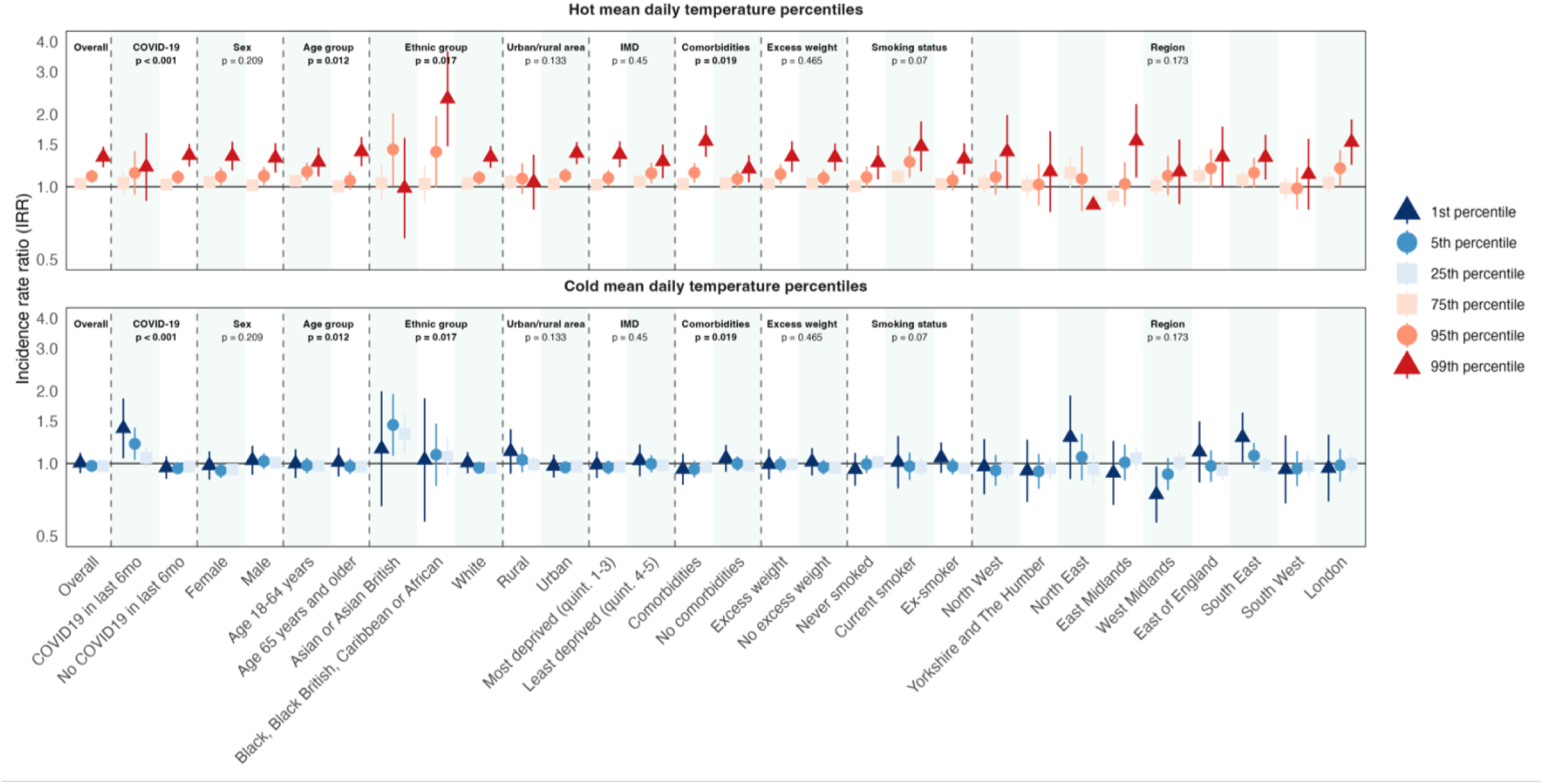
Associations between daily mean air temperature and the cumulative risk of venous thrombotic events at selected temperature percentiles across demographic, socioeconomic, clinical, COVID-19, and regional subgroups. Incidence rate ratios (IRRs) and 95% confidence intervals for venous thrombotic events are shown at selected hot (75th, 95th, and 99th percentiles; upper panel) and cold (1st, 5th, and 25th percentiles; lower panel) daily mean temperature percentiles, relative to the 50th percentile (reference). Estimates represent cumulative associations over lag days 0-21 and were adjusted for seasonality and long-term temporal trends. Results are presented for the overall population and stratified by demographic, socioeconomic, clinical, COVID-19, and regional subgroups. P-values correspond to tests for interaction between air temperature and each subgroup. The horizontal grey line indicates an IRR of 1.0.

Hot temperatures were also associated with higher risk of venous thrombotic events over the following 21 days among individuals aged ≥65 years (IRR 1.40, 95% CI 1.22-1.61; 99th percentile 23.1°C vs median 10.8°C) compared with individuals aged 18-64 years (IRR 1.27, 95% CI 1.10-1.45; 99th percentile 23.1°C vs median 10.8°C) (Figure 3, S9, Table S12). By ethnic group, the association was strongest among Black, Black British, Caribbean, and African individuals (IRR 2.32, 95% CI 1.47- 3.65; 99th percentile 23.1°C vs median 10.8°C), was also elevated among White individuals (IRR 1.33, 95% CI 1.20-1.48; 99th percentile 23.1°C vs median 10.8°C), but was not observed among Asian or Asian British individuals (Figure 3, S10, Table S12). Lastly, hot temperatures were associated with higher risk of venous thrombotic events over the following 21 days among individuals with pre-existing conditions (IRR 1.55, 95% CI 1.33-1.80; 99th percentile 23.1°C vs median 10.8°C) compared with individuals without pre-existing conditions (IRR 1.19, 95% CI 1.04-1.35; 99th percentile 23.1°C vs median 10.8°C) (Figure 3, S13, Table S12). No subgroup-specific temperature effects were observed for other cardiovascular outcomes over a 21-day lag period, and notably, air temperature affected thrombotic cardiovascular outcomes similarly in women and men (Figures S7-S19, Tables S13-S15).

### Impact analysis

An estimated 1,626 venous thrombotic events (95% CI 560-2,604) per year were attributable to moderate and extreme hot temperatures (temperatures above the 50th percentile) across the adult population of England and under the observed temperature conditions, equivalent to an attributable rate of 3.38 (95% CI 1.16-5.41) events per 100,000 person-years and 2.3% (95% CI 0.8-3.6%) of all events (Figure 4). No increased burden of venous thrombotic events was attributed to cold temperatures, nor could any burden of the other thrombotic cardiovascular outcomes consistently be attributed to air temperature (Table S16).

**Figure 4:**
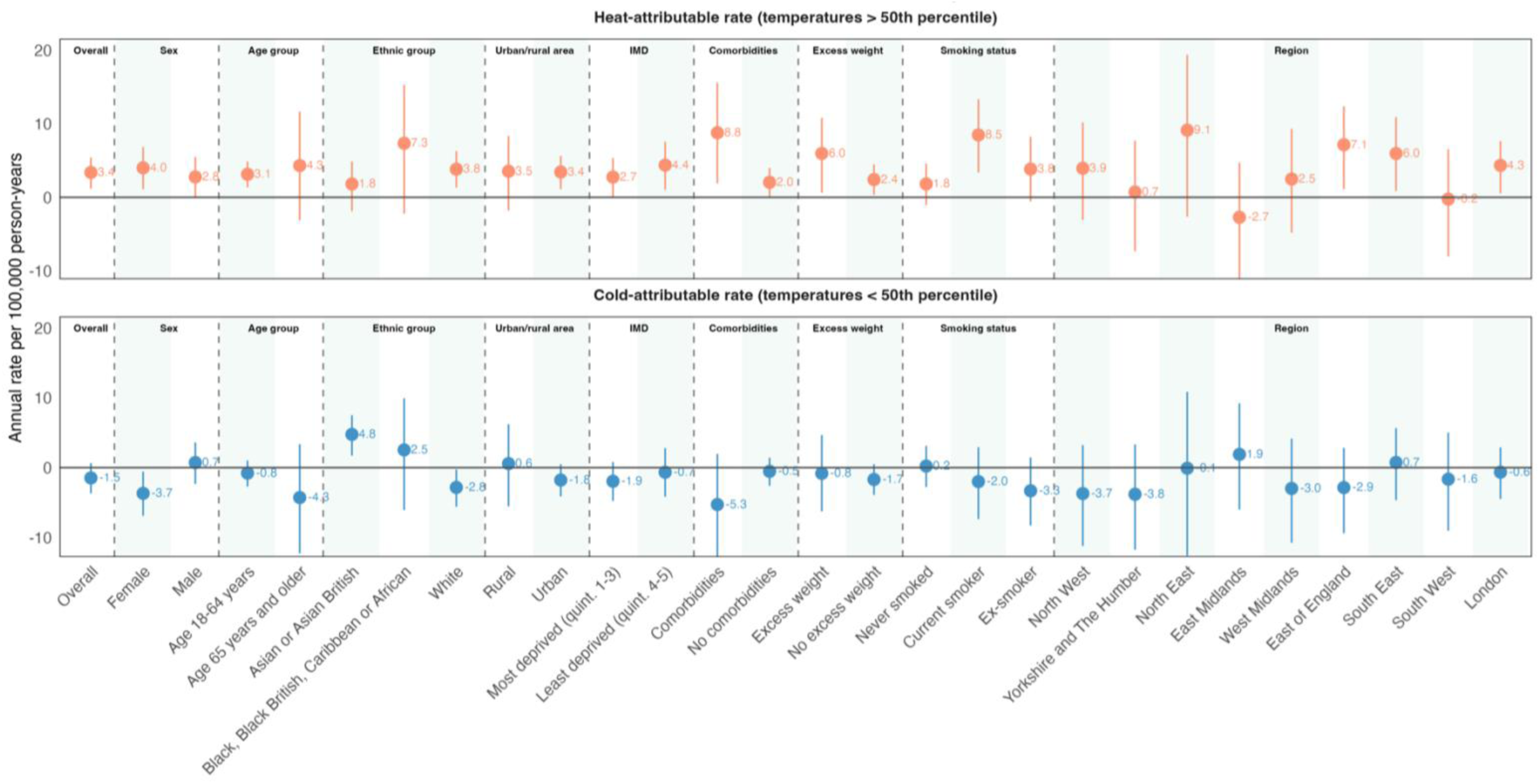
Heat- and cold-attributable rates of venous thrombotic events across demographic, socioeconomic, clinical, and regional subgroups. Annual heat- and cold-attributable rates of venous thrombotic events per 100,000 person-years, with 95% empirical confidence intervals, are shown for the overall population and stratified by demographic, socioeconomic, clinical, and regional subgroups. Heat-attributable rates were calculated for temperatures above the 50th percentile of the national daily mean temperature distribution (upper panel), and cold-attributable rates for temperatures below the 50th percentile (lower panel). Attributable rates were estimated using distributed lag non-linear models over lag days 0–21 and adjusted for seasonality and long-term temporal trends. Positive values indicate excess events attributable to heat or cold, whereas negative values indicate a reduction in events relative to the reference temperature. The horizontal grey line indicates an attributable rate of 0 events per 100,000 person-years.

Impact analysis by baseline characteristics of venous thrombotic event cases, showed an annual heat-attributable burden of 4.31 (95% CI -3.12-11.7) events per 100,000 person-years in individuals aged ≥65 years, 3.13 (95% CI 1.35-4.86) in individuals aged 18-64 years, 1.81 (95% CI - 1.89-4.86) in Asian or Asian British, 7.34 (95% CI -2.23-15.28) in Black, Black British, Caribbean and African, and 3.83 (95% CI 1.30-6.29) in White individuals. Cold was associated with an excess rate of 4.76 (95% CI 1.72-7.50) per 100,000 among Asian or Asian British individuals. Moreover, heat-attributable burden was 8.77 (1.87-15.55) per 100,000 person-years in those with pre-existing conditions, and 2.04 (0.03-3.99) per 100,000 person-years in those without (Figure 4, S20, Table S16).

### Sensitivity analysis

Restricting the study period to April 2020 onwards or including only first events yielded exposure-response curves of similar shape to the main analysis (Figures S21-22; Tables S17-S20). Fatal-event analyses showed no clear associations for most outcomes (Figures S23-S25), were based on limited sample sizes and were less precise (Table S11), except for fatal arterial thrombotic events, where colder temperatures were associated with increased risk (1st vs. 50th percentile IRR 1.20, 95% CI 1.05-1.37; Table S18). Censoring at the actual date of death resulted in higher relative risks at both temperature extremes for all outcomes (Figure S26), consistent with the potential bias described in Table S7. The association between temperature and venous thrombotic events was not observed when shorter lag periods (4- or 7-day) were applied (Figures S27-S28).

Over a 14-day lag, cold (1st percentile) was associated with an increased risk of arterial thrombotic events (IRR 1.08, 95% CI 1.03-1.14) and acute myocardial infarction (IRR 1.13, 95% CI 1.05-1.21) (Figure S29). Subgroup analyses for a 14-day lag period indicated that the cold-related effect on myocardial infarction was more pronounced in individuals aged ≥65 years (Figure 5, S30-31; Table S19). 14-day lag period impact analyses showed a significant annual cold-attributable burden of 10.18 (95% CI 2.21-17.84) acute myocardial infarctions per 100,000 person-years in those aged ≥65 years (Figure S32).

**Figure 5:**
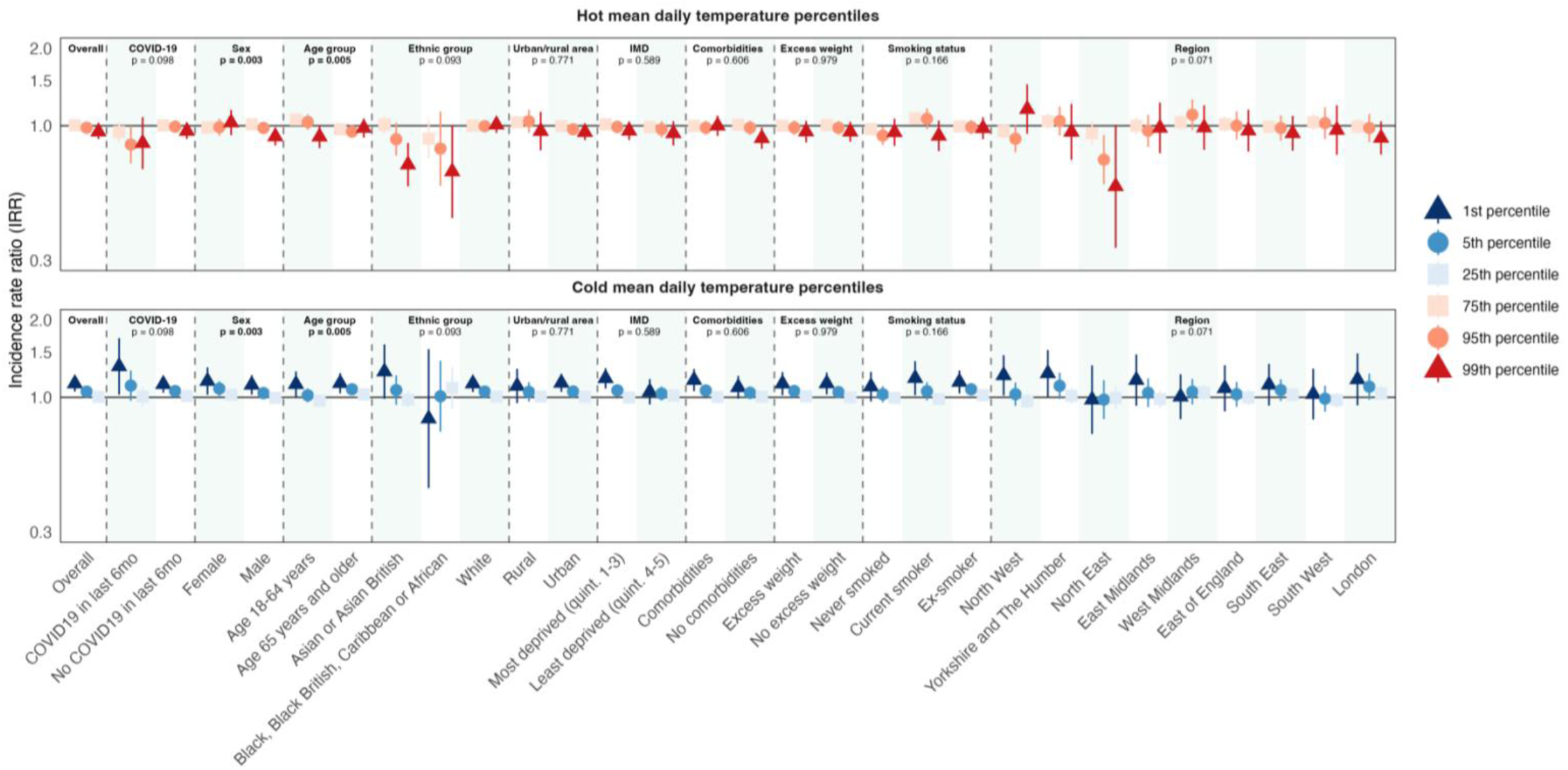
Associations between daily mean air temperature and the cumulative risk of acute myocardial infarction over lag days 0-14 at selected temperature percentiles across demographic, socioeconomic, clinical, COVID-19, and regional subgroups. Incidence rate ratios (IRRs) and 95% confidence intervals for acute myocardial infarction are shown at selected hot (75th, 95th, and 99th percentiles; upper panel) and cold (1st, 5th, and 25th percentiles; lower panel) daily mean temperature percentiles, relative to the 50th percentile (reference). Estimates represent cumulative associations over lag days 0-14 and were adjusted for seasonality and long-term temporal trends. Results are presented for the overall population and stratified by demographic, socioeconomic, clinical, COVID-19, and regional subgroups. P-values correspond to tests for interaction between air temperature and each subgroup. The horizontal grey line indicates an IRR of 1.0

## Discussion

Using whole-population electronic health records from over 49 million adults in England and high-resolution environmental data, we provide a novel assessment of the association between daily mean air temperature and thrombotic cardiovascular outcomes during the COVID-19 pandemic. Our findings suggest a temperature-sensitive pathway for venous thrombotic events, and possibly acute myocardial infarction, but no consistent associations were observed for arterial thrombotic events or ischaemic stroke. Extreme hot temperatures (99th percentile) were associated with an increased subacute (2-3 weeks) risk of venous thrombotic events. This effect was strongest in individuals of Black, Black British, Caribbean, or African ethnic group and those with pre-existing health conditions. An estimated 1,626 venous thrombotic events per year were attributable to heat (temperatures >50th percentile).

After recent COVID-19 diagnosis, we observed a relative higher risk of venous thrombotic events at extreme cold (1st percentile) temperatures instead. We also found a relatively higher risk of developing an acute myocardial infarction two weeks after exposure to extreme cold temperatures as well, particularly among older adults (≥65 years).

Hot temperatures were robustly associated with fatal and non-fatal venous thrombotic events, providing new evidence in a field with limited data. Previous research found an association between temperature extremes and pulmonary embolism, but not venous thrombosis^16^. Our study extends this evidence across a whole-population cohort and diverse subgroups. However, the delayed heat effect (2-3 weeks) contrasts with the typically reported immediate risk of heat^26,27^. This delay may suggest that heat may not act as an acute trigger but could gradually destabilise physiological homeostasis in vulnerable individuals. Heat-induced dehydration with progressive haemoconcentration provides a biologically plausible subacute pathway linking heat exposure to venous thrombosis^28^. Usual delays in diagnosis of deep vein thrombosis or pulmonary embolism may also contribute^29^.

We found evidence that colder temperatures were associated with a higher risk of acute myocardial infarction over the two weeks following exposure, while no associations between hot or cold air temperature and other arterial thrombotic outcomes were observed. Previous studies have reported hot and cold air temperature associations with ischaemic stroke, myocardial infarction, and other cardiovascular-related mortality; however, these associations have generally been more pronounced than those reported for non-fatal outcomes, including hospital admissions^3,9^. In sensitivity analyses restricted to fatal outcomes, although limited by small numbers, the exposure-response curve suggested a potential U-shaped association for all arterial thrombotic outcomes. These findings may indicate that short-term temperature effects are more pronounced among individuals already at high risk of death. Thus, extreme temperatures may accelerate death among those already nearing the end of life, which may also account for the immediate impact of heat exposure on health found in existing studies, but not in our study comprising predominantly non-fatal events and only a small number of fatal events due to the short period of follow-up.

Our analyses suggest that recent COVID-19 diagnosis may modify the association between air temperature and venous thrombotic outcomes. This is biologically plausible, as COVID-19 induces endothelial dysfunction and a prothrombotic state^30^, which may amplify susceptibility to cold-related cardiovascular stress. These mechanisms are not unique to COVID-19 and raise the possibility that other respiratory pathogens with vascular effects, such as influenza^31^, could similarly interact with environmental exposures to influence venous thrombotic risk.

Black, Black British, Caribbean, or African individuals experienced a stronger heat-associated increase in venous thrombotic risk than individuals in the other ethnic groups. As the case time series design adjusts by design for stable individual characteristics within each event month, inherited thrombotic susceptibility and baseline socioeconomic characteristics are unlikely to account for this finding. Instead, differences in adaptive capacity and other time-varying factors, such as behavioural responses to heat, occupational exposure, or housing conditions, are more plausible explanations.

This study has several key strengths. First, a major innovation is the linkage of whole-population electronic health records covering over 49 million adults in England, with high-resolution environmental data, specifically daily mean air temperature estimates on a 1 × 1 km grid assigned at the small-area (LSOA) level. Importantly, LSOA was implemented as a time-varying covariate: individuals’ residential locations were followed throughout the study period and updated whenever a new LSOA registration was recorded. This unprecedented integration of clinical and environmental datasets provides spatial precision and enables population-scale investigation of temperature-related cardiovascular risks. Second, the study uses a case time series design, allowing each individual to serve as their own control over time. This approach inherently adjusts for time-invariant confounding by baseline personal characteristics (e.g., age, sex, genetic factors, and baseline comorbidity), isolating the impact of within-person variation in exposure, and supports causal interpretation of the estimated associations, assuming the case time series assumptions are met and residual time-varying confounding is minimal. Access to individual-level sociodemographic and clinical information also allowed stratified analyses across demographic, social, lifestyle, and clinical subgroups, yielding novel insights into differential vulnerability to temperature-related thrombotic events.

This study has potential limitations that warrant consideration. First, temperature exposure was defined using outdoor daily mean air temperature, which may not fully reflect indoor condition and assignment was based on individuals’ registered residential LSOA. While this does not capture daily mobility patterns, it offers a considerably more refined exposure definition than the aggregated approaches that have typically been used in the field, and represents the most precise method currently feasible at national scale. Second, follow-up time was limited compared to existing environmental studies. Third, deprivation was available only at the area level, limiting interpretation of person-specific risk. In line with this, although the large cohort allowed for extensive subgroup analysis, further granularity (e.g., disaggregated ethnic subgroups or intersecting vulnerabilities such as ethnic group and deprivation) was constrained by insufficient sample sizes for model stability. Finally, although models adjusted for measured time-varying confounders, including day of week, COVID-19 cases, and air pollution, residual confounding from other unmeasured time-varying factors cannot be excluded.

To conclude, we showed that elevated daily mean air temperature was associated with a delayed increase in venous thrombotic events over 2-3 weeks and that viral respiratory disease, such as COVID-19, may modify this effect by shifting increased risk toward colder temperatures. Cold exposure was also associated with an increased risk of acute myocardial infarction. No consistent associations were observed for arterial thrombotic events, or ischaemic stroke. The unequal distribution of temperature-related cardiovascular events mirrors existing social and health inequalities, underscoring disproportionate risks among already disadvantaged groups. In the context of accelerating climate change, our results underscore the need for targeted public health and clinical strategies to protect at-risk populations from the cardiovascular consequences of non-optimal temperatures.

## Data Availability

This study made use of anonymised data held in NHS England Secure Data Environment service for England, and accessed via the BHF Data Science Centre CVD-COVID-UK/COVID-IMPACT Consortium. This work used data provided by patients and collected by the NHS as part of their care and support. All results produced are contained in the manuscript and supplemental materials. All code and codelists are publicly available on GitHub and the HDRUK Phenotype library (PH4045-PH4061).

https://github.com/BHFDSC/CCU076_01

## Acknowledgements

This work was supported by the British Heart Foundation Data Science Centre (grant SP/19/3/34678); awarded to Health Data Research UK. This study made use of anonymised data held in NHS England’s Secure Data Environment service for England, and made available via the BHF Data Science Centre’s CVD-COVID-UK/COVID-IMPACT Consortium. This work used data provided by patients and collected by the NHS as part of their care and support. We would also like to acknowledge all data providers who make health relevant data available for research. The BHF Data Science Centre’s Health Data Science Team provided data curation resources and support.

## Funding statement

This work was supported by the British Heart Foundation Data Science Centre (grant No SP/19/3/34678, awarded to Health Data Research (HDR) UK) funded co-development (with NHS England) of the Secure Data Environment service for England, provision of linked datasets, data access, and data management and wrangling support, to coordinate national COVID-19 priority research. Consortium partner organisations funded data access, user software licences and computational usage, and the time of contributing data analysts, biostatisticians, epidemiologists, and clinicians. This work was also supported by core funding from the British Heart Foundation (RG/F/23/110103), NIHR Cambridge Biomedical Research Centre (NIHR203312) [*], BHF Chair Award (CH/12/2/29428), Cambridge BHF Centre of Research Excellence (RE/24/130011), and by Health Data Research UK, which is funded by the UK Medical Research Council, Engineering and Physical Sciences Research Council, Economic and Social Research Council, Department of Health and Social Care (England), Chief Scientist Office of the Scottish Government Health and Social Care Directorates, Health and Social Care Research and Development Division (Welsh Government), Public Health Agency (Northern Ireland), British Heart Foundation and the Wellcome Trust.

*The views expressed are those of the authors and not necessarily those of the NIHR or the Department of Health and Social Care.

## Data Availability

The data used in this study are available in NHS England’s Secure Data Environment (SDE) service for England, but as restrictions apply they are not publicly available (https://digital.nhs.uk/services/secure-data-environment-service). The CVD-COVID-UK/COVID- IMPACT research programme, led by the BHF Data Science Centre (https://bhfdatasciencecentre.org/), received approval to access data in NHS England’s SDE service for England from the Advisory Group for Data (AGD) (https://digital.nhs.uk/about-nhs-digital/corporate-information-and-documents/advisory-group-for-data) – formerly the Independent Group Advising on the Release of Data (IGARD) – via an application made in the Data Access Request Service (DARS) Online system (ref. DARS-NIC-381078-Y9C5K) (https://digital.nhs.uk/services/data-access-request-service-dars/dars-products-and-services). The CVD-COVID-UK/COVID-IMPACT Approvals & Oversight Board (https://bhfdatasciencecentre.org/areas/cvd-covid-uk-covid-impact/) subsequently granted approval to this project (CCU076_01) to access the data within NHS England’s SDE service for England. The anonymised data used in this study were made available to accredited researchers only. Those wishing to gain access to the data should follow the application process of the relevant national data custodian.

## Notes

Author support: I.J.W. is supported by the Swedish Research Council for Sustainable Development (Formas; grant no. 2022-01845). A.S. is supported by the Medical Research Council (as part of UKRI) through the International Science Partnerships Fund (ISPF; MC_PC_24002). S.B.L. is supported by Health Data Research UK (HDRUK2023.0226). S.P. is supported by the Swedish Research Council for Sustainable Development (Formas; grant no. 2023-01774). L.O. is supported by the Swedish Research Council (Vetenskapsrådet; grant no. 2022-06599). M.M.O.N. is supported by the Swedish Research Council for Health, Working Life and Welfare (FORTE; grant no. 2024-00833) and the Swedish Research Council for Sustainable Development (Formas; grant no. 2022-01845). N.C. is supported by a Wellcome Trust Career Development Award (grant no. 318034/Z/24/Z), the Research Foundation Flanders (grant no. 12ZU922N), and KU Leuven internal funding. Y.L. is supported by the Wellcome Trust (grant no. 317540/Z/24/Z). S.I. is supported by Cancer Research UK (EDDAPA- 2024/100011). G.M. is supported by the Swedish Research Council (Vetenskapsrådet; grant no. 2022- 06599). A.G. is supported by the Wellcome Trust (grant no. 320878/Z/24/Z). A.W. is supported by the BHF Data Science Centre (HDRUK2023.0239), Health Data Research UK (Big Data for Complex Disease; HDR-23012), and as an NIHR Research Professor (NIHR303137). E.R. is supported by the Swedish Research Council for Sustainable Development (Formas; grant nos. 2022-01845 and 2023- 01774), the Swedish Research Council for Health, Working Life and Welfare (FORTE; grant nos. 2022-00882 and 2024-00833), and the Swedish Research Council (VR; grant nos. 2023-01982 and 2022-06599).

### Competing Interest Statement

The authors have declared no competing interest.

### Author Declarations

The North East-Newcastle and North Tyneside 2 research ethics committee provided ethical approval for the CVD-COVID-UK/COVID-IMPACT research programme (REC No 20/NE/0161) to access, within secure trusted research environments, unconsented, whole-population, de-identified data from electronic health records collected as part of routine healthcare.

## References

1. Semenza JC, Lindgren E, Balkanyi L, et al. Determinants and Drivers of Infectious Disease Threat Events in Europe. Emerg Infect Dis 2016; 22(4): 581–9.

2. Semenza JC, Paz S. Climate change and infectious disease in Europe: Impact, projection and adaptation. The Lancet Regional Health – Europe 2021; 9.

3. Liu J, Varghese BM, Hansen A, et al. Heat exposure and cardiovascular health outcomes: a systematic review and meta-analysis. Lancet Planet Health 2022; 6(6): e484–e95.

4. He Y, Liu WJ, Jia N, Richardson S, Huang C. Viral respiratory infections in a rapidly changing climate: the need to prepare for the next pandemic. EBioMedicine 2023; 93: 104593.

5. Copernicus. 2024 is the first year to exceed 1.5°C above pre-industrial level. 2025.

6. van Daalen KR, Tonne C, Semenza JC, et al. The 2024 Europe report of the Lancet Countdown on health and climate change: unprecedented warming demands unprecedented action. Lancet Public Health 2024.

7. IPCC. Weather and Climate Extreme Events in a Changing Climate. In: Change IPoC, ed. Climate Change 2021 – The Physical Science Basis: Working Group I Contribution to the Sixth Assessment Report of the Intergovernmental Panel on Climate Change. Cambridge: Cambridge University Press; 2023: 1513–766.

8. Gasparrini A, Masselot P, Scortichini M, et al. Small-area assessment of temperature-related mortality risks in England and Wales: a case time series analysis. Lancet Planet Health 2022; 6(7): e557–e64.

9. Achebak H, Rey G, Lloyd SJ, Quijal-Zamorano M, Méndez-Turrubiates RF, Ballester J. Ambient temperature and risk of cardiovascular and respiratory adverse health outcomes: a nationwide cross-sectional study from Spain. European Journal of Preventive Cardiology 2024; 31(9): 1080–9.

10. Wang X, Cao Y, Hong D, et al. Ambient Temperature and Stroke Occurrence: A Systematic Review and Meta-Analysis. Int J Environ Res Public Health 2016; 13(7).

11. Sun B, Singh N, Wigmann C, Singh N, Herder C, Schikowski T. Temperature and Inflammation: Investigating the impact of short-term exposure to ambient temperature on biomarkers of subclinical inflammation. Environmental Research 2025; 285: 122382.

12. Alahmad B, Khraishah H, Royé D, et al. Associations Between Extreme Temperatures and Cardiovascular Cause-Specific Mortality: Results From 27 Countries. Circulation 2023; 147(1): 35–46.

13. Ni W, Stafoggia M, Zhang S, et al. Short-Term Effects of Lower Air Temperature and Cold Spells on Myocardial Infarction Hospitalizations in Sweden. J Am Coll Cardiol 2024; 84(13): 1149–59.

14. Bhaskaran K, Hajat S, Haines A, Herrett E, Wilkinson P, Smeeth L. Short term effects of temperature on risk of myocardial infarction in England and Wales: time series regression analysis of the Myocardial Ischaemia National Audit Project (MINAP) registry. Bmj 2010; 341: c3823.

15. Alahmad B, Khraishah H, Kamineni M, et al. Extreme Temperatures and Stroke Mortality: Evidence From a Multi-Country Analysis. Stroke 2024; 55(7): 1847–56.

16. Di Blasi C, Renzi M, Michelozzi P, et al. Association between air temperature, air pollution and hospital admissions for pulmonary embolism and venous thrombosis in Italy. European Journal of Internal Medicine 2022; 96: 74–80.

17. Knight R, Walker V, Ip S, et al. Association of COVID-19 With Major Arterial and Venous Thrombotic Diseases: A Population-Wide Cohort Study of 48 Million Adults in England and Wales. Circulation 2022; 146(12): 892–906.

18. Kwong JC, Schwartz KL, Campitelli MA, et al. Acute Myocardial Infarction after Laboratory-Confirmed Influenza Infection. N Engl J Med 2018; 378(4): 345–53.

19. Nottmeyer L, Armstrong B, Lowe R, et al. The association of COVID-19 incidence with temperature, humidity, and UV radiation – A global multi-city analysis. Science of The Total Environment 2023; 854: 158636.

20. Met Office, Hollis D, Carlisle E, Kendon M, Packman S, Doherty A. HadUK-Grid Gridded Climate Observations on a 1km grid over the UK, v1.3.0.ceda (1836-2023). NERC EDS Centre for Environmental Data Analysis; 2024.

21. de la Cruz Libardi A, Masselot P, Schneider R, et al. High resolution mapping of nitrogen dioxide and particulate matter in Great Britain (2003-2021) with multi-stage data reconstruction and ensemble machine learning methods. Atmos Pollut Res 2024; 15(11): 102284.

22. Raffetti E, Bolton T, Nolan J, et al. COVID-19 diagnosis, vaccination during pregnancy, and adverse pregnancy outcomes of 865,654 women in England and Wales: a population-based cohort study. Lancet Reg Health Eur 2024; 45: 101037.

23. Gasparrini A. The Case Time Series Design. Epidemiology 2021; 32(6): 829–37.

24. Gasparrini A, Armstrong B, Kenward MG. Multivariate meta-analysis for non-linear and other multi-parameter associations. Stat Med 2012; 31(29): 3821–39.

25. Gasparrini A, Leone M. Attributable risk from distributed lag models. BMC Med Res Methodol 2014; 14: 55.

26. Guo Y, Gasparrini A, Armstrong B, et al. Global variation in the effects of ambient temperature on mortality: a systematic evaluation. Epidemiology 2014; 25(6): 781–9.

27. Ye X, Wolff R, Yu W, Vaneckova P, Pan X, Tong S. Ambient temperature and morbidity: a review of epidemiological evidence. Environ Health Perspect 2012; 120(1): 19–28.

28. Lacey J, Corbett J, Forni L, et al. A multidisciplinary consensus on dehydration: definitions, diagnostic methods and clinical implications. Ann Med 2019; 51(3-4): 232–51.

29. Ageno W, Agnelli G, Imberti D, et al. Factors associated with the timing of diagnosis of venous thromboembolism: Results from the MASTER registry. Thromb Res 2008; 121(6): 751–6.

30. Bonaventura A, Vecchié A, Dagna L, et al. Endothelial dysfunction and immunothrombosis as key pathogenic mechanisms in COVID-19. Nature Reviews Immunology 2021; 21(5): 319–29.

31. Babkina AS, Pisarev MV, Grechko AV, Golubev AM. Arterial Thrombosis in Acute Respiratory Infections: An Underestimated but Clinically Relevant Problem. Journal of Clinical Medicine 2024; 13(19).

