## Supplementary document for "Air temperature and thrombotic cardiovascular disease in England: case time series studies using whole-population electronic health records during 2020-2022"

Supplement version 6.

**Overview**

### **Supplementary methods**

**LSOA identification and definition**

For each individual in the cohort, we identified the Lower Layer Super Output Area (LSOA) recorded closest to the study baseline (1^st^ Jan 2020), prioritising records from before baseline. This LSOA is used to define baseline characteristics such as region of residence and socioeconomic deprivation (see *Baseline characteristics)*. We then extracted all available LSOA records throughout the study period from the following data sources: GDPPR (primary care), COVID-19 vaccination records, and HES data (secondary care).

In cases where multiple LSOA records occurred on the same date, we applied a hierarchical source preference to resolve ties:

GDPPR > COVID-19 vaccination > HES Admitted Patient Care > HES Outpatient > HES A&E Attendances.

If ties remained after applying this hierarchy, they were resolved randomly.

The final LSOA records were left-joined into the individual-level daily time series dataset. For individuals with multiple LSOA values across the study period, we filled in the periods between LSOA changes using the corresponding LSOA value, assigning each value up to the midpoint between recorded changes.

**Baseline characteristics**

Time-invariant variables were defined at baseline (1 January 2020) and included sociodemographic characteristics and pre-existing medical conditions. Sociodemographic variables were: age (continuous, but binary for stratification analysis: 18–64 years, ≥65 years), sex (female, male), ethnic group (Asian or Asian British; Black, Black British, Caribbean or African; Mixed or multiple ethnic groups; White ethnic group; Other ethnic group; missing), region of residence (North West, Yorkshire and the Humber, North East, East Midlands, West Midlands, East of England, South East, South West, London), urban-rural classification (rural conurbation, urban conurbation), and LSOA-based socioeconomic deprivation. Age is based on date of birth and age records from GDPPR, HES, COVID-19 vaccination data, or Sentinel Stroke National Audit Programme (SSNAP); sex is based on records from GDPPR, HES, or SSNAP; ethnic group is based on records from GDPPR or HES. The documentation on the methodology and the code to derive these variables can be found on the [BHF data science centre (DSC) github](https://github.com/BHFDSC/hds_curated_assets/blob/main/D07-demographics.py) and [Health Data Science Team Documentation](https://bhfdsc.github.io/documentation/).

Socioeconomic deprivation was measured using the 2019 English Index of Multiple Deprivation (IMD), linked from each individual’s residential Lower Layer Super Output Area (LSOA). IMD ranks were grouped into national quintiles (1=most deprived, 5=least deprived), and for the main analyses we categorised deprivation as more deprived (quintiles 1–3) versus less deprived (quintiles 4–5).

Pre-existing medical conditions were ascertained from primary and secondary care electronic health records using Systematized Nomenclature of Medicine - Clinical Terms (SNOMED CT) codes in the General Practice Extraction Service Data for Pandemic Planning and Research (GDPPR) and International Classification of Diseases, 10th Revision (ICD-10) codes in Hospital Episode Statistics (HES). Condition definitions followed phenotypes previously employed in the CVD-COVID-UK/COVID-IMPACT studies¹, most of which were originally adapted from the Health Data Research UK (HDRUK) Phenotype Library. Where necessary, new codelists were manually compiled and curated by two clinicians (IJW and LFB) (Table S2). The baseline conditions included history of hypertension, chronic kidney disease (CKD), diabetes, excess weight (overweight/obesity ICD-10 code or GDPPR-recorded BMI ≥25 kg/m²), depression, stroke, heart disease, smoking status (current, ex-smoker, never-smoker), and a composite comorbidity indicator. The composite indicator was defined as at least one of: diabetes, chronic obstructive pulmonary disease, asthma, liver disease, CKD, or cancer. The composite comorbidity indicator was created to enable stratification of the main analyses by comorbidity status, ensuring sufficient group sizes for robust comparisons. Stroke, heart disease, and CKD were based on manually curated code lists. Stroke as a baseline condition included ischaemic, not otherwise specified, and non-traumatic haemorrghagic stroke, and transient ischaemic attacks. Heart disease included heart failure, cardiomyopathy, coronary artery disease, congenital heart disease, pulmonary hypertension, valvular heart disease, aortic disease, arrhythmias, myocarditis, pericarditis, heart transplant, and hypertensive cardiac disease. The other conditions were based on the CVD-COVID-UK/COVID-IMPACT study by Raffetti et al. 2024.

Hypertension, depression, excess weight (ICD-10-based) and the composite comorbidity indicator were ascertained using a 5-year look-back period before baseline. Stroke, diabetes, heart disease and chronic kidney disease were defined using all available historical records (lifetime look-back). BMI measurements were taken from records 1.5 years before baseline to 3 months after baseline; where multiple values were available, the most recent measurement prior to baseline was prioritised.

Smoking status is defined as current, never, or former smoker based on SNOMED-coded records in GDPPR. To determine smoking status at baseline, records were selected according to a predefined temporal hierarchy relative to the baseline date. The time difference (x), expressed in years between the record date and the baseline date, was used to prioritise entries. Records dated within the 20 years preceding baseline (−20 ≤ x < 0) were considered first. If no eligible record was available during this period, records from up to three months after baseline (0 ≤ x < 0.25) were used. Thus, post-baseline information was incorporated only in the absence of relevant pre-baseline documentation. Given that smoking behaviour may change over time, earlier records were also reviewed to account for potential changes in classification. In particular, historical documentation indicating prior smoking was considered when more recent entries suggested never smoking, allowing individuals to be appropriately classified as former smokers where applicable.

**COVID-19 diagnosis**

The diagnosis of a COVID-19 was defined based on CVD-COVID-UK/COVID-IMPACT phenotypes by 1) a positive COVID-19 polymerase chain reaction or antigen test, 2) a confirmed COVID-19 diagnosis in primary healthcare records or 3) a confirmed COVID-19 diagnosis in secondary healthcare hospital admission records^1^.

COVID-19 diagnosis was examined as a binary effect modifier. Analyses were restricted to the first outcome event per individual. Individuals were classified according to whether they had a recorded COVID-19 diagnosis within the six months preceding the event (yes/no). This binary variable was added as an interaction term with air temperature in the Poisson regression.

**Time-varying covariates**

Daily PM2.5 air pollution was defined as a continuous time series with values of unit micrograms per cubic meter. To account for temporal and seasonal variation, three time variables were defined as daily series:

1) Calendar year (categorical variable with values 2020, 2021, and 2022); 2) Calendar week (integer values 1–52 or 1–53 depending on the year); and 3) Day of the week (integer values 1–7, representing Monday to Sunday).

In the Poisson regression models that were adjusted for COVID-19 status, COVID-19 diagnosis was included as a time-varying covariate. To avoid counting closely spaced diagnoses as separate events, a 42-day washout period was applied. Diagnoses occurring within 42 days of a previous COVID-19 record were considered part of the same episode, whereas diagnoses occurring more than 42 days later were treated as distinct episodes. Lockdowns were defined as the periods 23-03-2020 to 10-05-2020, 05-11-2020 to 02-12-2020, and 06-01-2021 to 08-03-2021^2^. In the daily time series lockdown is a binary indicator where any day within the national lockdown periods had value 1, all other days had value 0.

### **Study proposal**

| **BHF Data Science Centre: CVD-COVID-UK / COVID-IMPACT Project Proposal Form** | |
| --- | --- |
| **Project reference:** | CCU076 |
| **Project title:** | The effect of COVID-19 infection on cardiovascular outcomes: an interaction analysis with environmental exposure |
| **Proposal version:** | 1.1 |
| **Start date (best estimate):** | 01/11/2023 |
| **End date (best estimate)^[1]^:** | 31/11/2024 |
| **Named project lead and institution/organisation:** | Elena Raffetti, University of Cambridge  Rachel Denholm, University of Bristol  Angela Wood, University of Cambridge |

**Plain English summary**

- *Approx. 200 words^[2]^ overall to succinctly summarise the project in language suitable for a non-specialist lay public.^[3]^ This summary should be written using the headings below, with approximately one paragraph for each heading.*
- ***Please utilise your PPIE group to help write and/or review the summary before submitting your proposal****.*
- *See guidance on how to write a plain English summary* [*here*](https://www.nihr.ac.uk/documents/plain-english-summaries/27363#how-to-write-a-summary)

Describe the **challenge** or problem your project will address:

COVID-19 infection may become an annual winter virus and follow a clear seasonal pattern, like influenza. Environmental factors, such as extremes in temperature and pollution, could make it easier for the COVID-19 infection to increase the chances of getting a severe case of the virus. This has consequences for cardiovascular risk, as both COVID-19 infection and such environmental factors are known to increase the occurrence of cardiovascular events. However, no study has yet examined the combined effect of both COVID-19 infection and air temperature/pollution on cardiovascular risk.

What is the aim of your research and how will your project be the **solution** to address/understand the challenge or problem?

Our research aims to quantify how environmental factors, specifically air temperature and pollution levels, interact with COVID-19 and affect cardiovascular outcomes like heart attacks and strokes. We will also examine whether this interaction is higher in individuals with pre-existing heart conditions.

What is the potential **impact** from this work, e.g. how will it benefit patients/NHS, inform policy etc?

The findings from our research will offer valuable insights for the public, policy makers and healthcare providers. It will help identify who is at greatest risk of severe cardiovascular outcomes due to environmental factors and COVID-19 infection. This information can help us take specific actions to prevent problems. By helping to identify those at highest risk, our results will support more informed decision-making and inform planning and resource allocation for healthcare services. Global climate change underlines the importance of such research.

**Background**

- *Approx. 300 words^2^ summarising why the question(s) you are addressing matter, and how your project fits within the broad scope of* [*CVD-COVID-UK / COVID-IMPACT*](https://bhfdatasciencecentre.org/areas/cvd-covid-uk-covid-impact/)

Unprecedented international mobility facilitates the escalation of a local outbreak to an epidemic and in turn to a pandemic. Recent research points to the possibility of progressively more epidemics. Next to this, climate change has led to global warming and shifting distribution of temperature anomalies. Whilst heatwaves have increased, cold spells have not decreased as expected. The recent events in Western North America illustrate these dual aspects: an extreme heatwave (a high of 49.6 °C recorded in British Columbia, Canada) in mid-June 2021 followed by a record-cold wintertime temperature in December 2021.

New pathogens like COVID-19 and air temperatures extremes present new challenges for cardiovascular risk. Respiratory infections such as COVID-19 increase the risk of severe complications like myocardial infarction, cerebrovascular accidents, and other cardiovascular events. Similarly, exposure to non-optimal temperatures can adversely affect both short-term and long-term cardiovascular outcomes. Several underlying mechanisms may be involved (see Box 1).

Scholars hypothesise that COVID-19 infection may become a winter virus in an epidemic phase and follow a clear seasonal pattern, like influenza. Environmental factors may play a key role, both facilitating transmission of the infection and increasing the risk of severe SARS-CoV-2 infection. For example, increases in relative humidity may increase SARS-CoV-2 transmission rates, whilst shifts in air temperature increase the risk of more severe cardiovascular-related outcomes.

*Box. 1 How exposure to SARS-CoV-2 infection and non-optimal and extreme temperatures affects the cardiovascular risk*

**SARS-CoV-2 infection.** This infection may increase the risk of cardiovascular outcomes. Specifically, the viremia and inflammatory phase of the infection may increase metabolic and haemodynamic demands, induce a pro-coagulation and inflammatory state.

**Non-optimal and extreme temperatures**. Heat exposure causes direct cell damage (heat cytotoxicity), while blood vessel dilatation and sweating processes increase the risk of ischaemia, hypokalaemia and in turn arrhythmias. A core body temperature below 32°C is associated with spontaneous depolarisation of pacemaker cells that triggers cardiac arrhythmias such as ventricular fibrillation, a decrease of cardiac output, and a loss of cerebrovascular autoregulation.

**Air pollution.** The short-term effect of PM on the risk of cardiovascular event is related to an alteration of the autonomic tone and the direct effect of oxidative stress on the target cells. For example, dysfunction of the cardiomyocytes stem from an increased ROS (Reactive oxygen species) production and dysregulation of calcium. The oxidative stress also damages the endothelial cells leading to vasoconstriction.

*GAP IN KNOWLEDGE*

While existing studies have explored the individual effects of air temperature, pollution, and COVID-19 infection on cardiovascular risk, current studies overlook how these factors interact synergistically or cumulatively on cardiovascular outcomes. Furthermore, it remains unclear how these risk factors may disproportionately affect vulnerable populations such as the elderly or those with pre-existing conditions.

**Research question(s)**

**Research questions.** How does air temperature/air pollution modify the association between COVID-19 infection and cardiovascular outcomes? How does COVID-19 infection modify the association between air temperature/air pollution and cardiovascular outcomes?

**Novelty.** This project is the first to examine the compound effect of exposure to air temperature/pollution and COVID-19 infection on cardiovascular outcomes.

**Significance.** This has the potential impact to improve cardiovascular health reducing cardiovascular risk and healthcare costs by focusing on prevention and facilitating the development of new guidelines.

**Patient/public contributor involvement**

- *The BHF DSC works with patients and the public to ensure transparency, and to build trust in the use of health data for research. Please complete the relevant section below to indicate your plans for involving patient/public contributors throughout your project.*
- *Note: we expect you to engage in PPIE activities* ***prior to submitting your project proposal****. If you require assistance, please contact* *to request an initial discussion with the BHF DSC team and patient/public contributors, or for any other support with the involvement of patient/public contributors in your project.*

The research team **has** consulted with public/patients on plans for this project.

*Please provide brief details of a) any specific groups you are engaging with; b) how their involvement has influenced the project/research question(s); c) examples of how their involvement has brought about other changes in your project; and
d) how you will continue involving public/patients throughout the project and in the dissemination of outcomes.*

We held a “Brainstorming/Priority Setting” workshop on 13th July 2023 with 8 patient and public contributors from the NIHR Cambridge Biomedical Research Council (BRC) to discuss the use of medical, biological, environmental and social data together with new computer-based technologies for disease prevention, including cardiovascular disease. The workshop highlighted important aspects relevant to this proposal, including the need to consider (i) people with pre-existing diseases, and (ii) the translation and benefit of the research findings to individuals, health and social care providers, society in the UK and beyond. All 8 of the patient and public contributors expressed an excitement about the project in general, and particularly about advancing chronic disease prevention and having a wider view of contributing factors such as pollution.

We plan to involve patients/lay members from the BHF DSC, HDR UK, NIHR Cambridge BRC as well as individuals who have experienced COVID-19 during the project and have a strong interest in climate change and related matters (e.g. Cambridgeshire and Peterborough Climate Action Coalition).  Below are the key areas of the research in which we seek their active participation:

- *Identifying Research Priorities*: We will consult with the groups to prioritise research questions that are most relevant to their communities.
- *Design and Methodology*: These groups will provide input on the study design, ensuring that it is sensitive to the experiences and needs of patients.
- *Interpreting and Communicating Results*: Inputs from these groups will also be crucial in interpreting and communicating the results and ensuring ongoing feedback.

**Methods**

- *Provide a* ***brief overview*** *of methods to be used - a detailed plan is not required at this stage.*
- *Please also complete the ‘data analysts’ and ‘data sources’ tables below to confirm the analyst(s) who you propose will work with the data in the TRE(s), and for information on why the datasets/years of data are required.*

**STUDY DESIGN** A population-based cohort study using linked electronic health records from 1^st^ Jan 2020 to 31^st^ December 2023.

**STUDY POPULATION**

Individuals will be included if they meet ALL of the following criteria

- Alive on the study start date (1^st^ Jan 2020)
- Aged ≥18
- Known region and Lower Layer Super Output Area (LSOA)
- Have a record in the primary care extract at the start of the study period

**DATA SOURCES**

***NHS England Secure Data Environment for England***

- Primary care data (GP Data for Pandemic Planning and Research via General Practice Extraction Service, GPES);
- HES: Hospital Episode Statistics (Admitted Patient Care, Adult Critical Care, Outpatients, Accident & Emergency, APC Maternity file)
- NICOR CVD audits
- Emergency Care Data Set (ECDS);
- Pillar 1 and Pillar 2 COVID-19 infection laboratory testing data;
- Office of National Statistics (ONS) death registration records;
- ICNARC: Intensive Care National Audit and Research Centre;
- Medicines Dispensed in Primary Care (NHS BSA);
- COVID-19 vaccination data (including adverse events).

**Environmental data**

We will include air temperature (Daily mean, , based on Met Office data, 1x1 km resolution, availability period 2001-2022), air pollution (daily mean for PM 2.5, PM 10, NO2 based on re-analysis by LSHTM, 1x1 km resolution, availability period 2001-2021), h. For example, for an individual living in a specific small geographical area, defined on the based of LSOA, daily environmental data will be recorded as follows:

Date: September 10, 2021

Air Temperature: Daily Mean = 20°C, Min = 15°C, Max = 25°C

Diurnal Temperature Range: 10°C (25°C - 15°C)

Interday Temp. Variation: 2°C (compared to September 9, 2021)

PM 2.5: 12 µg/m³

PM 10: 20 µg/m³

NO2: 18 µg/m³

Humidity: 60%

Wind Speed: 5 m/s

Precipitation: 1 mm

This environmental data will be linked with the electronic health records of the individual based on their specific LSOA location. Cold and hot days will be defined in relation to the air temperature with the minimum effects for each adverse health outcome. The intensity of hot and cold temperatures will be categorised as moderate heat (≤97.5th percentile of the distribution over the study period), and extreme heat (>97.5th percentile), extreme cold (<2.5th percentile) and moderate cold (≥ 2.5th percentile), with a lag exposure of 7 days for hot temperatures and 21 days for cold temperatures. Reasonable variations of these thresholds will be tested.

***SARS-COV-2 infection***

SARS-COV-2 infection and severity definitions will be based on [CCU013](https://www.thelancet.com/journals/landig/article/PIIS2589-7500(22)00091-7/fulltext).

***SARS-COV-2 infection severity***

Individuals with a hospital admission record that includes confirmed SARS-CoV-2 infection in the primary position within 28 days of first SARS-CoV-2 infection will be defined as ‘SARS-CoV-2 infection with hospitalisation’. All other individuals will be defined as ‘SARS-CoV-2 infection without hospitalisation’.  Similar categorization would be done considering admission to critical care (admission to ICU and ventilated within 28 days of first SARS-CoV-2 infection).

**CARDIOVASCULAR OUTCOMES (based on CCU002)**

- Arterial events: first of ischaemic stroke or stroke or unknown type or myocardial infarction or retinal infarction
- Venous events: first of pulmonary embolism or deep venous thrombosis or cerebral sinus thrombosis
- Ischaemic stroke and stroke of unknown type
- Intracranial haemorrhage (subarachnoid and intracerebral)
- Pulmonary embolism
- Deep vein thrombosis
- Dissection or rupture of major arteries
- Life threatening cardiac arrhythmias + sudden cardiac death
- Cardiomyopathy (acute heart failure) / myocarditis

Date defined as: either date of event; OR first of date of start of spell with event; date of GP consultation with event; death with event.

We will also include negative control outcomes. Fractures for hot temperatures, since is an outcome that is unlikely to be affected by an infection and hot temperatures, and urinary tract infection for both hot and cold temperature.

**Analysis – general approach**

The simplest method is to compare the prognosis of patients with a positive coronavirus test with the population without a positive coronavirus test accounting for the potential effect modification by air temperature and pollution, before and after adjustment for confounders. In the initial stage of the analysis, we will utilise a Cox proportional hazards model, comparing outcomes of interest in individuals before or without COVID-19, and after COVID-19 infection incorporating environmental factors at the time of infection as interaction terms. This design is prone to confounding by risk factors for vascular diseases and coronavirus infection. A range of confounding variables will be accounted for in the model. To address the potential biases introduced by selective COVID-19 testing and vaccination status, these factors will also be incorporated into our analytical framework. Subsequently, we will employ time series and case-time series methodologies to quantify:

1. the effect modification of air temperature and pollution on the relationship between COVID-19 infection and cardiovascular outcomes.
2. the effect modification of COVID-19 infection on the association between air temperature, pollution, and cardiovascular outcomes.

Finally, stratified analyses will be conducted to evaluate these associations across age and ethnicity groups among individuals with and without history of pre-existing heart disease and multimorbidity.

**Data access: funding and conflicts of interest**

- *To make the provision of data for the research community via the consortium more sustainable, all projects (including those with a cardiovascular theme, where possible) are expected to contribute towards the recovery of the costs of data access, charged by the TRE providers to the BHF Data Science Centre.*
- ***Projects without a cardiovascular theme will need to fund the full cost of data access (c. £1k per user, per month, for access to English data only).***
- *Costs will be pro-rated where an analyst is working on more than one project.*
- *Funds should be set aside for post-submission access to data to address reviewer comments.*

Please confirm **whether** **funds are available to cover the full cost of data access** and provide detail about the source of the funds:

The project does not have funding to cover data access. Our analysts already access the environment with data access costs covered.

Please provide detail about any **potential conflicts of interest in accessing data for this project**, for example, whether you have been asked by and/or have received funds from an industry partner or other organisation to answer the research questions, or have links with an organisation that may have an interest in the outcomes, etc:

We have no conflict of interests.

**Trusted Research Environments (TRE)**
**England:** [NHS England’s Secure Data Environment (SDE) service](https://digital.nhs.uk/services/secure-data-environment-service)
**Scotland:** [Scottish National Data Safe Haven](https://www.isdscotland.org/Products-and-Services/eDRIS/) *(for more information, view the* [*COVID-19 Research Database Dataset and Variable Specification*](https://www.isdscotland.org/Products-and-Services/eDRIS/COVID-19/)*)*
**Wales:** [Secure Anonymised Information Linkage (SAIL) Databank](https://saildatabank.com/)
**Northern Ireland:** Northern Ireland Honest Broker Service (not yet available)

***** FOR COVID-IMPACT^[4]^ PROJECTS, PLEASE COMPLETE THE ANALYST AND DATA SOURCE DETAILS FOR ENGLAND ONLY *****

**DATA ANALYSTS**

|  | ***PLEASE COMPLETE THIS COLUMN*** |
| --- | --- |
| **TRE** | **Analyst(s) requiring TRE access – please provide name, institution, and email if not already a consortium member** |
| England | Isabel Walter, Rachel Denholm, Elena Raffetti, Alexia Sampri, Angela Wood |
| Scotland |  |
| Wales |  |
| Northern Ireland |  |

**DATA SOURCES**

|  |  |  |  |  | ***PLEASE COMPLETE THESE COLUMNS*** | | |
| --- | --- | --- | --- | --- | --- | --- | --- |
| **TRE** | **Category** | **Dataset Name** | **Year data available from** | **Available in TRE** | **Required**  **(X)** | **Years of data required (ALL or range)** | **Brief justification of why you need each dataset / date range** |
| England | Primary care | [**GDPPR: GPES Data for Pandemic Planning and Research**](https://digital.nhs.uk/coronavirus/gpes-data-for-pandemic-planning-and-research/guide-for-analysts-and-users-of-the-data) | From the start of each individual’s records^[5]^ | Yes | X | ALL | To measure covariates and outcomes for the analysis |
| England | Secondary care | [**HES: Hospital Episode Statistics**](https://digital.nhs.uk/data-and-information/data-tools-and-services/data-services/hospital-episode-statistics/hospital-episode-statistics-data-dictionary)  **- Admitted Patient Care** | 1997 | Yes | X | ALL | To measure covariates and outcomes for the analysis |
| England | Secondary care | **- Adult Critical Care** | 2013 | Yes | X | ALL | To derive COVID status and severity |
| England | Secondary care | **- Outpatients** | 2019 | Yes | X | ALL | To measure covariates and outcomes for the analysis |
| England | Secondary care | **- Accident & Emergency** | 2007 | Yes | X | ALL | To measure covariates and outcomes for the analysis |
| England | Secondary care | [**SUS: Secondary Uses Service**](https://digital.nhs.uk/services/secondary-uses-service-sus/payment-by-results-guidance) | 2019 / earlier | Yes | X | ALL | To measure covariates and outcomes for the analysis |
| England | Secondary care | **SUS/Uncurated Low Latency Hospital Data (Admitted Patient Care, Outpatients, Critical Care)** |  | Yes |  |  |  |
| England | Secondary care | [**Emergency Care Data Set (ECDS)**](https://web.www.healthdatagateway.org/dataset/7749d90e-dd7a-4697-8a9c-666491c64a0c) |  | Yes |  |  |  |
| England | COVID testing | [**COVID-19 SGSS: Second Generation Surveillance System**](https://digital.nhs.uk/about-nhs-digital/corporate-information-and-documents/directions-and-data-provision-notices/data-provision-notices-dpns/sgss-and-chess-data)^[6]^ | From start of records (2020) | Yes | X | ALL | To derive COVID status |
| England | COVID testing | [**Pillar 2 Antigen**](https://web.www.healthdatagateway.org/dataset/f06ec631-77d0-4b12-a21f-f11e7af49ba5) | April 2020 | Yes | X | ALL | To derive COVID status |
| England | COVID testing | [**Pillar 3 Antibody**](https://web.www.healthdatagateway.org/dataset/a77e52cc-d404-45d6-835f-32e2dc46778a) | September 2020 | Yes | X | ALL | To derive COVID status |
| England | COVID testing | **Variant strain data (COG-UK)** |  | Expected TBC |  |  |  |
| England | COVID vaccinations | [**Vaccination Status**](https://web.www.healthdatagateway.org/dataset/98372a87-f3e6-475d-be12-46c0f9587413) | December 2020 | Yes | X | ALL | To measure covariates and outcomes for the analysis |
| England | COVID vaccinations | [**Vaccination Adverse Reactions**](https://web.www.healthdatagateway.org/dataset/080a90e4-4bc6-4153-a4c6-c120eef5ba94) | December 2020 | Yes | X | ALL | To measure covariates and outcomes for the analysis |
| England | Deaths | **Civil Registration – Deaths**  ([ONS guidance](https://www.ons.gov.uk/peoplepopulationandcommunity/birthsdeathsandmarriages/deaths/methodologies/userguidetomortalitystatisticsjuly2017) /  [NHSD mortality data review](https://digital.nhs.uk/coronavirus/coronavirus-data-services-updates/mortality-data-review)) | 1993 | Yes | X | ALL | To derive COVID status and severity |
| England | ITU | **ICNARC: Intensive Care National Audit and Research Centre** |  | Yes | X | ALL | To derive COVID status and severity |
| England | ITU/HDU admissions | [**COVID-19 SARI-Watch (formerly CHESS: COVID-19 Hospitalisation in England Surveillance System)**](https://digital.nhs.uk/about-nhs-digital/corporate-information-and-documents/directions-and-data-provision-notices/data-provision-notices-dpns/sgss-and-chess-data) | From start of records (2020) | Yes | X | ALL | To derive COVID status and severity |
| England | Prescribing/ dispensing | [**Medicines Dispensed in Primary Care (NHS BSA)**](https://digital.nhs.uk/about-nhs-digital/corporate-information-and-documents/directions-and-data-provision-notices/data-provision-notices-dpns/nhs-bsa-medicines-data-provision-notice) | April 2015 | Yes | X | ALL | To measure covariates and outcomes for the analysis |
| England | Prescribing/ dispensing | **Secondary Care Prescribed Medicines (EPMA)** |  | Yes | X | ALL | To measure covariates and outcomes for the analysis |
| England | NICOR CVD audits | **NICOR – MINAP: Myocardial Ischaemia National Audit Project** |  | Yes | X | ALL | To measure MI outcome for the analysis |
| England | NICOR CVD audits | **NICOR – PCI: Percutaneous Coronary Interventions** |  | Yes | X | ALL | To measure cardiovascular outcomes for the analysis |
| England | NICOR CVD audits | **NICOR – NHFA: National Heart Failure Audit** |  | Yes | X | ALL | To measure cardiovascular outcomes for the analysis |
| England | NICOR CVD audits | **NICOR – NACSA: National Adult Cardiac Surgery Audit** |  | Yes | X | ALL | To measure cardiovascular outcomes for the analysis |
| England | NICOR CVD audits | **NICOR – NACRM: National Audit of Cardiac Rhythm Management** |  | Yes | X | ALL | To measure cardiovascular outcomes for the analysis |
| England | NICOR CVD audits | **NICOR – NCHDA: National Congenital Heart Disease Audit** |  | Yes | X | ALL | To measure cardiovascular outcomes for the analysis |
| England | NICOR CVD audits | **NICOR – TAVI: Transcatheter Aortic Valve Implantation** |  | Yes | X | ALL | To measure cardiovascular outcomes for the analysis |
| England | Stroke audit | **SSNAP: Sentinel Stroke National Audit Programme** |  | Yes | X | ALL | To measure cardiovascular outcomes for the analysis |
| England | National Vascular Registry | **National Vascular Registry Audit** |  | Expected  TBC |  |  |  |
| England | Other | [**Diagnostic Imaging Dataset**](https://web.www.healthdatagateway.org/dataset/86f46be3-bc29-4820-8363-de6530a94c6f) |  | Expected  TBC |  |  |  |
| England | Other | [**Improving Access to Psychological Therapies (IAPT)**](https://web.www.healthdatagateway.org/dataset/bcf6e5ce-986d-4b84-9c9c-69de966e8bbd) **v2.0 & v2.1** | Sep 2020 | Yes |  |  |  |
| England | Other | [**Maternity Services Dataset (MSDS)**](https://web.www.healthdatagateway.org/dataset/11d3cf0f-ae5d-42f4-a756-b98c3eff46bc) | April 2019 | Yes |  |  |  |
| England | Other | [**Mental Health Services Dataset (MHSDS)**](https://web.www.healthdatagateway.org/dataset/fa2ec63d-f76d-4555-8fc1-f4bd66e3c60d) | April 2019 | Yes |  |  |  |
| England | Other | [**Patient Reported Outcome Measures (PROMs)**](https://web.www.healthdatagateway.org/dataset/55614ed3-2484-4408-a4e9-e5fffed9e8e4) |  | Expected  TBC |  |  |  |
| Scotland | Primary care | [**Primary care**](https://web.www.healthdatagateway.org/dataset/8f486c3d-9504-4a0e-8e24-ab96f057fc9e)**^[7]^** |  | Yes |  |  |  |
| Scotland | Secondary care | [**Outpatient Appointments and Attendances - Scottish Morbidity Record (SMR00)**](https://web.www.healthdatagateway.org/dataset/04cb9964-54fb-4529-b09b-735c3daa1c7b) | 1997 | Yes |  |  |  |
| Scotland | Secondary care | [**General Acute Inpatient and Day Case - Scottish Morbidity Record (SMR01)**](https://web.www.healthdatagateway.org/dataset/98cda353-0011-45b2-80ca-4ed24cd084bf) | 1997 | Yes |  |  |  |
| Scotland | Secondary care | [**Accident & Emergency**](https://web.www.healthdatagateway.org/dataset/05e6752e-3a0b-4809-aa14-207b4761ef60) | 2007 | Yes |  |  |  |
| Scotland | COVID testing | [**COVID-19 laboratory and lighthouse testing (ECOSS)**](https://web.www.healthdatagateway.org/dataset/5542cadc-11a4-4957-bbdd-8a6c46bcfb87)**^[8]^** | From start of records (2020) | Yes |  |  |  |
| Scotland | COVID testing | **Covid Tests^[9]^** |  | Yes |  |  |  |
| Scotland | COVID testing | [**Variant strain data (COG-UK)**](https://web.www.healthdatagateway.org/dataset/69a356d6-599e-4d02-8241-066ce3c297bd) |  | Yes |  |  |  |
| Scotland | COVID vaccinations | **Vaccination data** |  | Yes |  |  |  |
| Scotland | Deaths | [**Deaths**](https://web.www.healthdatagateway.org/dataset/e600dae2-a83c-4b7a-8d23-af4ac31ca374) |  | Yes |  |  |  |
| Scotland | ITU | [**Intensive care data - Daily (SICSAG)**](https://web.www.healthdatagateway.org/dataset/811a54fb-8b79-442f-8e28-a725a0561a15)**^[10]^** |  | Yes |  |  |  |
| Scotland | ITU | [**Intensive care data - Episodes (SICSAG)**](https://web.www.healthdatagateway.org/dataset/6f8182d2-2993-400d-829d-b52cdb324bf3)**^[11]^** |  | Yes |  |  |  |
| Scotland | Prescribing/ dispensing | [**Dispensed/Prescribed/Paid**](https://web.www.healthdatagateway.org/dataset/22e3943e-edb5-44a1-9e4e-22b0f7a31767) **(Prescribing Information System)** | 2015 | Yes |  |  |  |
| Scotland | Stroke audit | **Scottish Stroke Care Audit** | TBC | Yes |  |  |  |
| Scotland | Other | **Diabetes covariates** |  | Yes |  |  |  |
| Scotland | Other | **Scottish Renal Registry^[12]^** | 2019 | Yes |  |  |  |
| Wales | Primary care | **GPCD: Welsh Longitudinal General Practice**  **(Daily COVID codes only)** | 2020 | Yes |  |  |  |
| Wales | Primary care | [**WLGP: Welsh Longitudinal General Practice**](https://web.www.healthdatagateway.org/dataset/33fc3ffd-aa4c-4a16-a32f-0c900aaea3d2) | 2000 | Yes |  |  |  |
| Wales | Secondary care | [**CCDS: Critical Care Dataset**](https://web.www.healthdatagateway.org/dataset/1789286a-deaf-49df-88cd-660e92934af6) | 2007 | Yes |  |  |  |
| Wales | Secondary care | [**EDDD: Emergency Department Dataset Daily**](https://web.www.healthdatagateway.org/dataset/fa257aa8-51fc-4eb7-a448-e473fa54686d) | 2010 | Yes |  |  |  |
| Wales | Secondary care | [**EDDS: Emergency Department Dataset**](https://web.www.healthdatagateway.org/dataset/75c4dcb8-33bf-43f4-b2bb-db51b6621b2c) | 2009 | Yes |  |  |  |
| Wales | Secondary care | [**OPDW: Outpatient Dataset for Wales**](https://web.www.healthdatagateway.org/dataset/d331159b-b286-4ab9-8b36-db39123ec229) | 2004 | Yes |  |  |  |
| Wales | Secondary care | [**OPRD: Outpatient Referral Dataset**](https://web.www.healthdatagateway.org/dataset/7465c65e-c321-42e9-a2f4-6bc664caf1fc) | 2009 | Yes |  |  |  |
| Wales | Secondary care | [**PEDW: Patient Episode Dataset for Wales**](https://web.www.healthdatagateway.org/dataset/4c33a5d2-164c-41d7-9797-dc2b008cc852) | 1995 | Yes |  |  |  |
| Wales | COVID testing | [**PATD: COVID-19 Test Results (Laboratory Information Management System [Pillar 1&2 NHS/Lighthouse Labs Results & Pillar 3 Antibody Results])**](https://web.www.healthdatagateway.org/dataset/f5f6d882-163d-4ef1-a53e-000fba409480) | March 2020 | Yes |  |  |  |
| Wales | COVID testing | [**CTTP: COVID-19 Test, Trace and Protect**](https://web.www.healthdatagateway.org/dataset/c0fc84af-aeab-4c7e-aa0b-8264baff55d9) |  | Yes |  |  |  |
| Wales | COVID testing | [**CVSP: COVID-19 Shielded People List**](https://web.www.healthdatagateway.org/dataset/dc22db56-8791-4e96-9ce6-6b6a58b1241f) | May 2020 | Yes |  |  |  |
| Wales | COVID testing | [**CVSD: COVID-19 Sequence Data**](https://web.www.healthdatagateway.org/dataset/602cbc9b-0113-4712-8c4a-0efe8d86b77c)**^[13]^** |  | Yes |  |  |  |
| Wales | COVID testing | **ONS COVID-19 Infection Survey^[14]^** |  | Expected TBC |  |  |  |
| Wales | COVID vaccinations | [**CVVD: Covid Vaccination Dataset**](https://web.www.healthdatagateway.org/dataset/471f101c-a45e-4620-8710-be3036a46fba) |  | Yes |  |  |  |
| Wales | Deaths | [**ADDD: Annual District Death Daily (ONS Deaths)**](https://web.www.healthdatagateway.org/dataset/584bf8c8-d58f-44c6-9b65-e1611144fd54) | 2016 | Yes |  |  |  |
| Wales | Deaths | [**ADDE: Annual District Death Extract (ONS Deaths)**](https://web.www.healthdatagateway.org/dataset/15cf4241-abad-4dcc-95b0-8cd7c02be999) | 1996 | Yes |  |  |  |
| Wales | Deaths | [**CDDS: COVID-19 Consolidated Deaths**](https://web.www.healthdatagateway.org/dataset/70e37f44-5c3e-4c83-a42a-89b3476d1d45) | 2019 | Yes |  |  |  |
| Wales | ITU | [**ICCD: ICNARC – Intensive Care National Audit & Research Centre *(COVID-19 only admissions)***](https://web.www.healthdatagateway.org/dataset/add6226b-0f21-439a-84a6-51dc26cdc425) | March 2020 | Yes |  |  |  |
| Wales | ITU | [**ICNC: ICNARC – Intensive Care National Audit & Research Centre *(All admissions)***](https://web.www.healthdatagateway.org/dataset/fa8bf269-4769-491b-b398-fa63137d14ab) |  | Yes |  |  |  |
| Wales | Prescribing/ Dispensing | [**WDDS: Wales Dispensing Dataset**](https://web.www.healthdatagateway.org/dataset/50ef6443-ed4b-40f9-97fb-1cfd53be6579) | 2015 | Yes |  |  |  |
| Wales | NICOR CVD audits | **NICO: NICOR Audits and Registers** |  | Expected TBC |  |  |  |
| Wales | Stroke audit | **HQIP: HQIP Stroke Audit** |  | Expected TBC |  |  |  |
| Wales | National Vascular Registry | **NVR: National Vascular Registry** |  | Expected TBC |  |  |  |
| Wales | Other | [**ADBE: Annual District Birth Extract**](https://web.www.healthdatagateway.org/dataset/12a77014-4a77-4eb1-8b26-f49689352d1b) | 1996 | Yes |  |  |  |
| Wales | Other | [**MIDS: Maternity Indicators Dataset**](https://web.www.healthdatagateway.org/dataset/a0c27454-8cbe-418f-bdae-a85d5e92e9d4) | 2014 | Yes |  |  |  |
| Wales | Other | [**NCCH: National Community Child Health**](https://web.www.healthdatagateway.org/dataset/20fe153c-a5e5-4991-900e-8fa9988e771a) |  | Yes |  |  |  |
| Wales | Other | [**CARE: Care Homes Index**](https://web.www.healthdatagateway.org/dataset/aae02f9e-1bad-415d-9730-c46a07a990aa) | 2018 | Yes |  |  |  |
| Wales | Other | [**CARS: (CARIS – Congenital Anomaly Register and Information Service)**](https://web.www.healthdatagateway.org/dataset/6fe7e004-eb51-4c7b-9905-1ec9a4827dca) | 1998 | Yes |  |  |  |
| Wales | Other | [**CENW: Office of National Statistics Census (2011)**](https://web.www.healthdatagateway.org/dataset/fc24482c-0f9b-445f-af7a-8598627c3b15)**^[15]^** | March 2011 only | Yes |  |  |  |
| Wales | Other | [**RTTD: Referral to Treatment Times**](https://web.www.healthdatagateway.org/dataset/0bf54842-c1db-4bdb-9f55-c5be490bf758) | 2012 | Yes |  |  |  |
| Wales | Other | [**SDEC: SAIL Dementia e-Cohort**](https://web.www.healthdatagateway.org/dataset/f48b7c05-6a44-480c-84c6-4f7a684f29f8) | March 2019 | Yes |  |  |  |
| Wales | Other | [**WASD: Welsh Ambulance Services NHS Trust**](https://web.www.healthdatagateway.org/dataset/d415a5c5-8432-4a81-937f-12135ade6de7) | 2013 | Yes |  |  |  |
| Wales | Other | [**WDSD: Welsh Demographic Service Dataset**](https://web.www.healthdatagateway.org/dataset/8a8a5e90-b0c6-4839-bcd2-c69e6e8dca6d) | 1990 | Yes |  |  |  |
| Wales | Other | [**WRRS: Welsh Results Reporting Service**](https://web.www.healthdatagateway.org/dataset/48ef8f54-9606-4e6f-9256-3cc07ffd10b8) |  | Yes |  |  |  |
| Northern Ireland |  | **TBC** |  | Expected TBC |  |  |  |

### **Protocol**

**Version history**

| **Version** | **Date** | **Changes** | **Reviewers** |
| --- | --- | --- | --- |
| V1.0 | 12-02-2024 | First internal draft | Isabel Walter, Elena Raffetti, Alexia Sampri. |
| V2.0 | 24-05-2024 | Second draft | Isabel Walter, Elena Raffetti,  Alexia Sampri. |
| V3.0 | 10-06-2024 | Third draft | Isabel Walter, Elena Raffetti,  Alexia Sampri. |

**Title**

Associations between air temperature, air pollution and cardiovascular health outcomes during the COVID-19 pandemic in England.

**Lay summary (<200 words)**

Both COVID-19 and environmental factors, such as air pollution and air temperature, increase the rates of cardiovascular events. However, current literature 1) is mainly focused on the effect of environmental factors on the spreading and severity of COVID-19, and not on how environmental factors impacted cardiovascular health during the pandemic. 2) Rarely includes person level data to examine the impact of environmental factors on cardiovascular health, and 3) Overlooks the interaction effect of COVID-19 and air temperature/pollution on cardiovascular risk. Climate change underlines the relevance of such research.

Our research aims to quantify the effect of environmental factors, specifically air temperature and pollution levels, on cardiovascular outcomes like heart attacks and strokes

during the COVID-19 pandemic and how environmental factors interact with COVID-19 and affect cardiovascular outcomes like heart attacks and strokes. It will help identify who is at greatest risk of severe cardiovascular outcomes due to environmental factors and who is most vulnerable during a pandemic such as COVID-19. This will support more informed decision-making regarding planning and resource allocation for healthcare services.

**Background**

The increasing emergence of infectious diseases over the last decades, exemplified most recently by the COVID-19 pandemic, is influenced by various factors such as population size, mobility, urbanisation, and potentially climate change^3^. This highlights the intricate interplay between human activity and environmental conditions.

As we grapple with the health implications of new infectious diseases, we are also confronted with the challenge of climate change. The last decade alone has seen the 10 warmest years in a 174-year record with the current global mean temperature of 1.18°C above pre-industrial level^4,5^. Changing demographics such as an ageing population, and an increased incidence of non-communicable diseases, heighten the vulnerability of the population to temperature extremes. Combined with rising temperatures, this drives an increase in global heat-related mortality, one-third of which are attributable to anthropogenic climate change^6^. While cold spells are expected to become milder and less frequent^7,8^, their health burden is likely to remain higher in some regions than that of heatwaves for several decades, and cold-related mortality may increase locally on the background of a warming climate^9^.

Considering cardiovascular-specific outcomes, both extreme heat and cold are related with a higher risk of ischaemic heart disease, stroke, heart failure, and cardiovascular death as compared to the temperature associated with least mortality^10^. Air pollution, especially PM 2.5, is associated with cardiovascular disease, such as myocardial infarction and heart failure^11^. Emerging pathogens, such as SARS-CoV-2, present new challenges for cardiovascular risk as well. Indeed, COVID-19 has been found to be associated with myocardial infarction, stroke, heart failure and atrial fibrillation^12^.

The wealth of health data collection during the COVID-19 pandemic provides the unique opportunity to investigate 1) the association between environmental factors and cardiovascular health outcomes using person level data in a time period of restricted human mobility, and 2) the interaction effect of viral respiratory disease and environmental factors^12,13^. Mechanistic interaction, also known as biological interaction or synergism, suggests that

the presence of one exposure can change the way another exposure affects a health outcome^14^. Therefore, it is expected that environmental factors and viral respiratory diseases

may interact, influencing their effects on cardiovascular outcomes.

This study is relevant because existing studies mainly explore the association between air temperature/air pollution and COVID-19 severity and spreading^15-21^. However, they do not address how these environmental factors affect cardiovascular risk during a pandemic. Additionally, most research on the association between air temperature/pollution and cardiovascular disease relies on aggregated data approaches^10,11,22,23^. While the individual effects of air temperature, air pollution, and COVID-19 on cardiovascular risk have been studied, current evidence overlooks how these factors interact synergistically or cumulatively on cardiovascular outcomes. Furthermore, it remains unclear how these risk factors may disproportionately affect vulnerable populations such as the elderly or those with pre-existing conditions.

Assessing the association between COVID-19 and cardiovascular outcomes is not an objective of this analysis, as this has been extensively studied by the CVD-COVID-UK consortium^1^.

**Research aims**

1) Estimate to what extent air temperature and air pollution are associated with cardiovascular outcomes during the COVID-19 pandemic, and assess drivers of vulnerability.
2) Estimate to what extent environmental factors and COVID-19 interact in their association with cardiovascular outcomes.

This project focusses on short-term health effects.

**Study design**

A case time series of a population-based cohort using linked electronic health records from 1st Jan 2020 to 31st December 2023.

**Study population**

Individuals will be included if they meet each of the following criteria:

•	Alive on the study start date (1st Jan 2020).

•	Aged ≥18 years.

•	Known deprivation index and therefore a known lower layer super output area (LSOA) within England.

•	Have a record in the primary care extract at the start of the study period.

**Datasets for analysis**

*NHS England Secure Data Environment for England*

- Primary care data from the General Practice Extraction Service Extract for Pandemic Planning and Research (GDPPR);
- Secondary uses service (SUS): Secondary health care data that is updated in real-time. This is processed to create Hospital Episode Statistics data (see below).
- Hospital Episode Statistics: Admitted Patient Care (HES-APC), Adult Critical Care (HES-CC), Outpatients (HES-OP), Accident & Emergency (HES-AE);
- COVID-19 laboratory testing data from the Public Health England Second Generation Surveillance System (SGSS) including 1) tests conducted based on clinical need and for healthcare personnel (Pillar 1) and 2) tests from the wider population (Pillar 2);
- COVID-19 Hospitalisations in England Surveillance System (CHESS);
- Office of National Statistics (ONS) death registration records;
- Medicines Dispensed in Primary Care (NHS BSA);
- COVID-19 vaccination data (including adverse events).

*Environmental data*

The environmental data includes air temperature (daily mean), air pollution (daily mean for PM 2.5, PM 10, NO2 based on re-analysis by LSHTM, 1x1 km resolution, availability period 2001-2021). For example, for an individual living in a specific small geographical area, defined on the based LSOA, daily environmental data will be recorded as follows:

Date: September 10, 2021

Air Temperature: Daily Mean = 20°C

PM 2.5: 12 µg/m³

PM 10: 20 µg/m³

NO2: 18 µg/m³

**Phenotype definitions**

*Air temperature*

Air temperature is defined as mean daily temperature linked to the electronic health records of each person based on their specific LSOA location on the corresponding date. We will also define cold spells and heatwaves as exposure.

*Air pollution*

Two measures of air pollution are used: daily mean PM2.5 and daily mean PM10. This information is linked to the electronic health records of each person based on their specific LSOA location on the corresponding date. Any analysis using air pollution data will be based on the time period from 1^st^ Jan 2020 until 31^st^ Dec 2021, because the air pollution data covers time until 2021.

*COVID-19*

COVID-19 definition will be partially based on [CCU013](https://www.thelancet.com/journals/landig/article/PIIS2589-7500(22)00091-7/fulltext)^24^. COVID-19 diagnosis can be defined by 1) a positive SARS-CoV-2 test, 2) COVID-19 diagnosis based on SNOMED-CT codes in primary care, and 3) hospital admission with COVID-19 diagnosis based on ICD-10 codes.

*Time invariant covariates*

These covariates are determined at baseline and used to study potential effect modification if the number of events in the subgroups are sufficient. They include age group at event (categories in years: 18 ≤ age < 40, 40 ≤ age < 60, 60 ≤ age), sex (categories: female, male), ethnicity (six groups: Asian or Asian British, Black or Black British, White, Mixed, Other Ethnicity, Unknown ethnicity), socioeconomic deprivation index (categories: low ≤2nd quintile, high ≥3rd quintile), smoking status (three categories: smoker, former smoker, non-smoker), history of hypertensive disorders (categories: yes or no), history of heart disease (categories: yes or no), history of being overweight/obesity (categories: yes or no), history of depression (categories: yes or no) and a composite of other comorbidities (categories: yes or no). The composite includes at least one of the following health conditions: history of diabetic disorders, chronic obstructive pulmonary disease, liver disease, chronic kidney disease, cancer and surgical intervention.

*Time varying covariates*

These covariates are collected in a daily time series like air temperature and air pollution. They include age in years, calendar week, day of week.

*Cardiovascular outcomes*

Outcome events are defined using ICD-10 codes. As sensitivity analysis both ICD-10 and SNOMED-CT codes will be included.

- Arterial events composite: first of ischaemic stroke, stroke of unknown type, myocardial infarction, arterial retinal infarction, arterial dissection or ruptured arterial aneurysm, and any other arterial thrombosis.
  - Ischaemic stroke and myocardial infarction will additionally be evaluated as separate, non-composite outcomes.
- Venous events composite: first of pulmonary embolism, deep venous thrombosis, intracranial venous thrombosis.

Date defined as: either date of event; OR first of date of start of spell with event; date of GP consultation with event; death with event.

We will use appendicitis as a negative control outcome.

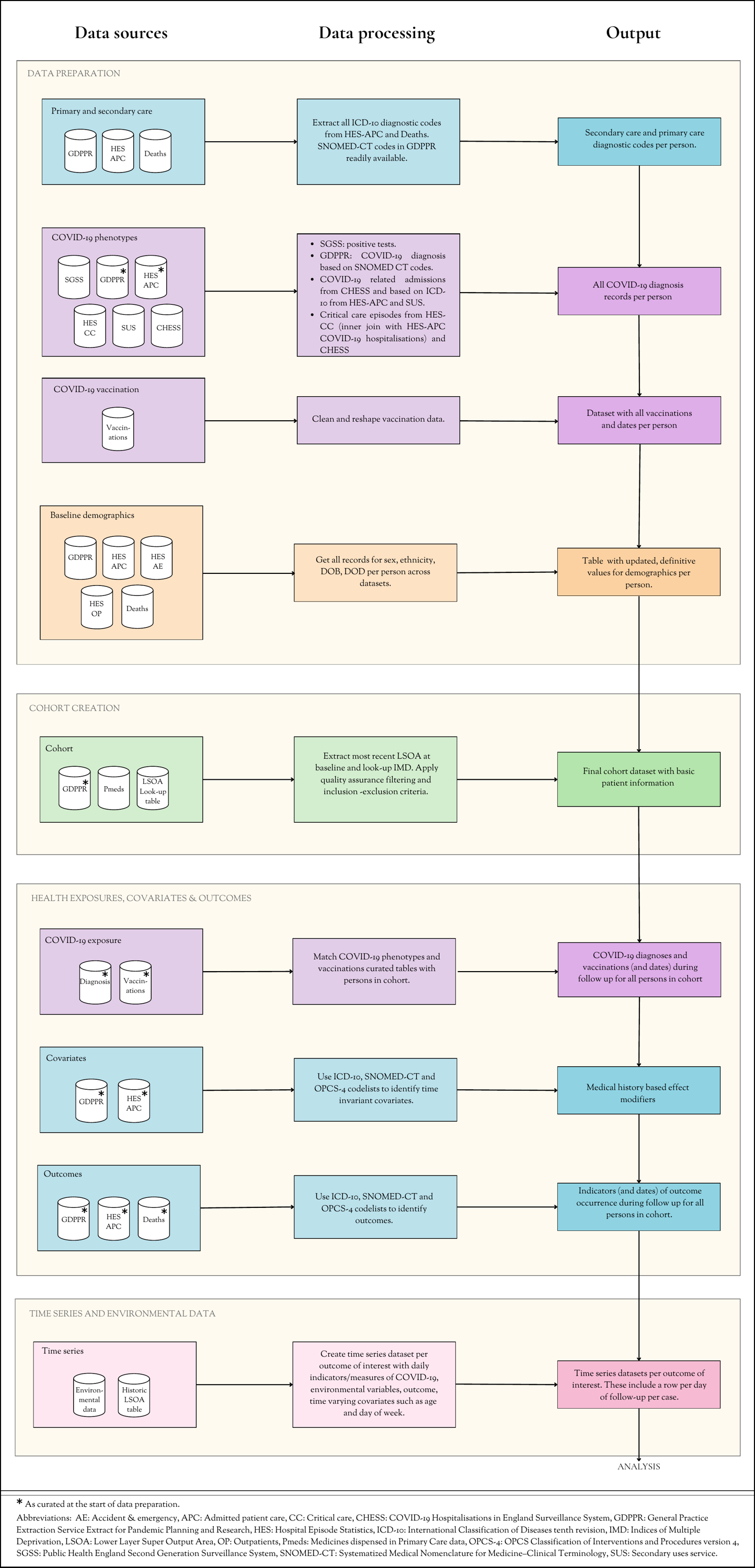

**Figure I. Data curation pipeline.**

**Cohort derivation**

The steps in data curation for analysis are presented in Figure I. A cohort with basic patient demographics is built based on primary care data, then curated COVID-19, deaths registry, primary care and secondary care datasets are used to expand the information in this cohort with relevant exposures, time invariant covariates and outcomes. Next, cases are identified for each of the outcomes of interest and time series datasets are created per outcome. A time series dataset includes only the cases of a specific outcome (e.g. arterial event). Per case it includes daily time series of LSOA, exposures, outcome and time-varying covariates. This is a long dataset with a row for each day of follow-up time per case. Follow-up time is defined as the calendar month in which the first outcome event took place. Variables (columns) are:

person ID, age in years, date, calendar week, day of week, outcome indicator, COVID-19 indicator, daily mean air temperature, daily mean PM2.5, and daily mean PM10.

**Statistical analysis**

Cohort demographics will be presented in a baseline characteristics table with descriptive statistics presented as numbers and percentages, stratified for COVID-19 status (no COVID-19 during follow-up and at least once COVID-19 during follow-up).

*Case time series*

Initially, we aim to assess the association between air temperature/pollution and cardiovascular outcome. Then we compare the prognosis of patients with a COVID-19 diagnosis to the population without a COVID-19 diagnosis, accounting for the potential interaction between COVID-19 and air temperature and air pollution. To address these objectives, a case time series analysis for each of the outcomes of interest will be performed.

A case time series allows for temporality found in time series with individual-level adjustment for time-varying confounders, while time invariant confounders are adjusted for by design.^25^

Daily time series are created for individuals who have the outcome of interest during study follow-up, as described in *Cohort derivation*. These time series include the calendar month when the first outcome event happened; each individual will have between 28 to 31 observation days. A Poisson regression model is fit to estimate the incidence relative risk of the outcome of interest.

A distributed lag model will be implemented to model COVID-19, air temperature, air pollution, and relevant interaction terms with a lag period defined based on current evidence. For COVID-19 a lag period of day 1-7 days and 8-28 days are used, to assess both the acute and subacute effects of infection. For air temperature a lag period of 0-1 days, 2-7 days and 8-28 days are used, to be able to assess immediate effects of hot temperatures and more delayed effects of cold temperatures^26^. For air pollution, the lag period is defined as 0-5 days, as short-term effects of air pollution on cardiovascular events are found to be most pronounced for these days^27^. Terms to adjust for calendar week and day of the week are added as continuous variables.

We will start by modelling: **Base model 1**) The association between air temperature and each outcome of interest**. Base model 2**) The association between air pollution and each outcome of interest. **Base model 3**) The association between COVID-19 and each outcome of interest. Figure II illustrates what the time series data could look like for three example subjects for base model 1. Results from model 1-3 will be compared with previously published evidence on the topic, where alternative methods were applied. Then, adjustments and interactions are added as described in Table I.

**Figure II. Time series data of three example subjects for base model 1. Yellow depicts the continuous air temperature exposure over the selected follow-up time. The blue circle is a binary indicator of the outcome during the selected follow-up time.**

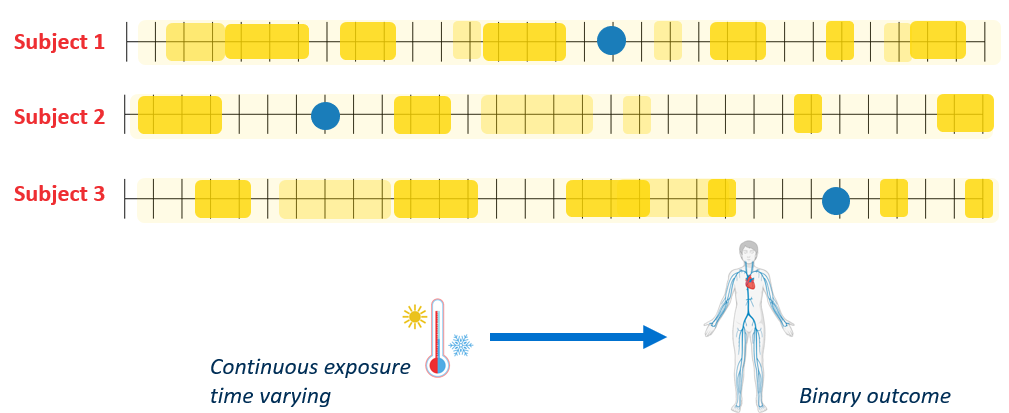

| Table I. Overview statistical modelling steps per aim. | | | |
| --- | --- | --- | --- |
| Objective | Base model | Adjustment | Interaction |
| Estimate to what extent air temperature is associated with cardiovascular outcomes during the COVID-19 pandemic. | 1 | Adjustment: Calendar week, day of week. | Not applicable. |
| Estimate to what extent air pollution is associated with cardiovascular outcomes during the COVID-19 pandemic. | 2 | Adjustment: Calendar week, day of week. | Not applicable. |
| Estimate to what extent COVID-19 modifies the association between air temperature and cardiovascular outcomes. | 1 | Adjustment: Calendar week, day of week. | Add interaction between air temperature and COVID-19 as time fixed covariate. |
| Estimate to what extent air temperature modifies the association between COVID-19 and cardiovascular outcomes. | 3 | Adjustment: Calendar week, day of week. | Add interaction between COVID-19 and average temperature 21 days before the event as time fixed covariate. |
| Estimate to what extent COVID-19 modifies the association between air pollution and cardiovascular outcomes. | 2 | Adjustment: Calendar week, day of week. | Add interaction between air pollution and COVID-19 as time fixed covariate. |
| Estimate to what extent air pollution modifies the association between COVID-19 and cardiovascular outcomes. | 3 | Adjustment: Calendar week, day of week. | Add interaction between COVID-19 and average pollution 7 days before the event as time fixed covariate. |

*Subgroup analysis*

Subgroup analyses are planned to assess potential effect modifiers described in the *time invariant covariates* section.

*Sensitivity analysis*

Planned sensitivity analyses are: 1) Include cases who died during the selected study follow-up. 2) A comparison between using ICD-10 codes to define outcomes of interest and using both ICD-10 and SNOMED-CT codes. 3) Extend the follow-up time (15 days in the previous and following calendar month) to examine associations up to 45 days from COVID-19 infection. 4) A comparison of the association between air temperature/pollution and cardiovascular outcome between national lockdown and non-lockdown periods.

If time allowed:

5) A comparison of estimated effects on cardiovascular outcome between COVID-19 vaccination status group at the outcome event, using the following definition:

Group 1: Never had COVID-19.

Group 2: COVID-19 and A) vaccinated with one or less doses OR B) previous COVID-19.

Group 3: COVID-19 AND vaccinated with 2 or more doses AND a previous COVID-19 infection.

6) Estimate the threshold of effect of air temperature between non-optimal cold and hot temperature.

### **Figures**

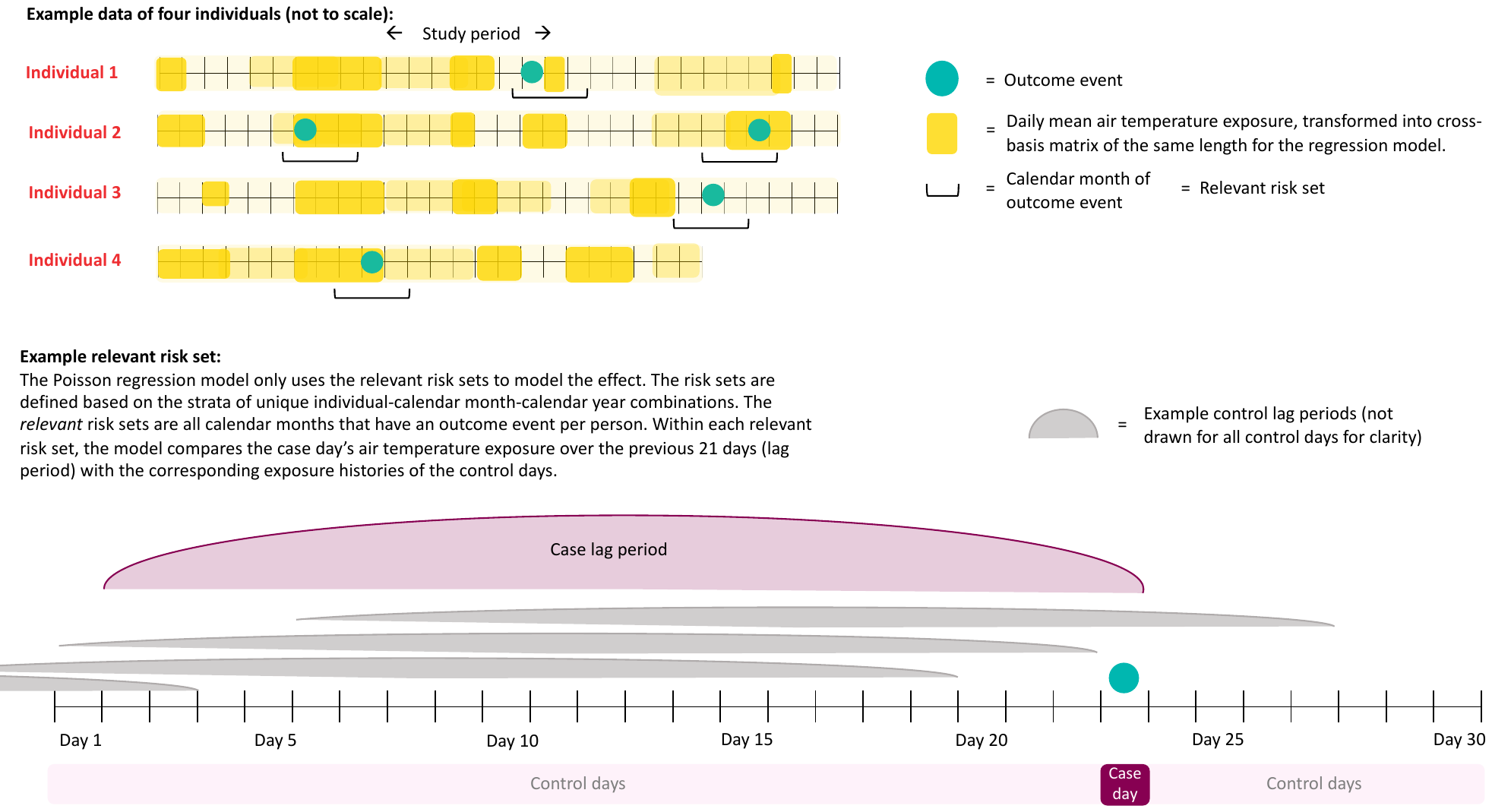

#### **Figure S1. Elaboration on regression modelling using a case time series study design.**

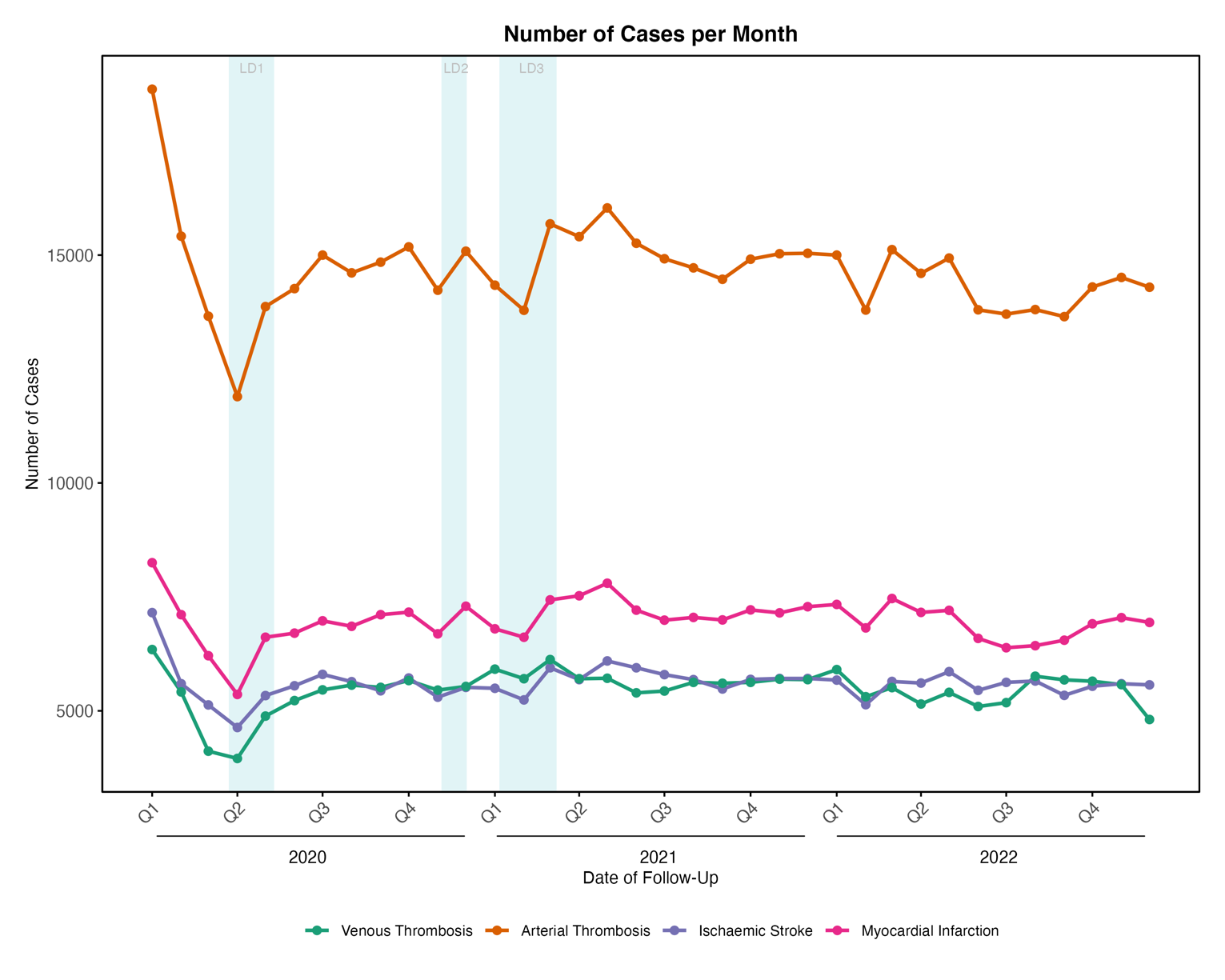

#### **Figure S2. Number of cases per month of follow-up.**

LD: lockdown period.

#### **Figure S3. Flowchart cohort derivation.**

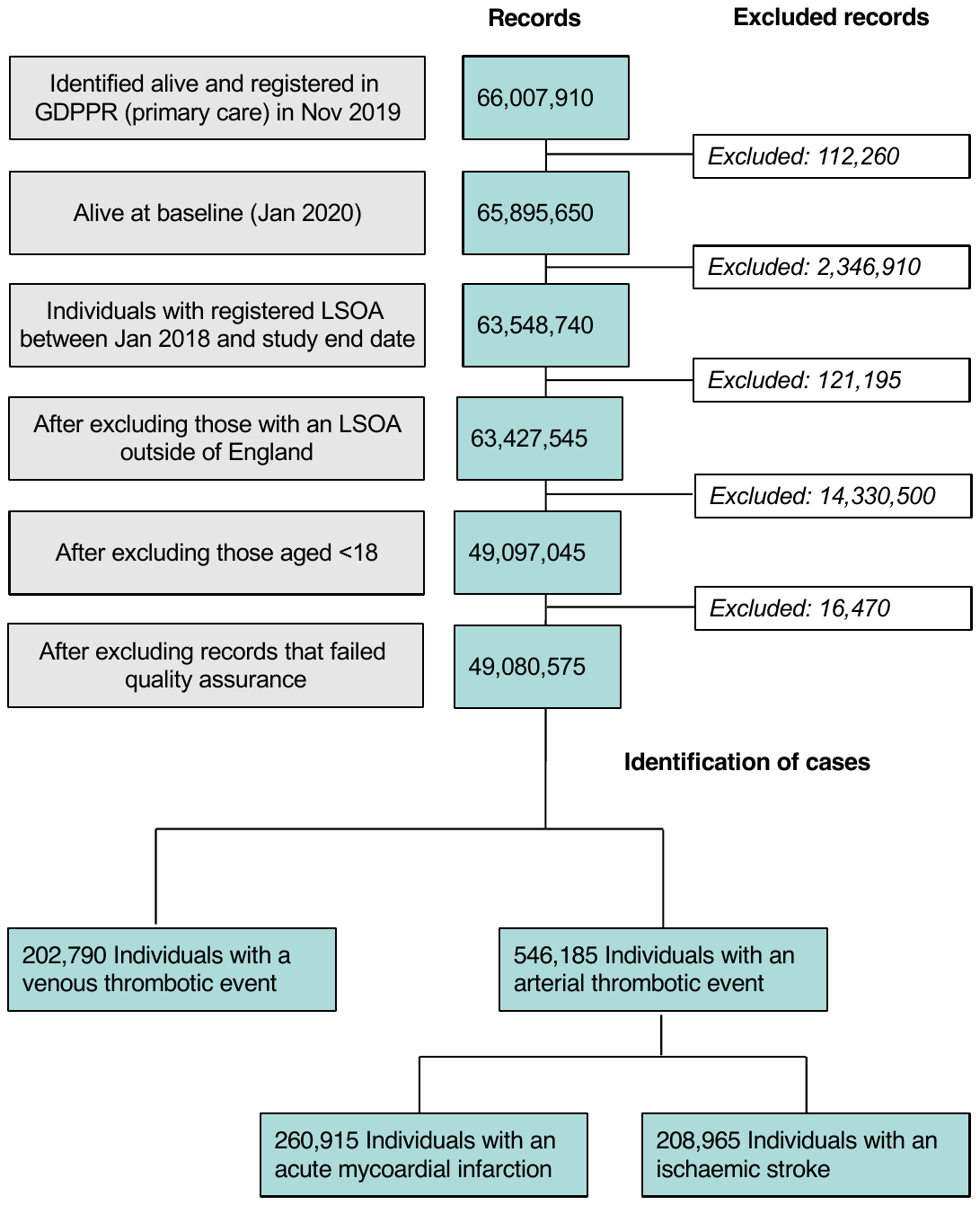

#### **Figure S4. Air pollution adjusted analysis.**

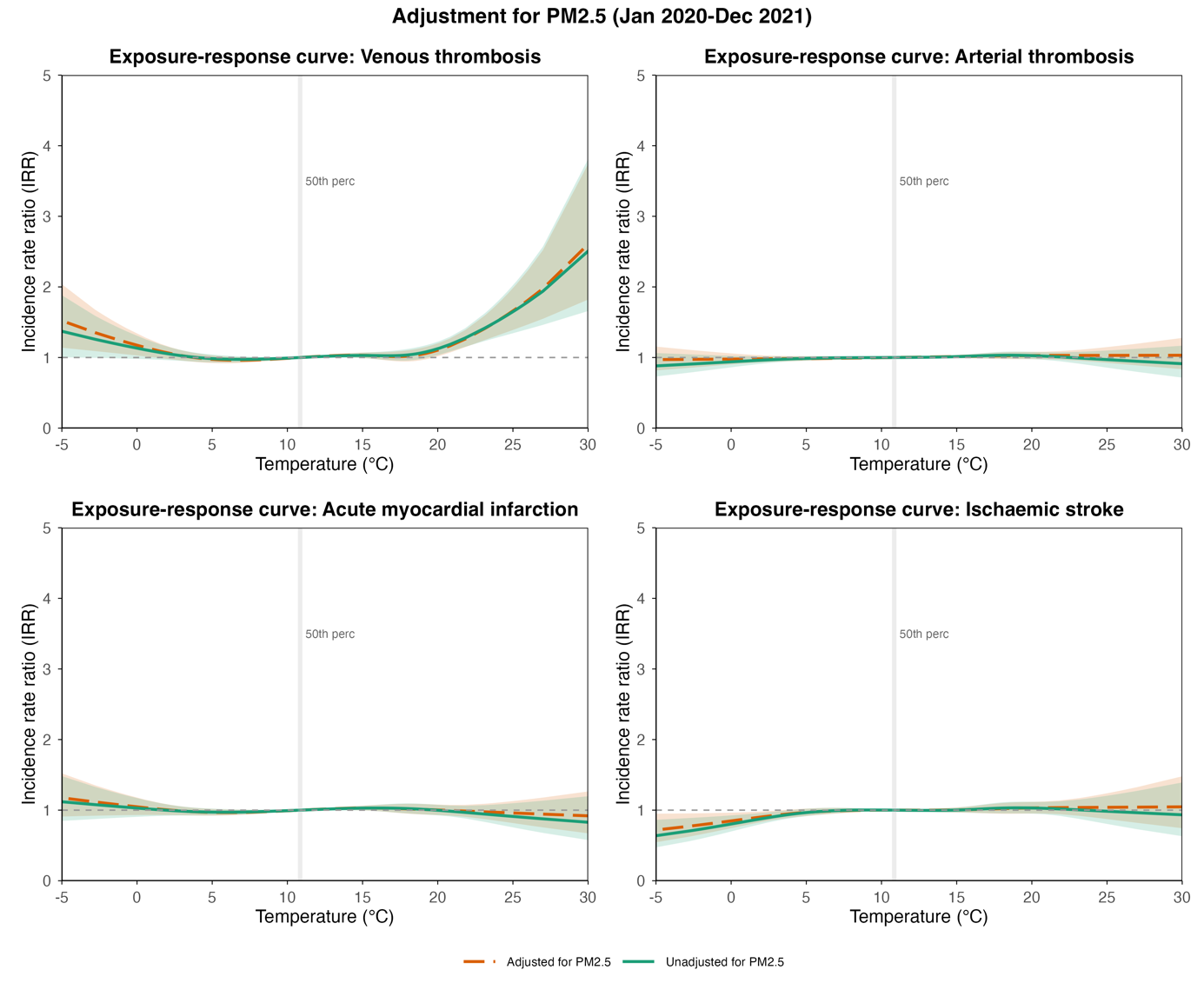

#### **Figure S5. COVID-19 diagnosis adjusted analysis.**

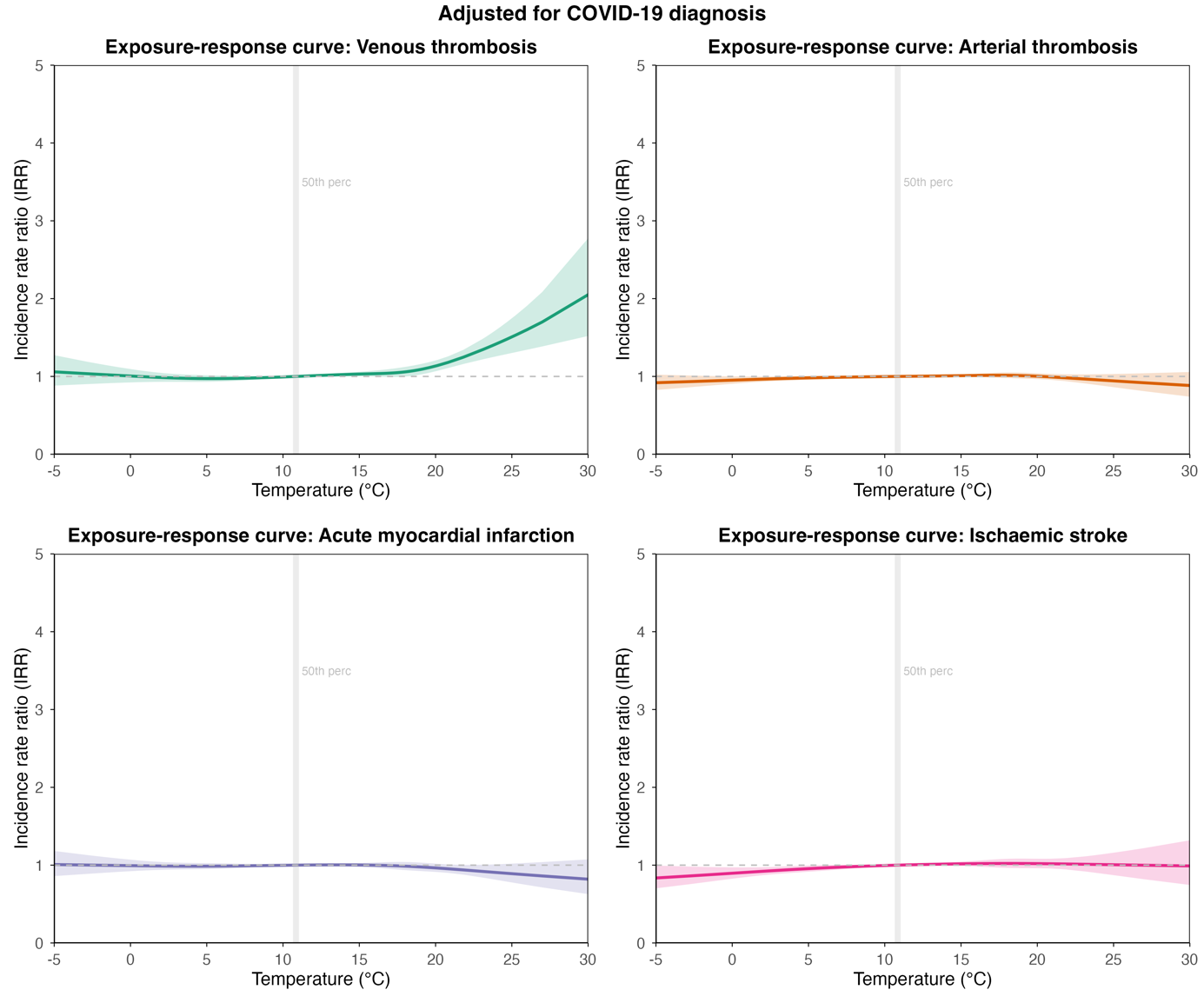

#### **Figure S6. National lockdown adjusted analysis.**

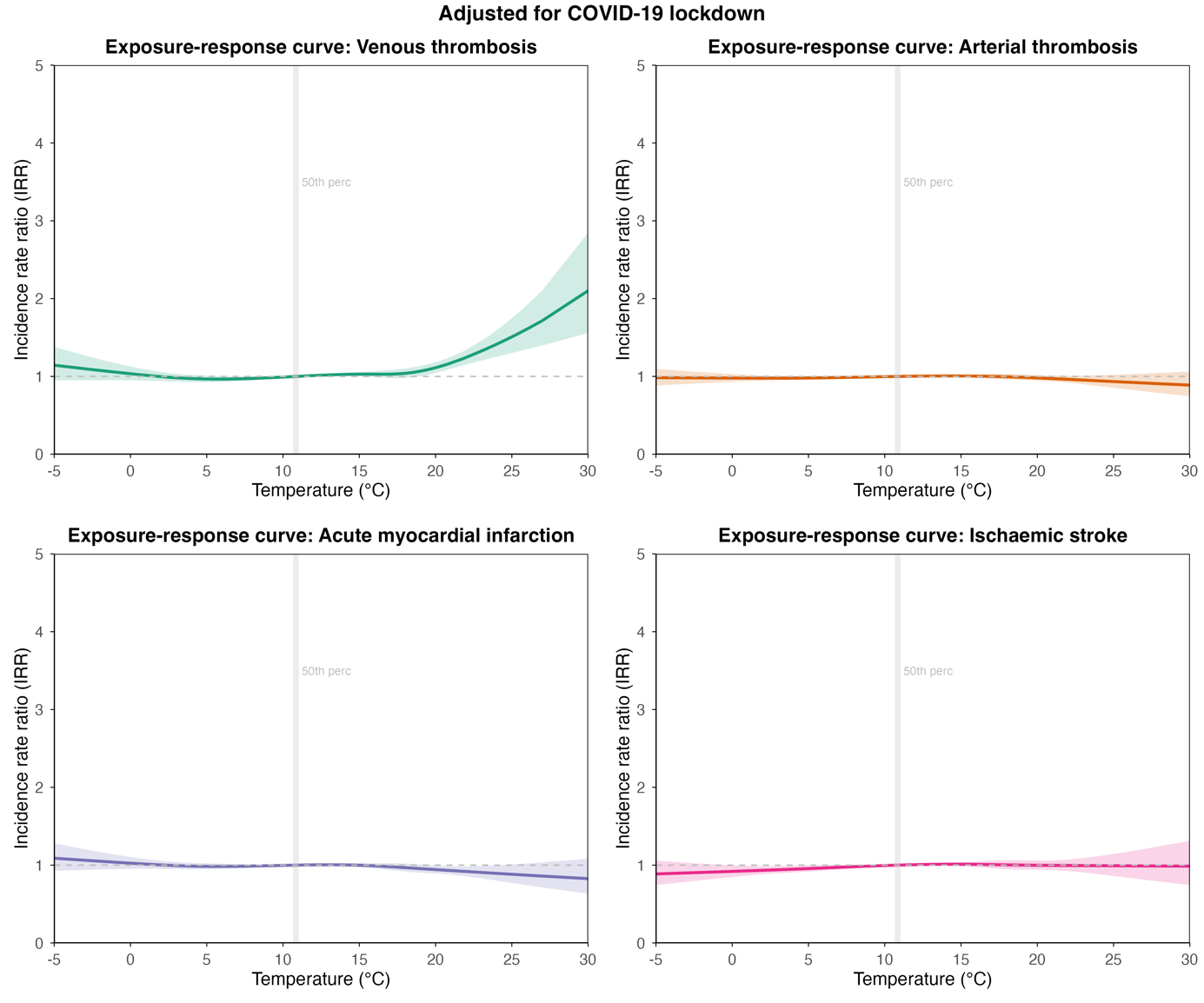

#### **Figure S7. Stratification by COVID-19 status (Diagnosis within 6 months before first event versus no diagnosis within 6 months before first event).**

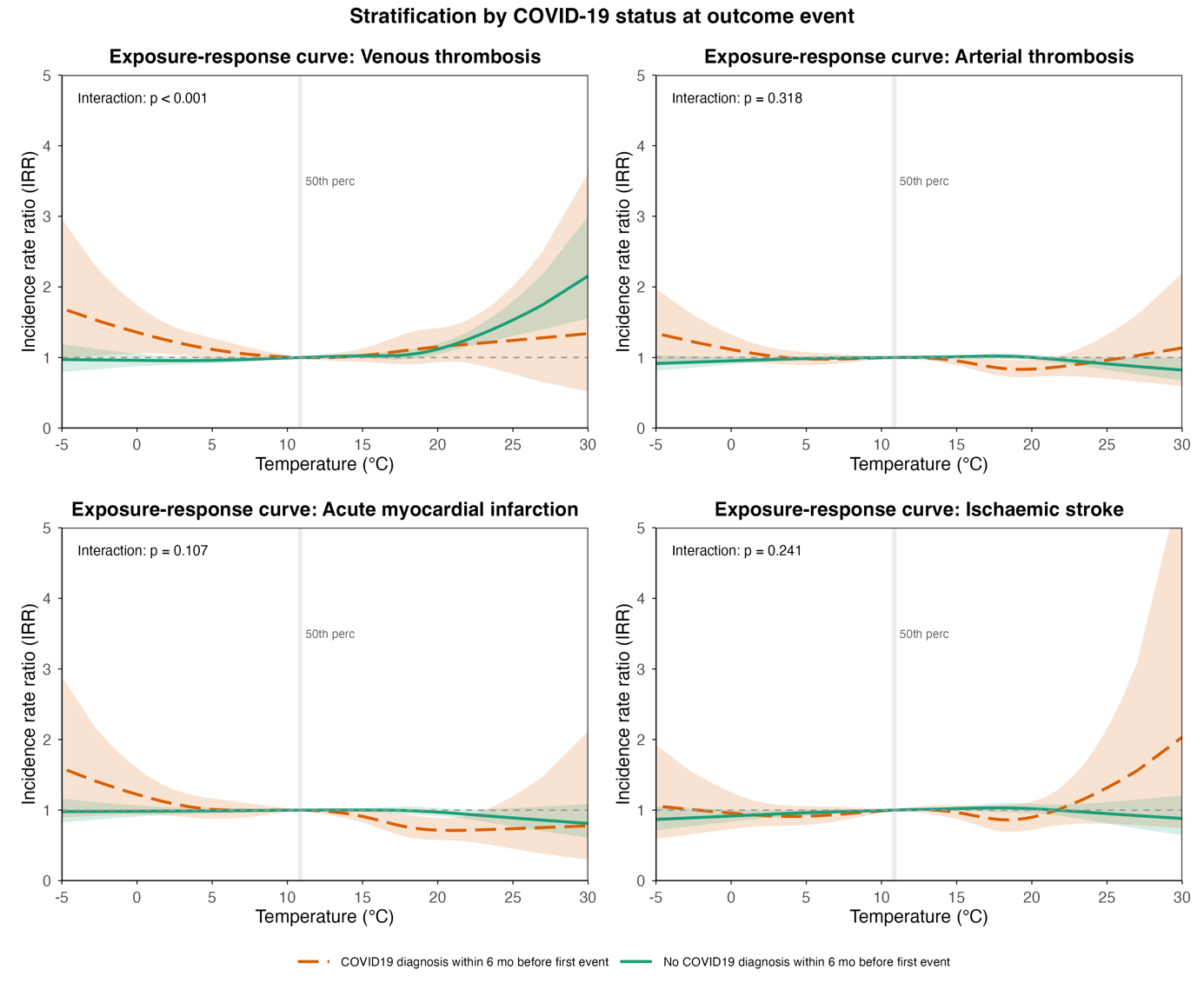

#### **Figure S8. Stratification by sex.**

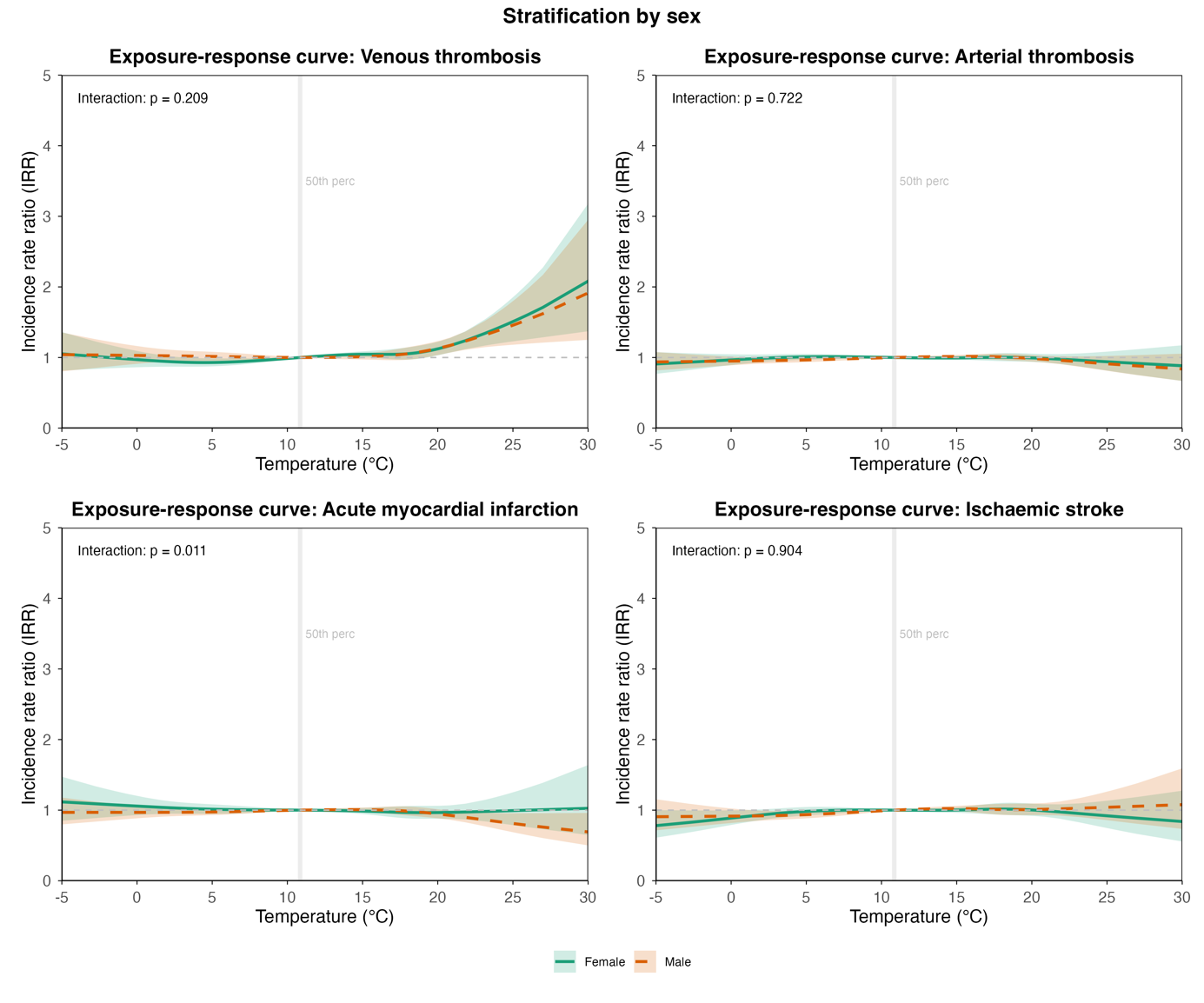

#### **Figure S9. Stratification by age group.**

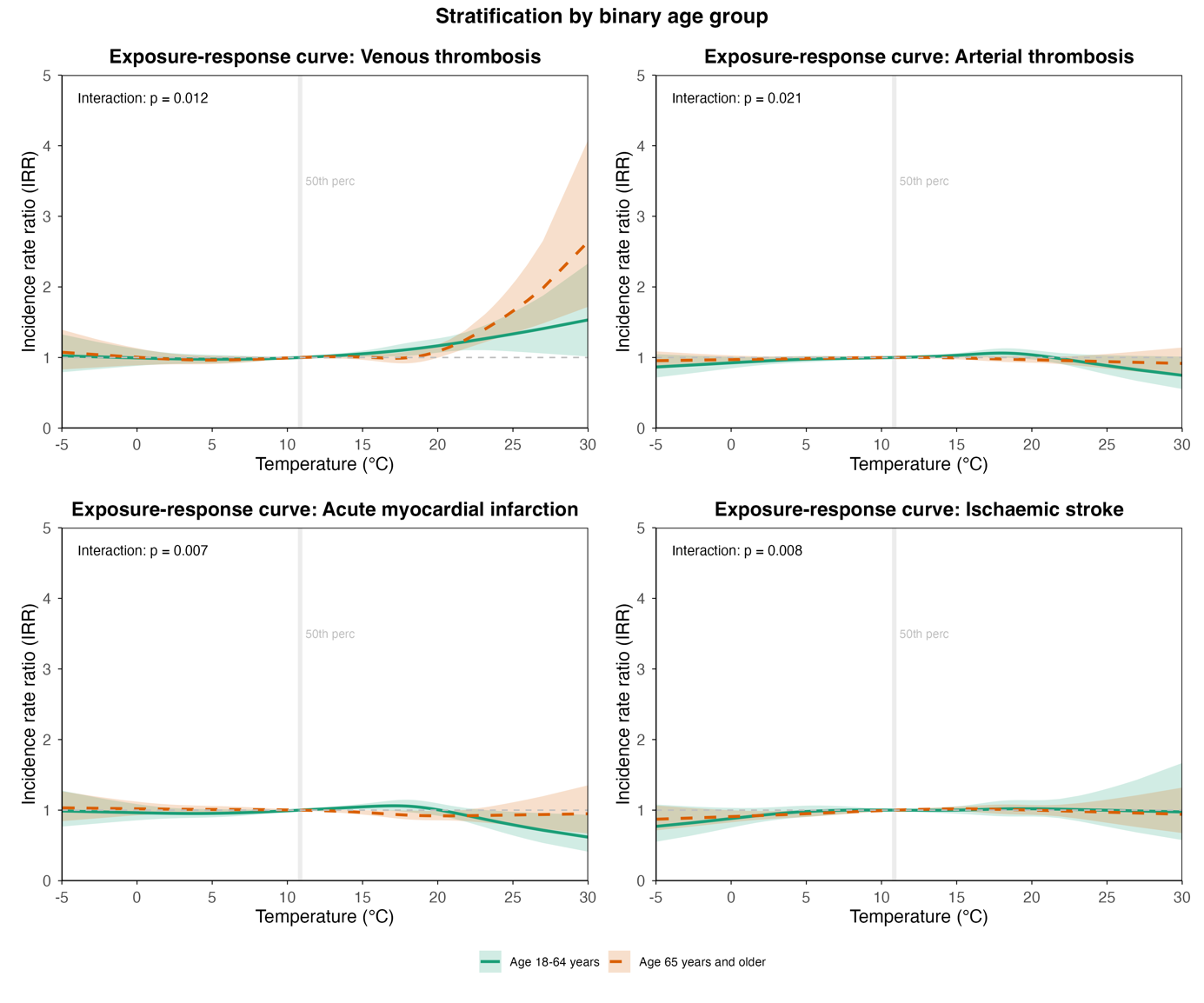

#### **Figure S10. Stratification by ethnic group.**

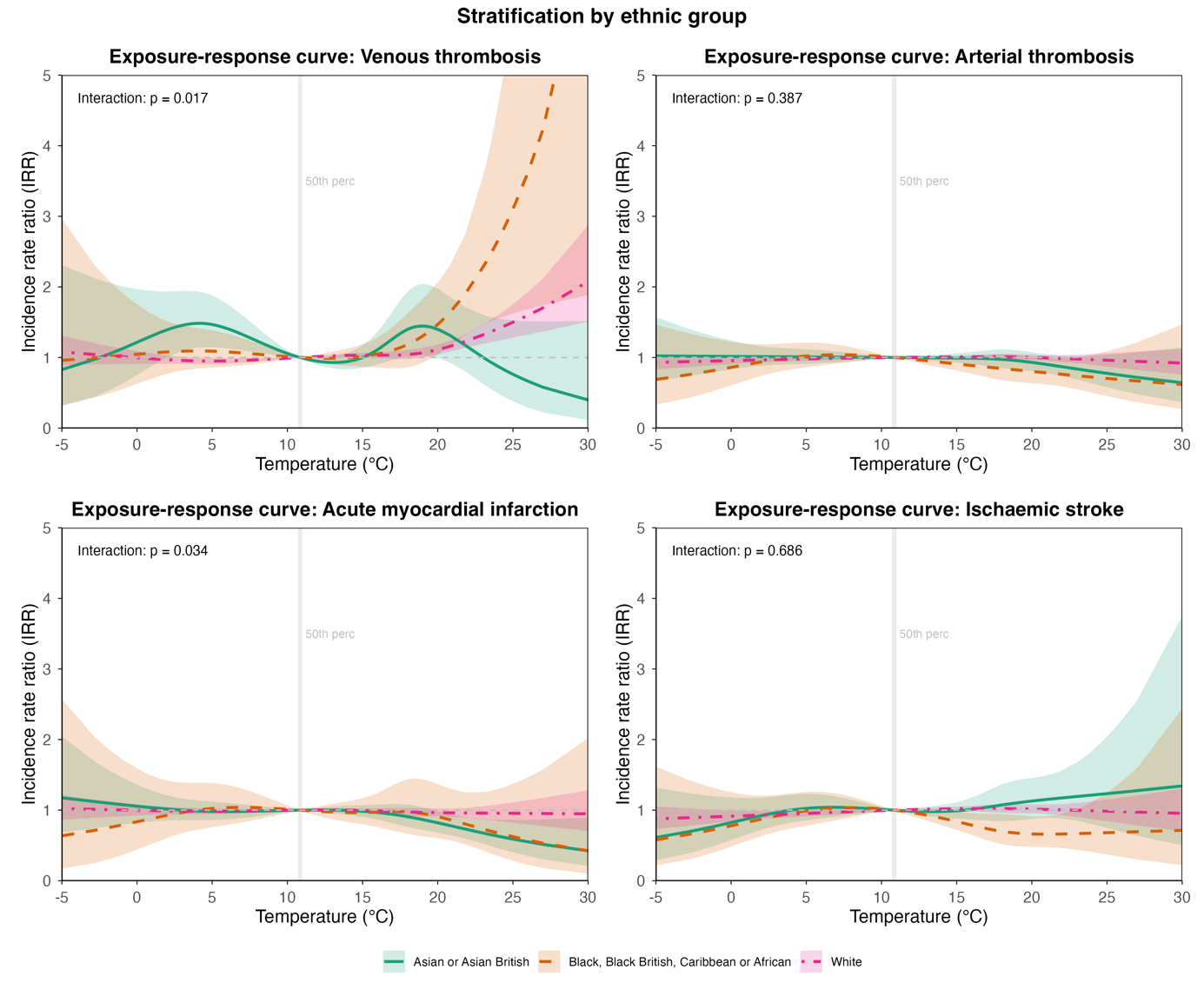

#### **Figure S11. Stratification by rural/urban area.**

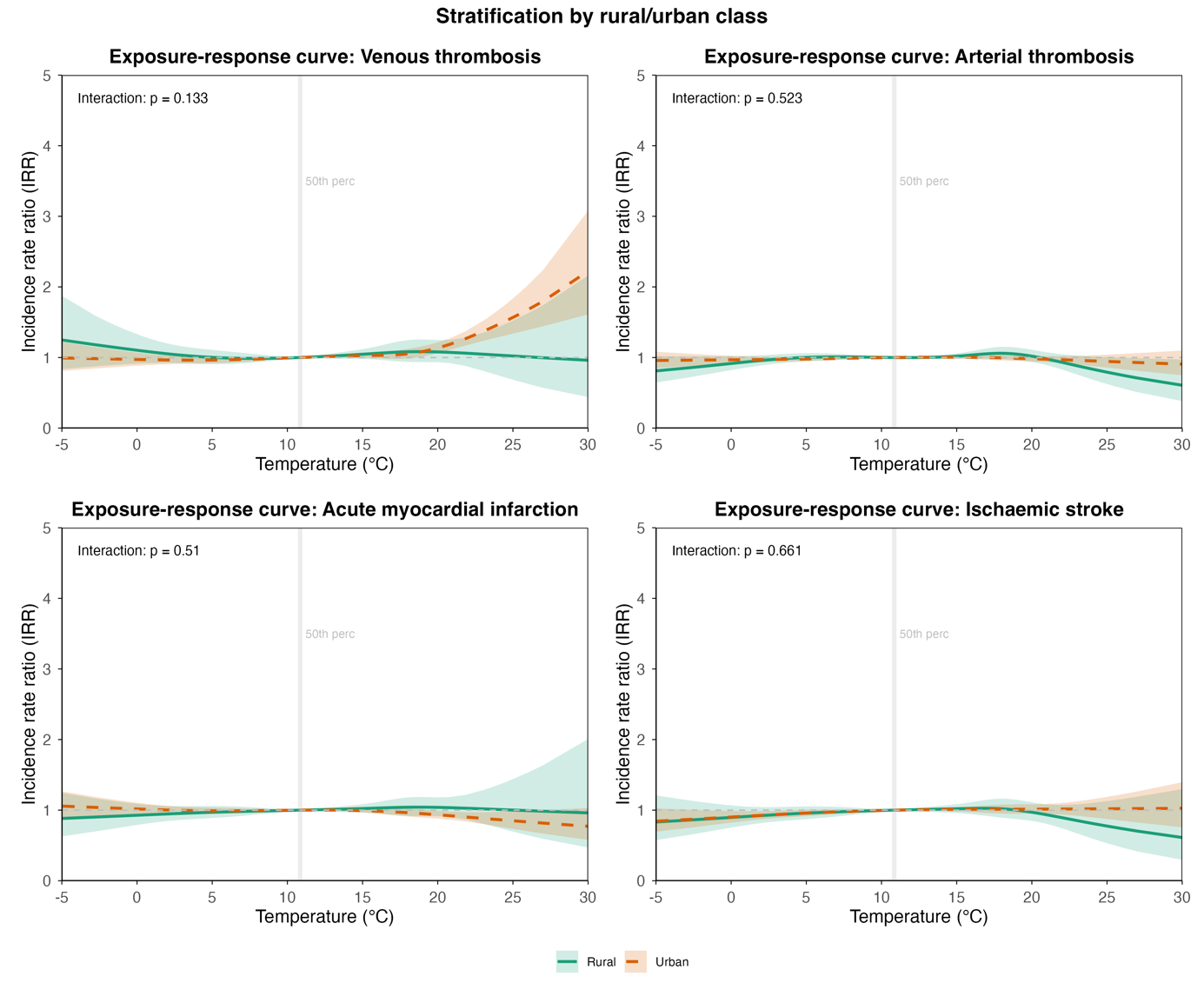

#### **Figure S12. Stratification by deprivation index quintiles.**

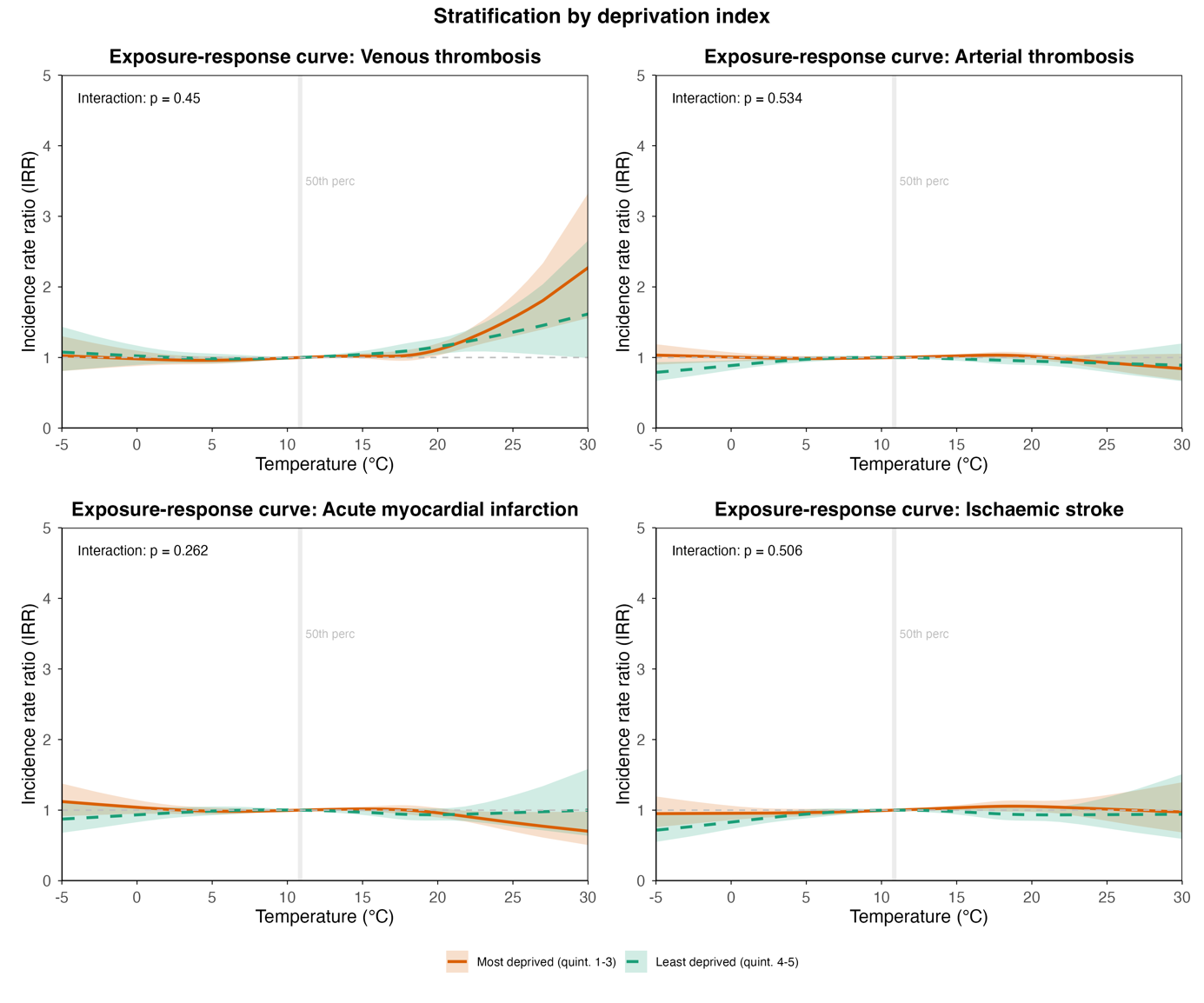

#### **Figure S13. Stratification by pre-existing health conditions.**

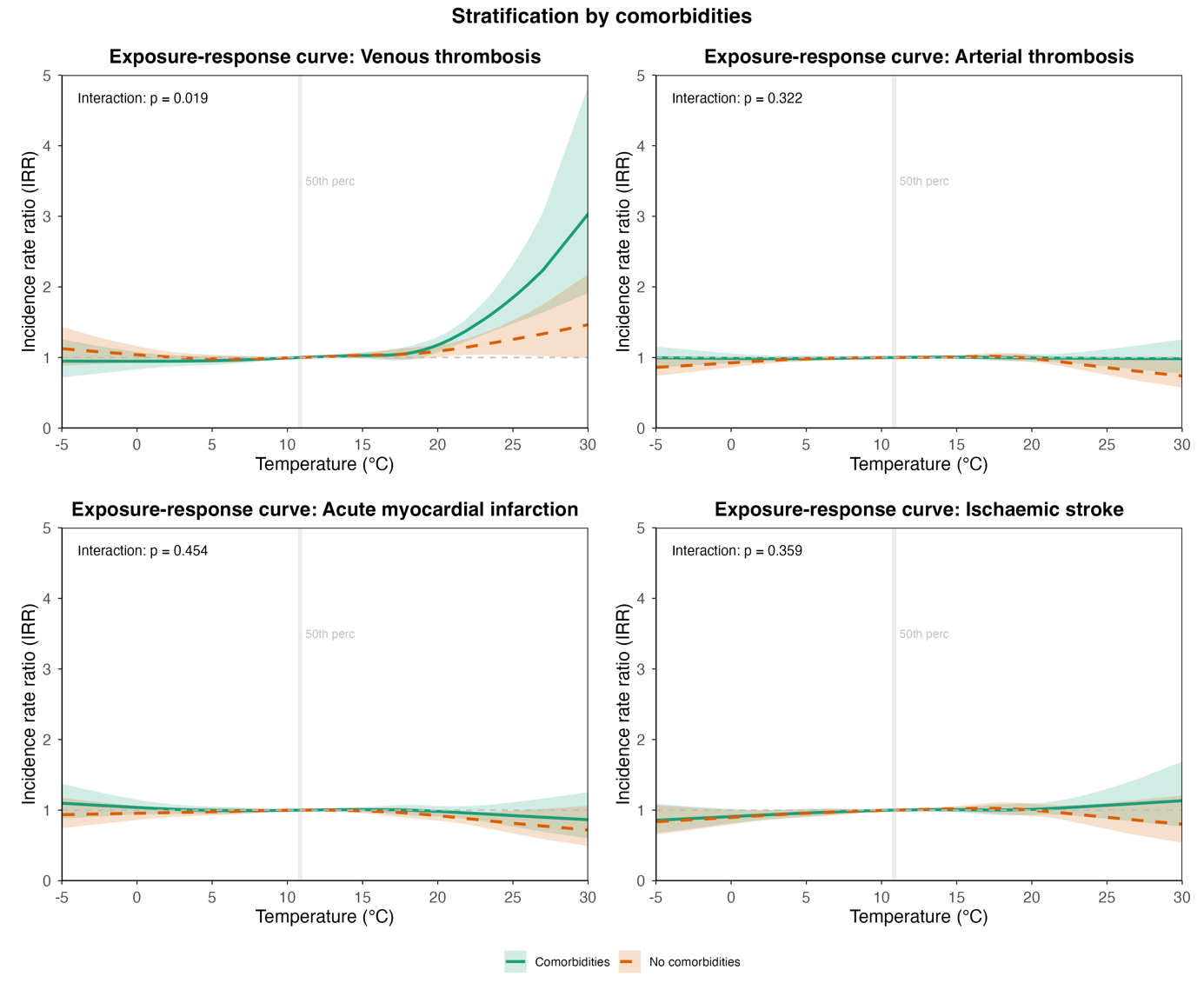

#### **Figure S14. Stratification by excess weight status.**

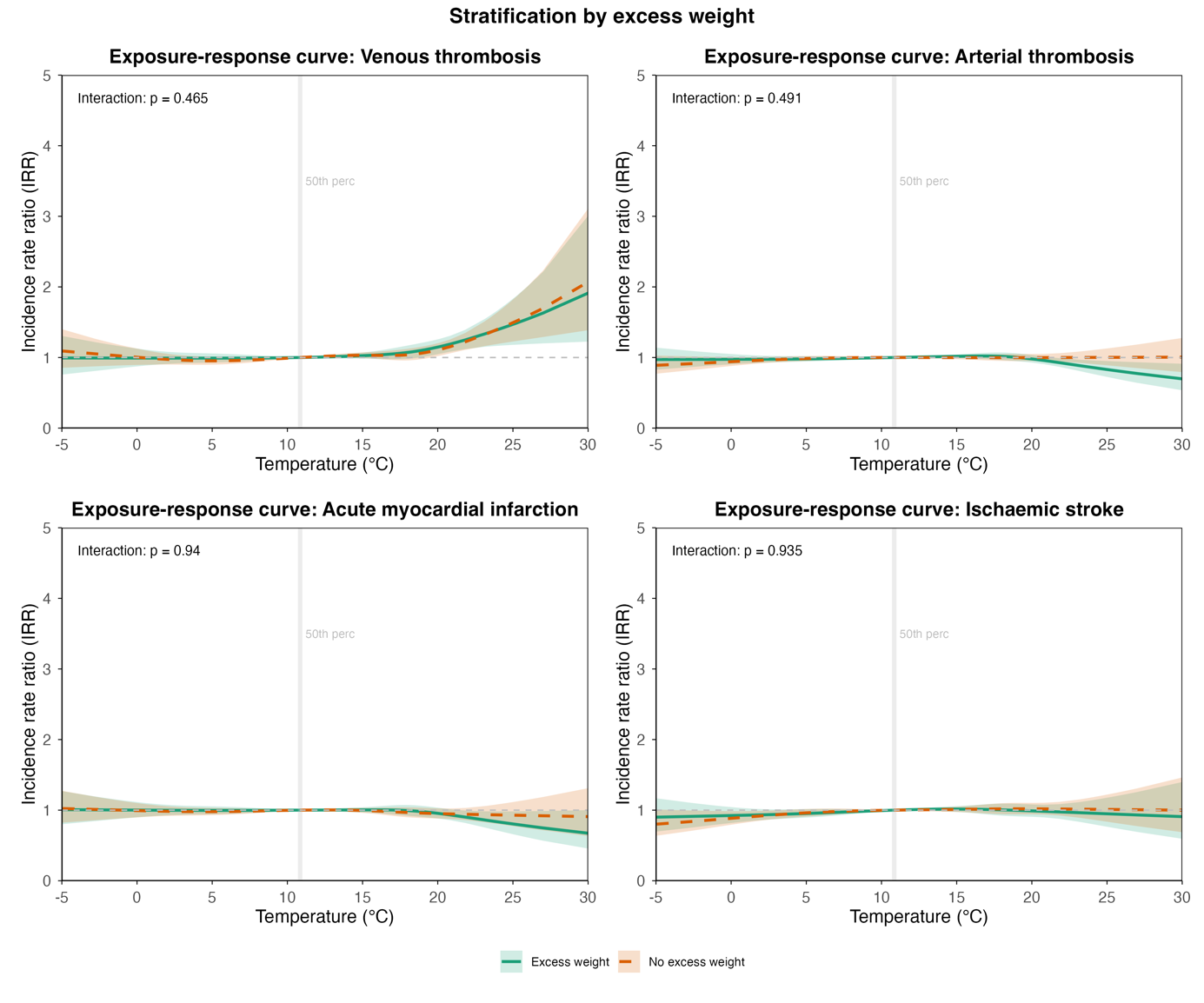

#### **Figure S15. Stratification by smoking status.**

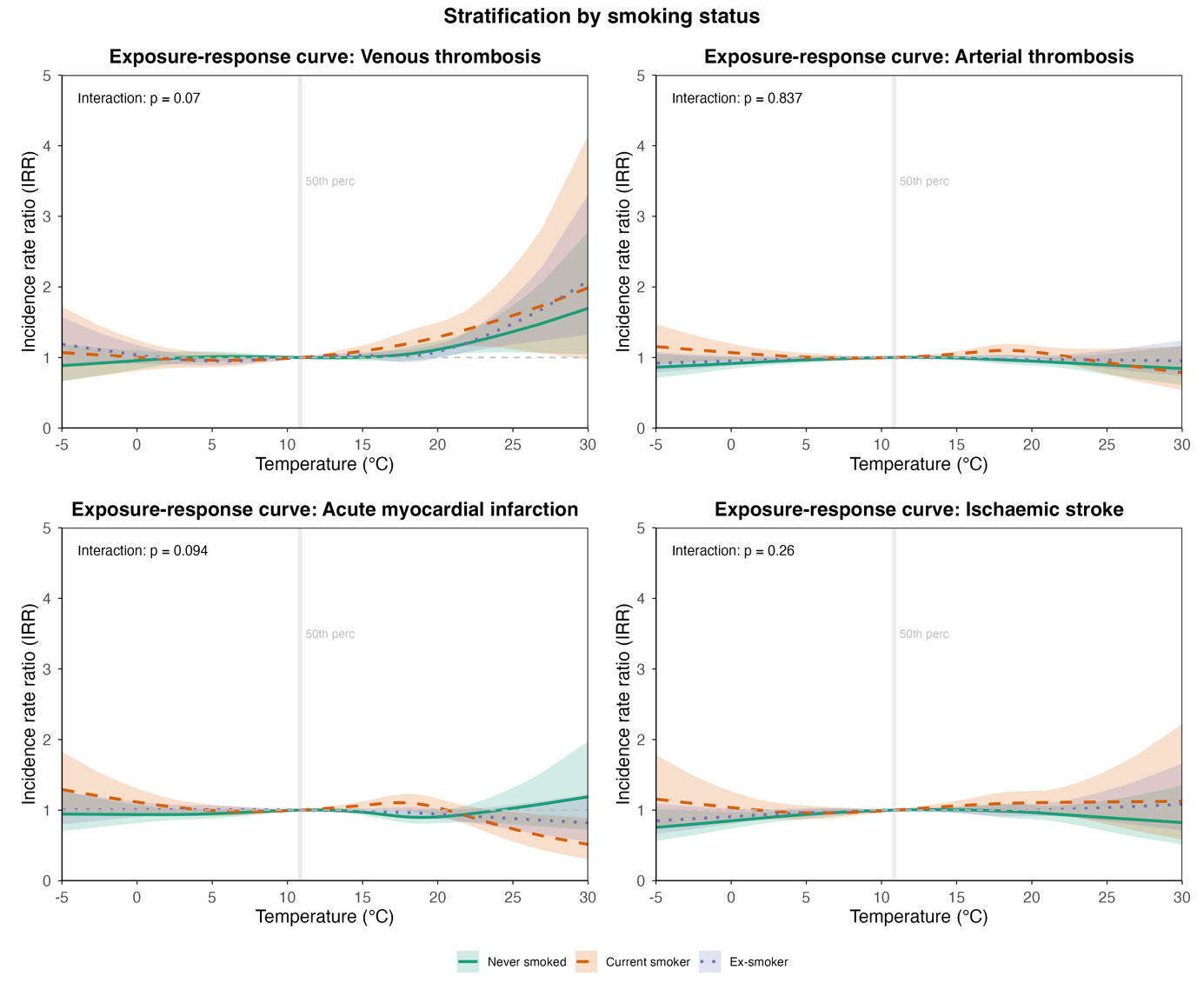

#### **Figure S16. Stratification by region venous thrombotic event.**

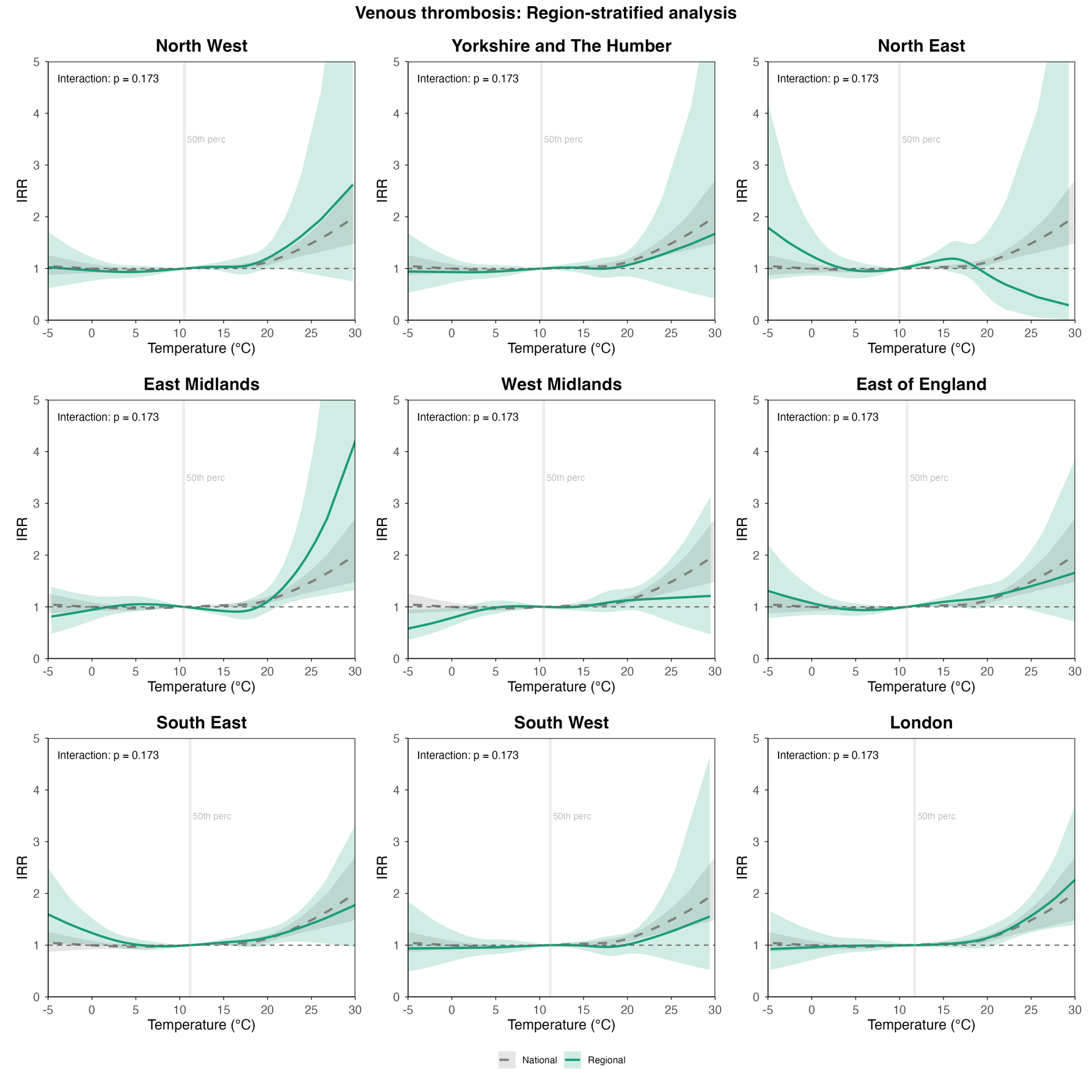

#### **Figure S17. Stratification by region arterial thrombotic event.**

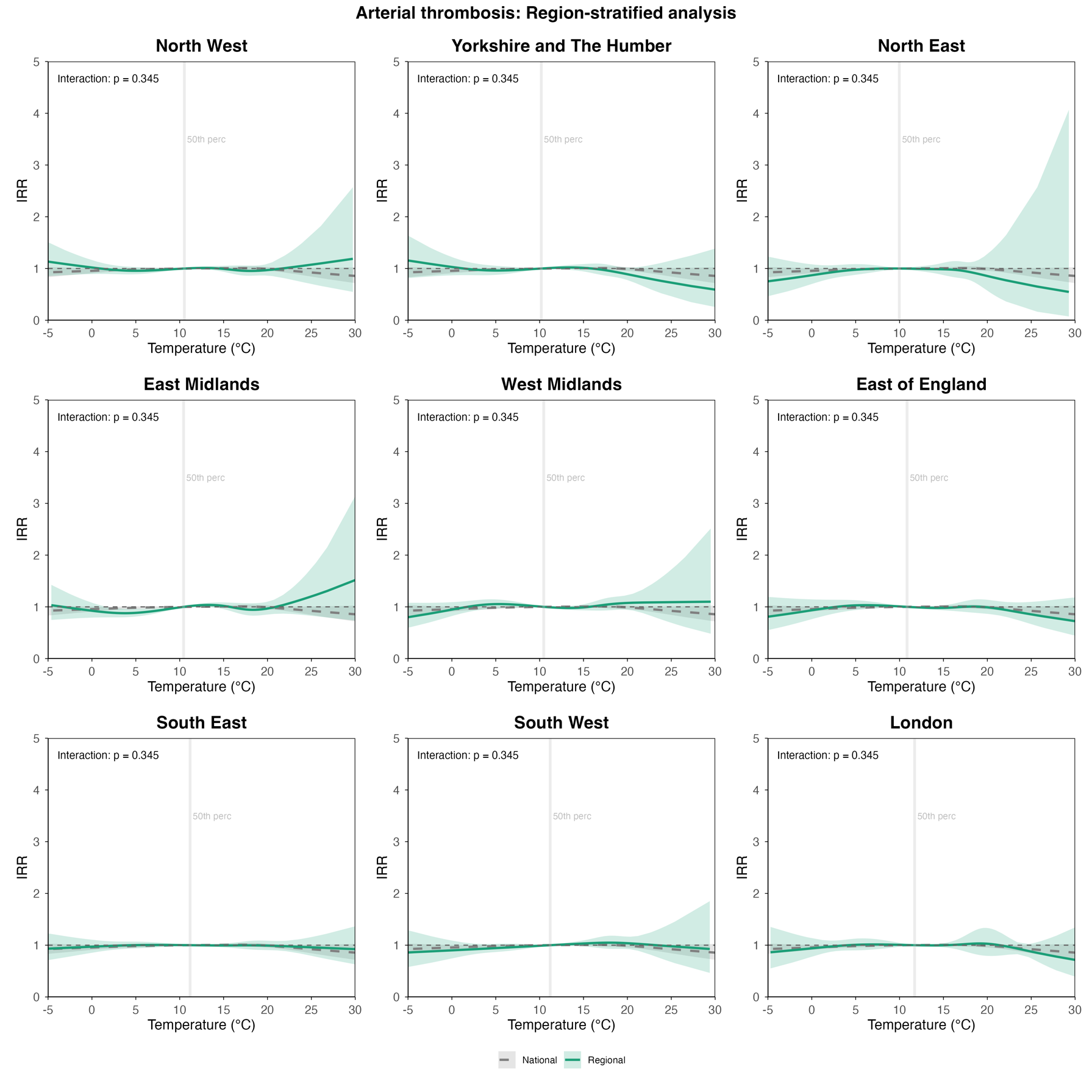

#### **Figure S18. Stratification by region acute myocardial infarction.**

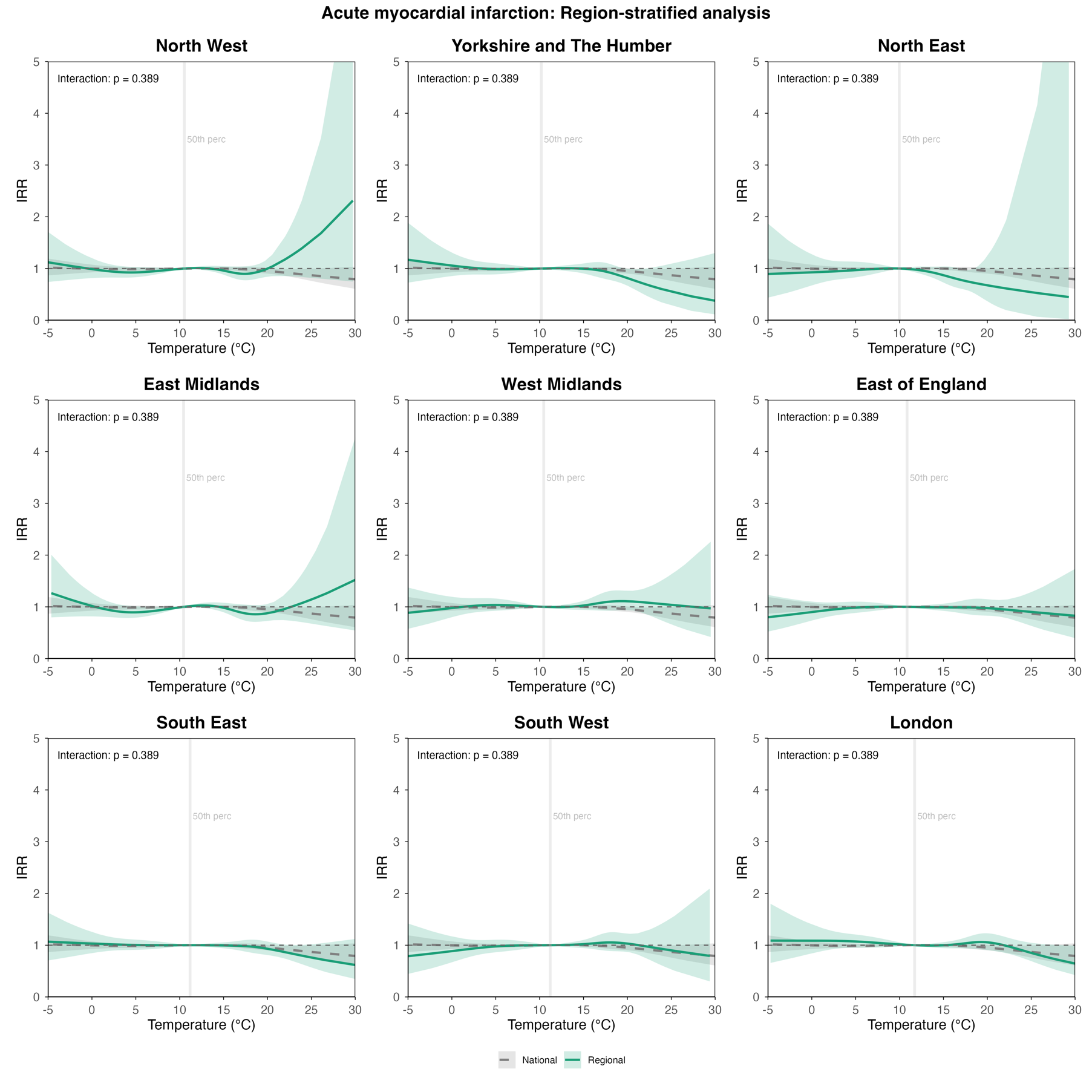

#### **Figure S19. Stratification by region ischaemic stroke.**

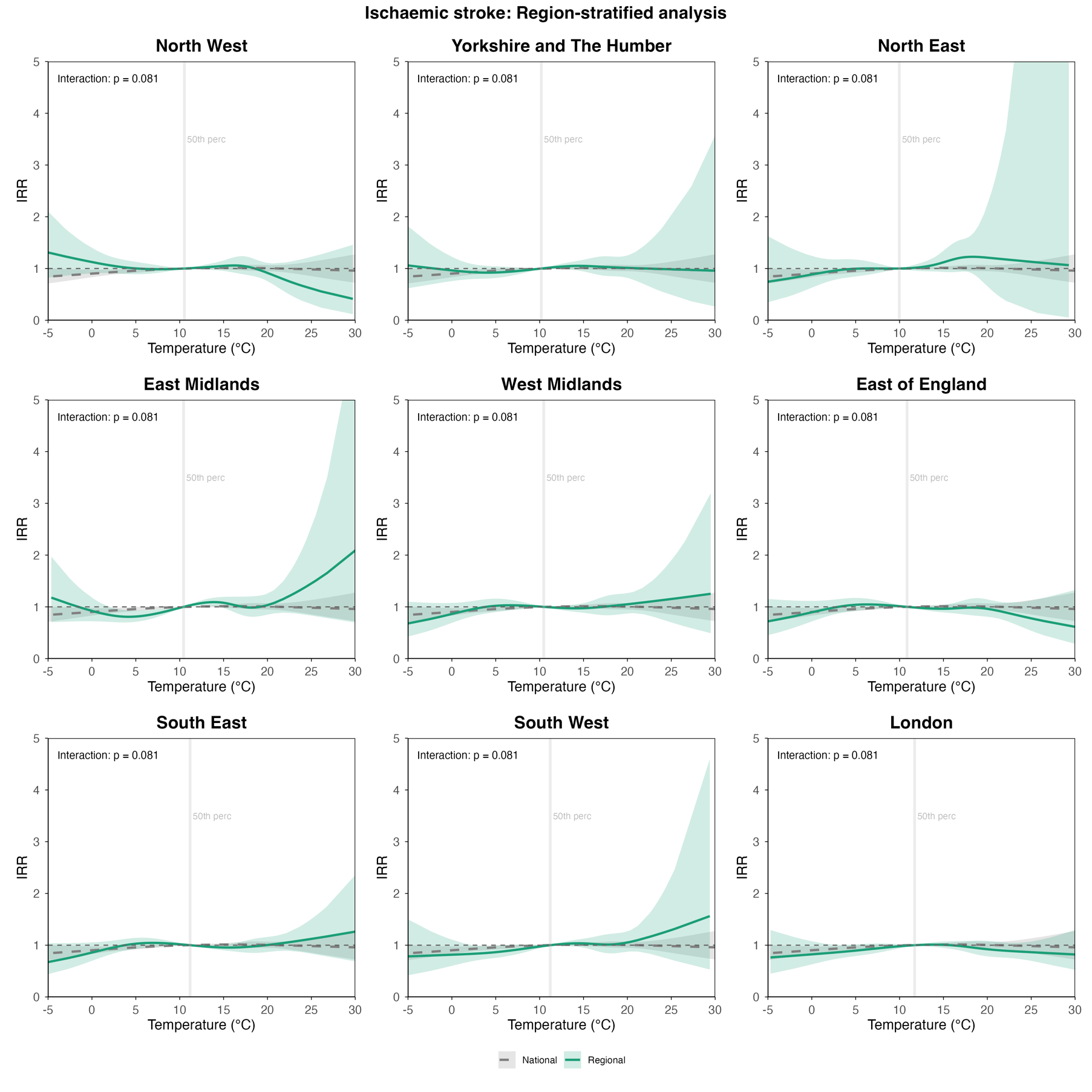

#### **Figure S20. Heat map of air temperature impact analysis across regions for venous thrombotic events.**

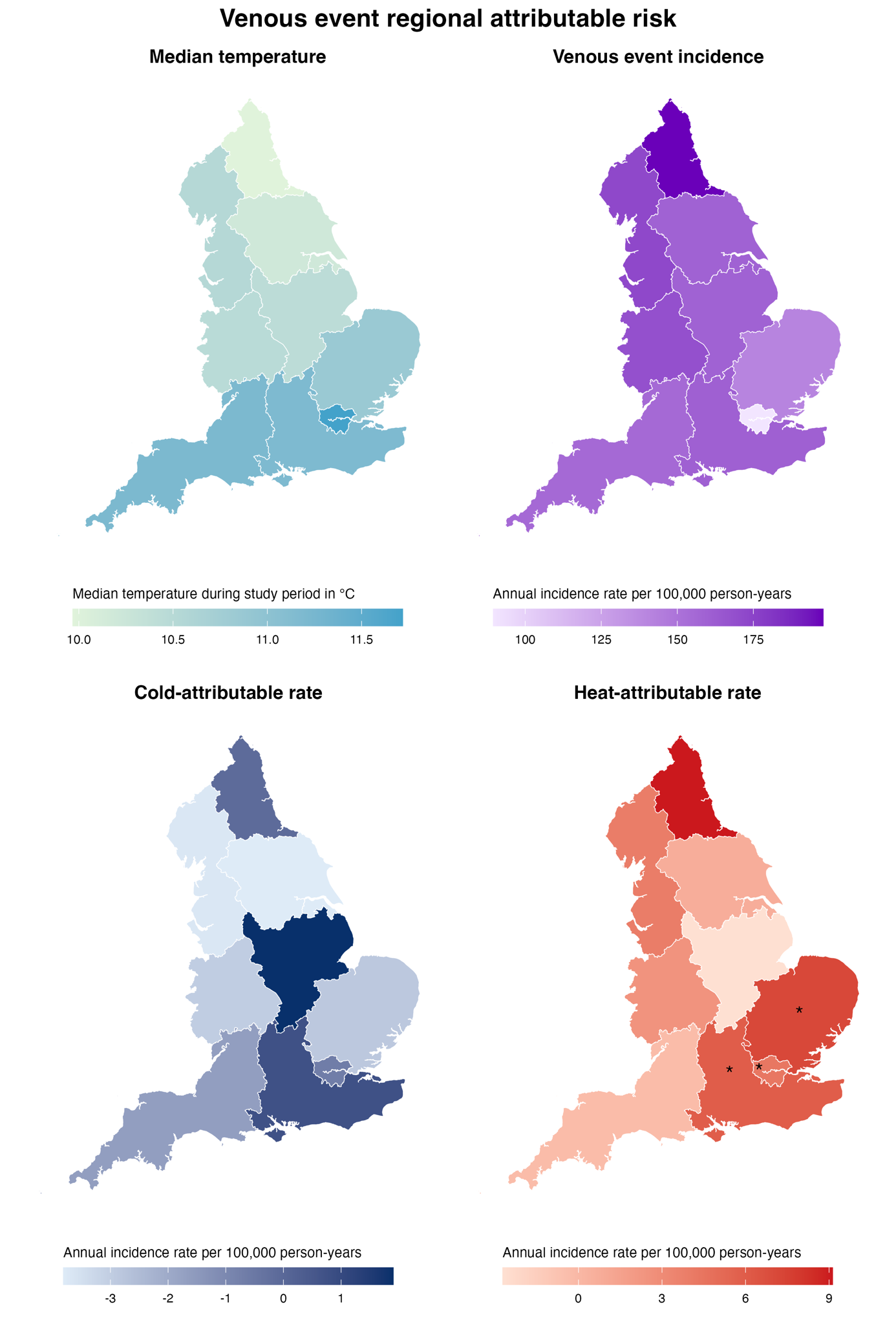

#### **Figure S21. Restricted study period sensitivity analysis.**

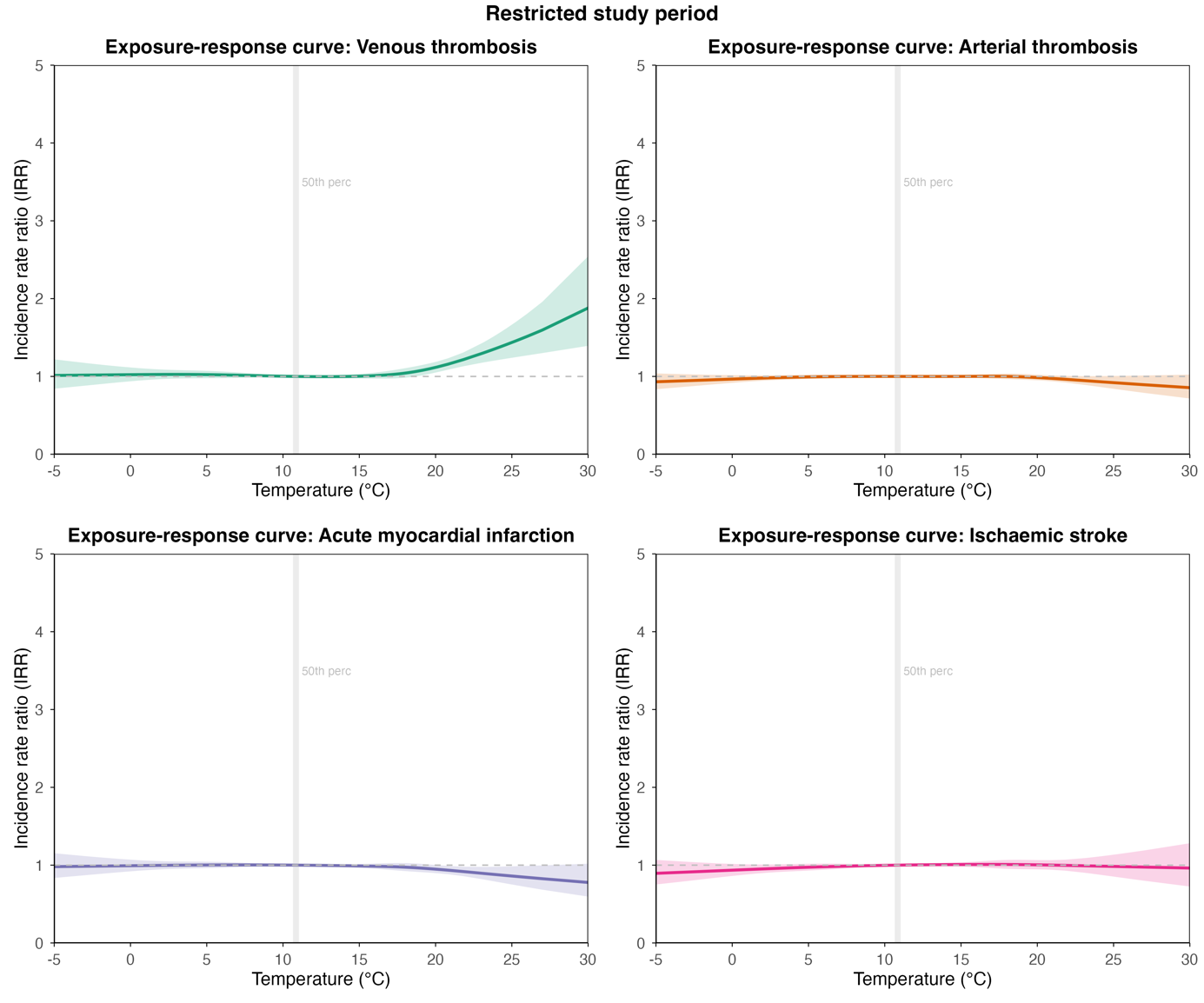

#### **Figure S22. First events only sensitivity analysis.**

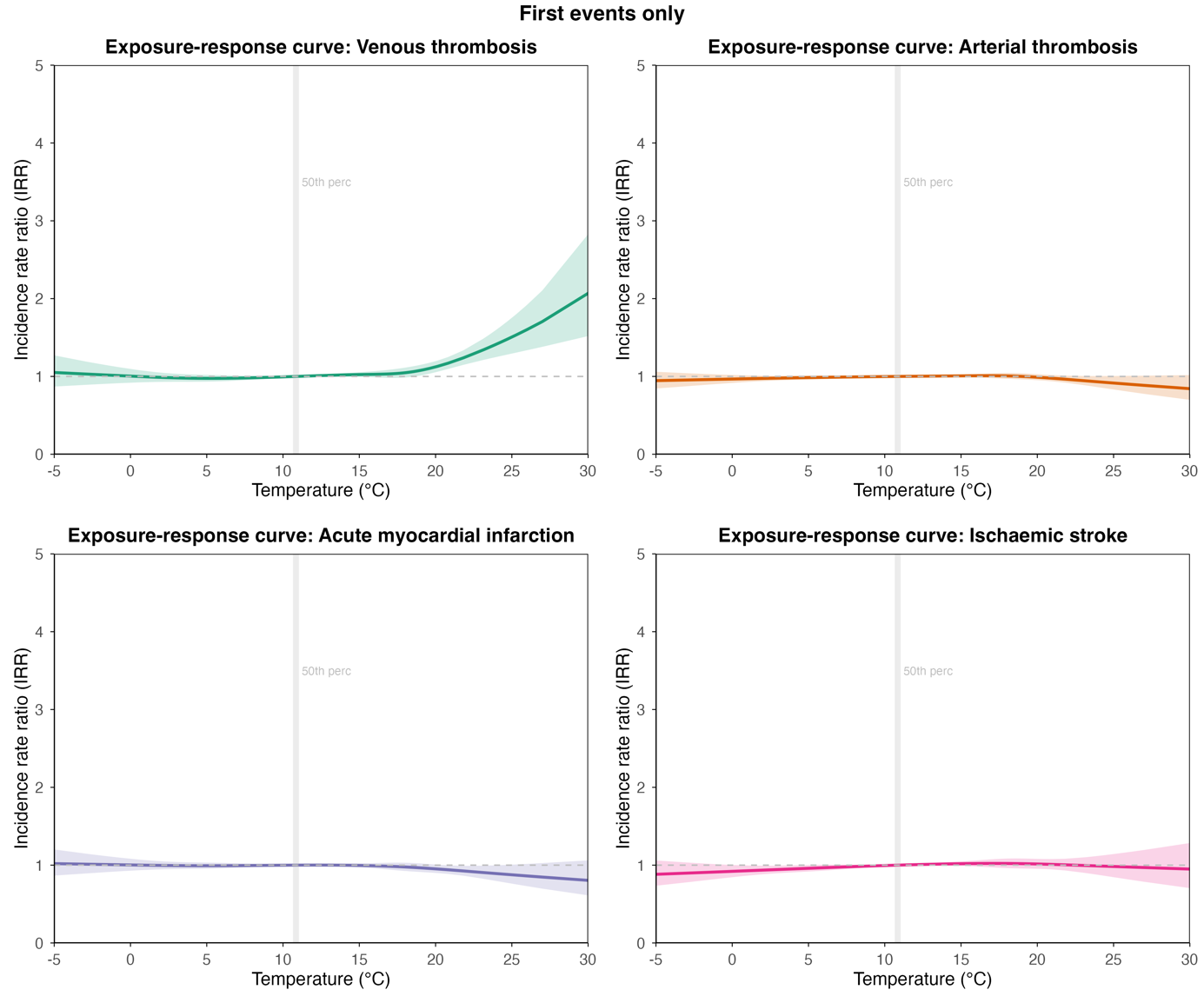

#### **Figure S23. Fatal vs non-fatal cases (death within 7 days definition).**

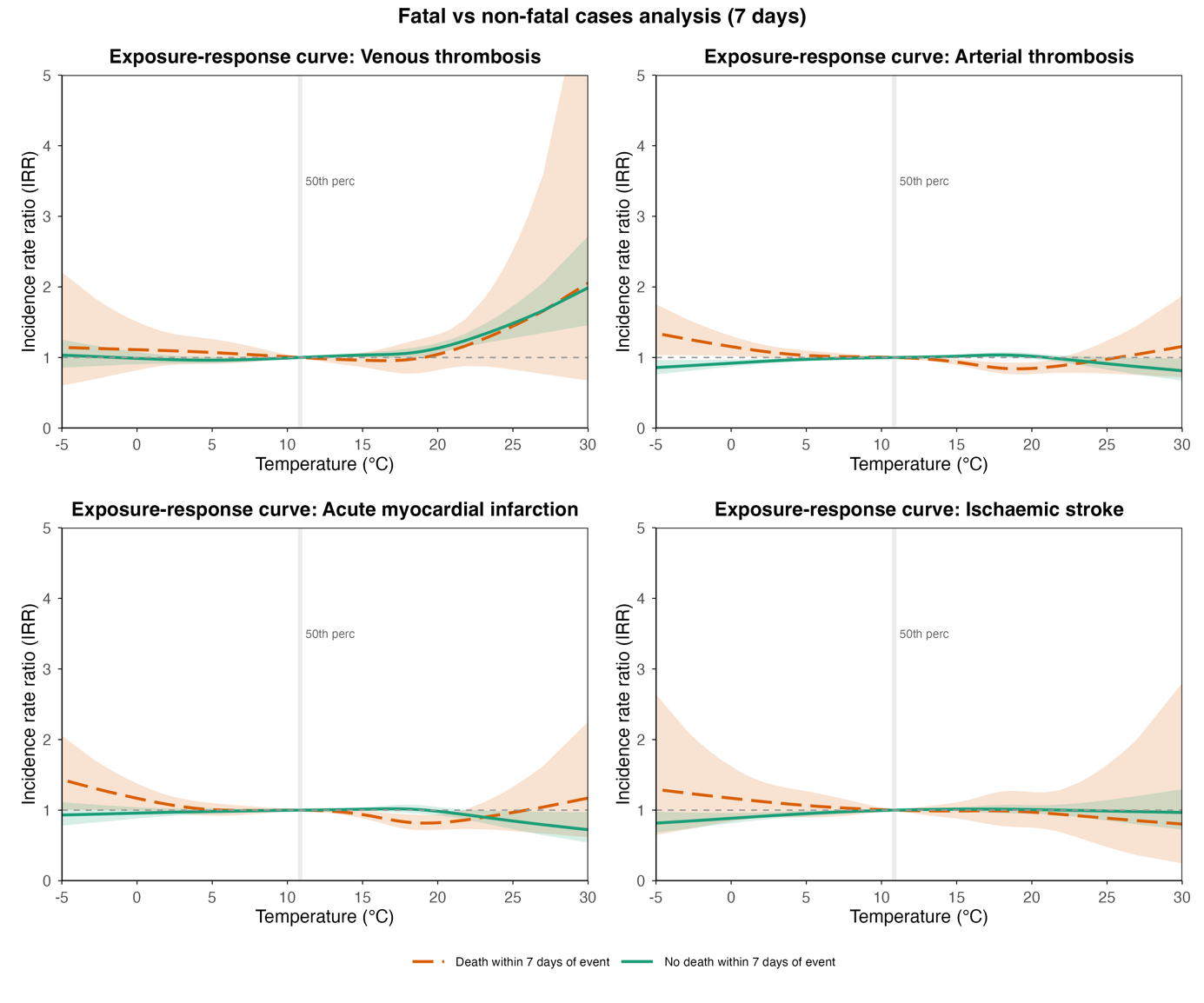

#### **Figure S24. Fatal vs non-fatal cases (death within 14 days definition).**

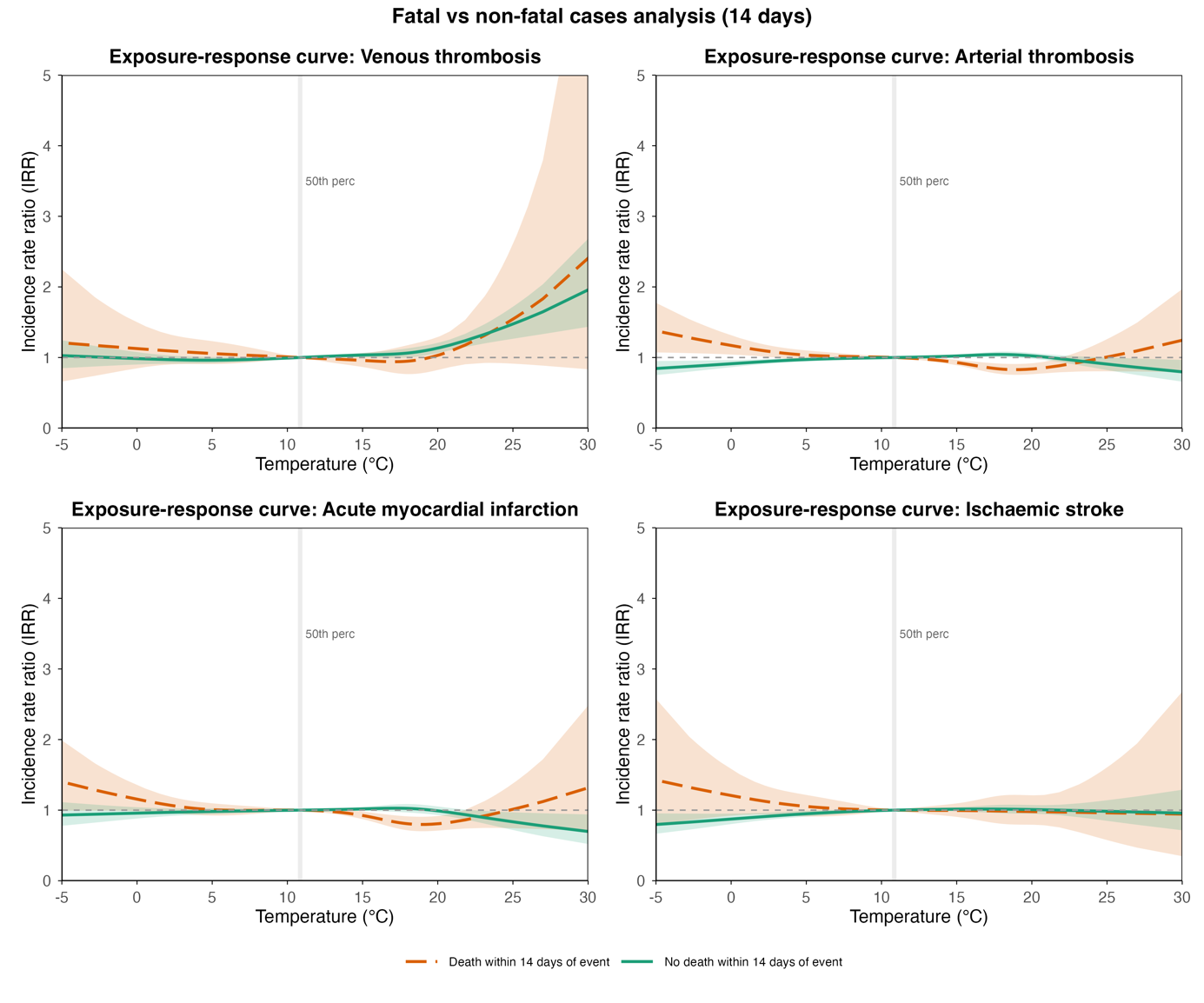

#### **Figure S25. Fatal vs non-fatal cases (death within 30 days definition).**

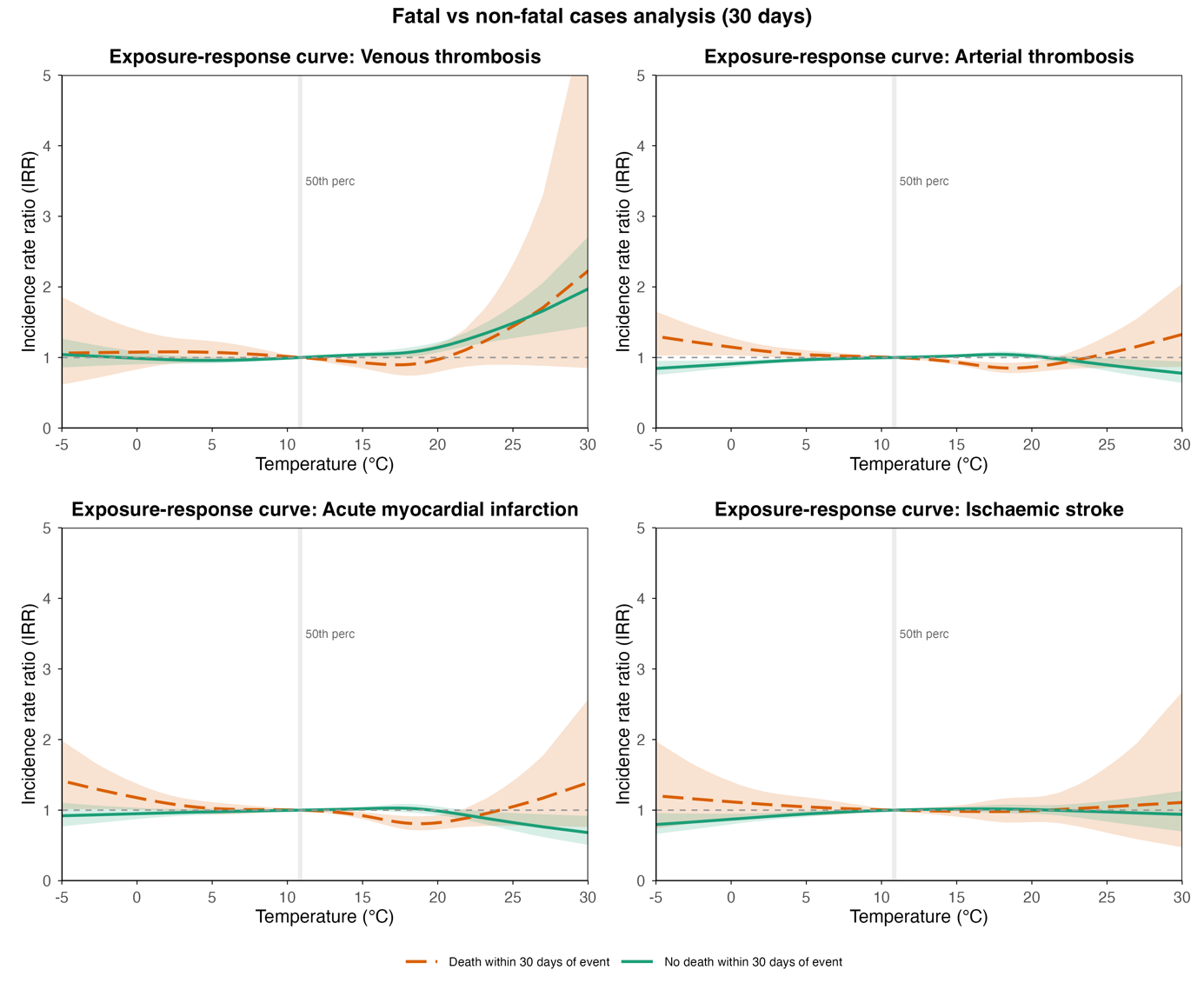

#### **Figure S26. Censoring at actual date of death analysis.**

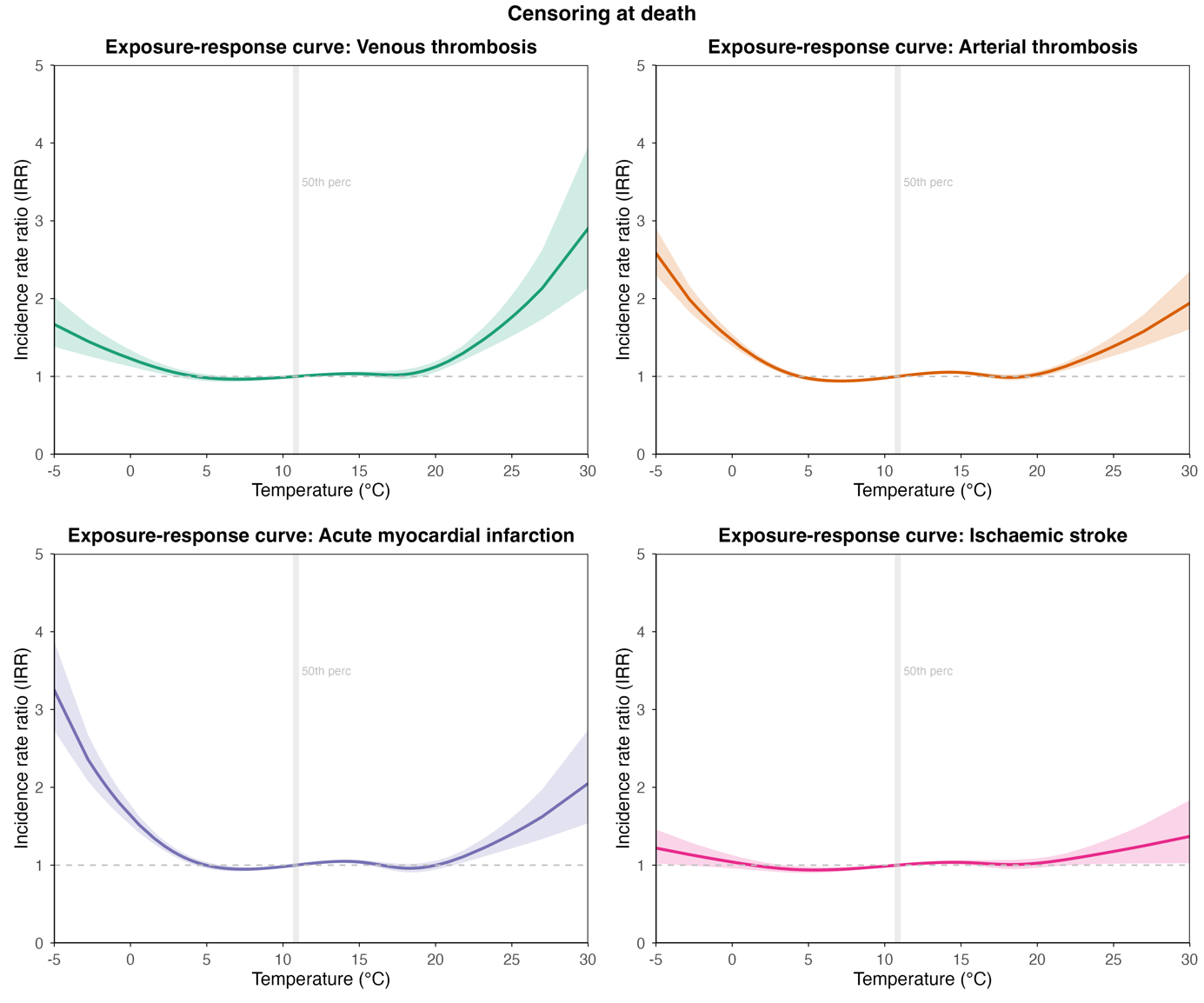

#### **Figure S27. Cumulative IRR over 4-day lag period.**

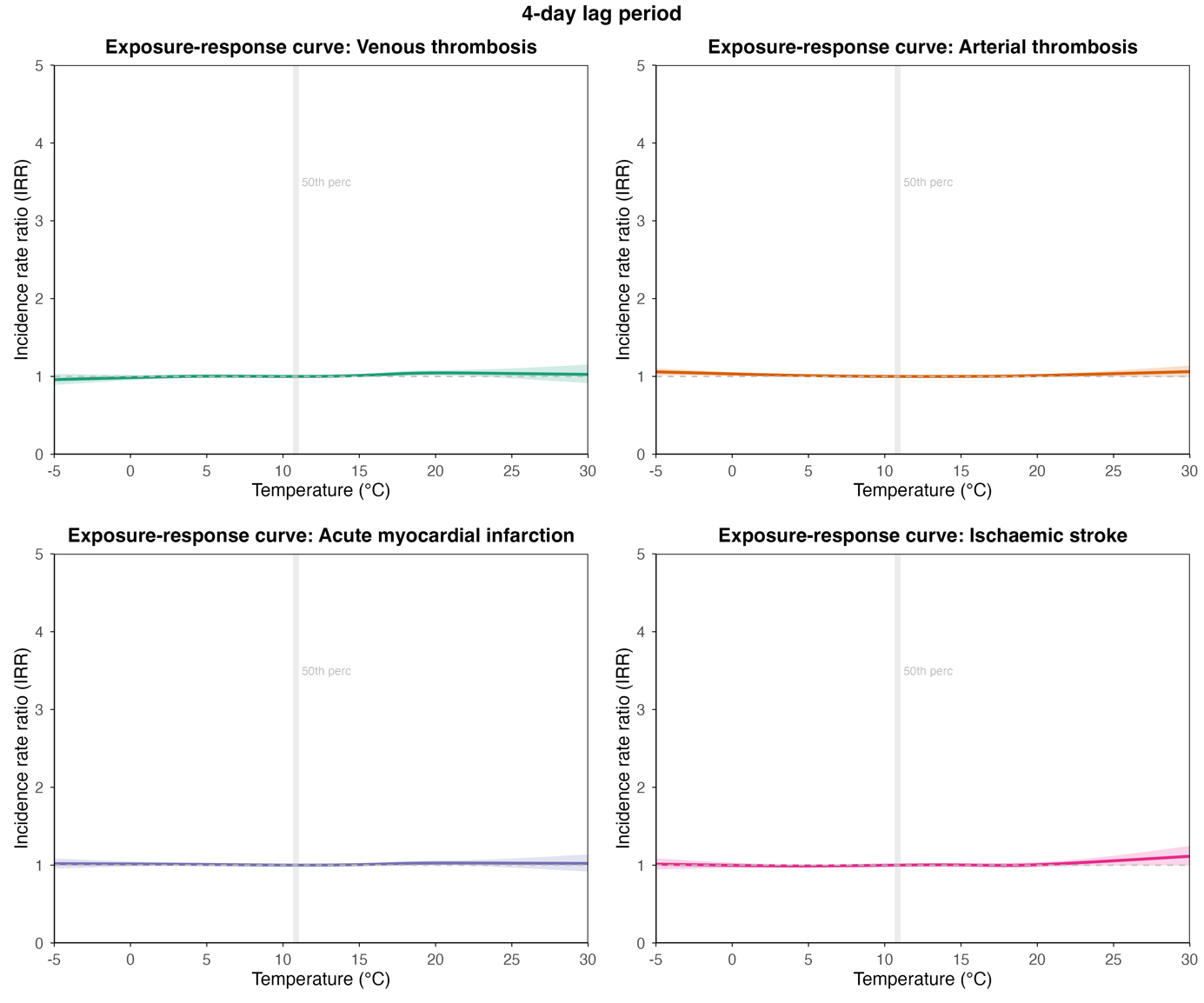

#### **Figure S28. Cumulative IRR over 7-day lag period.**

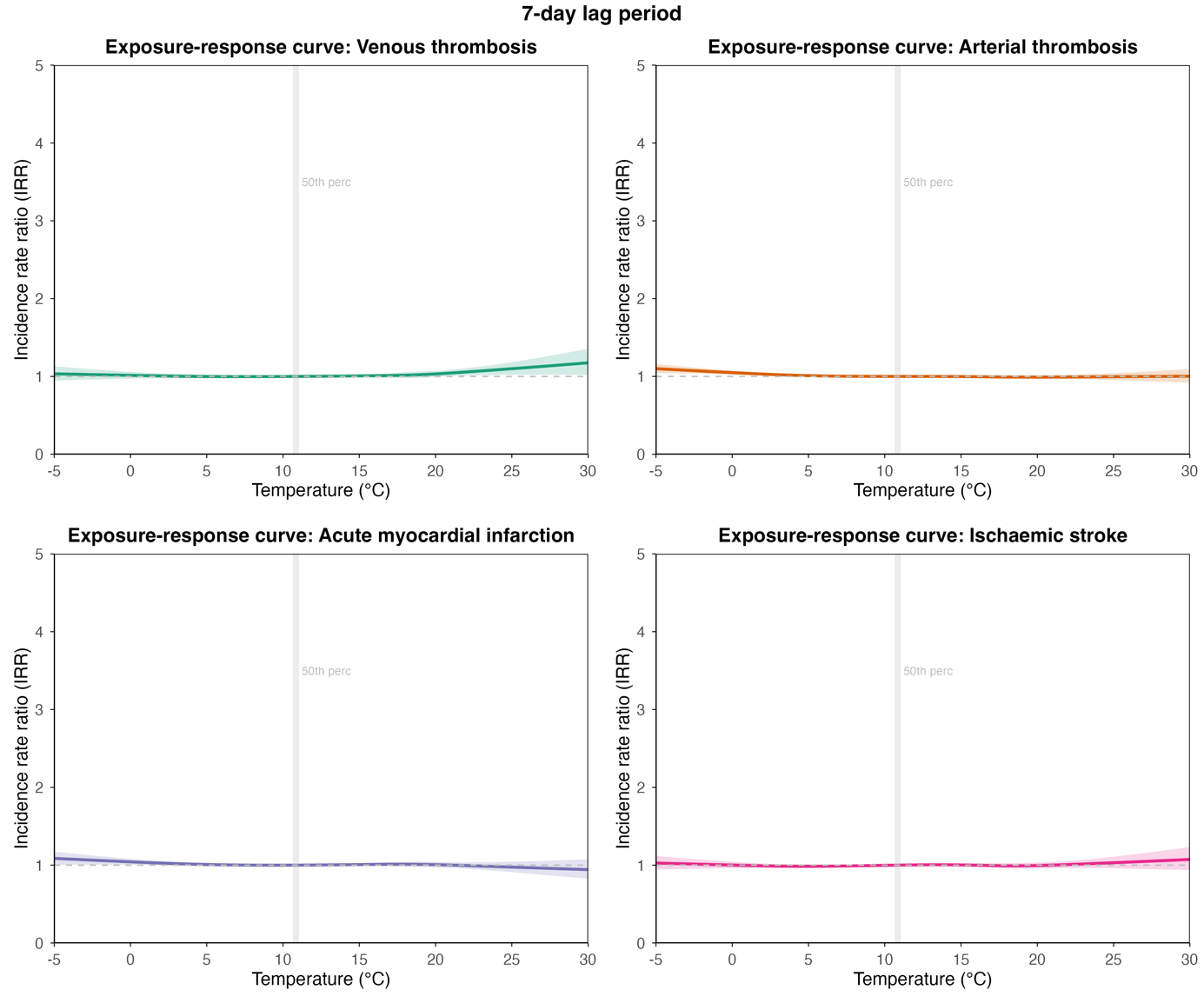

#### **Figure S29. Cumulative IRR over 14-day lag period.**

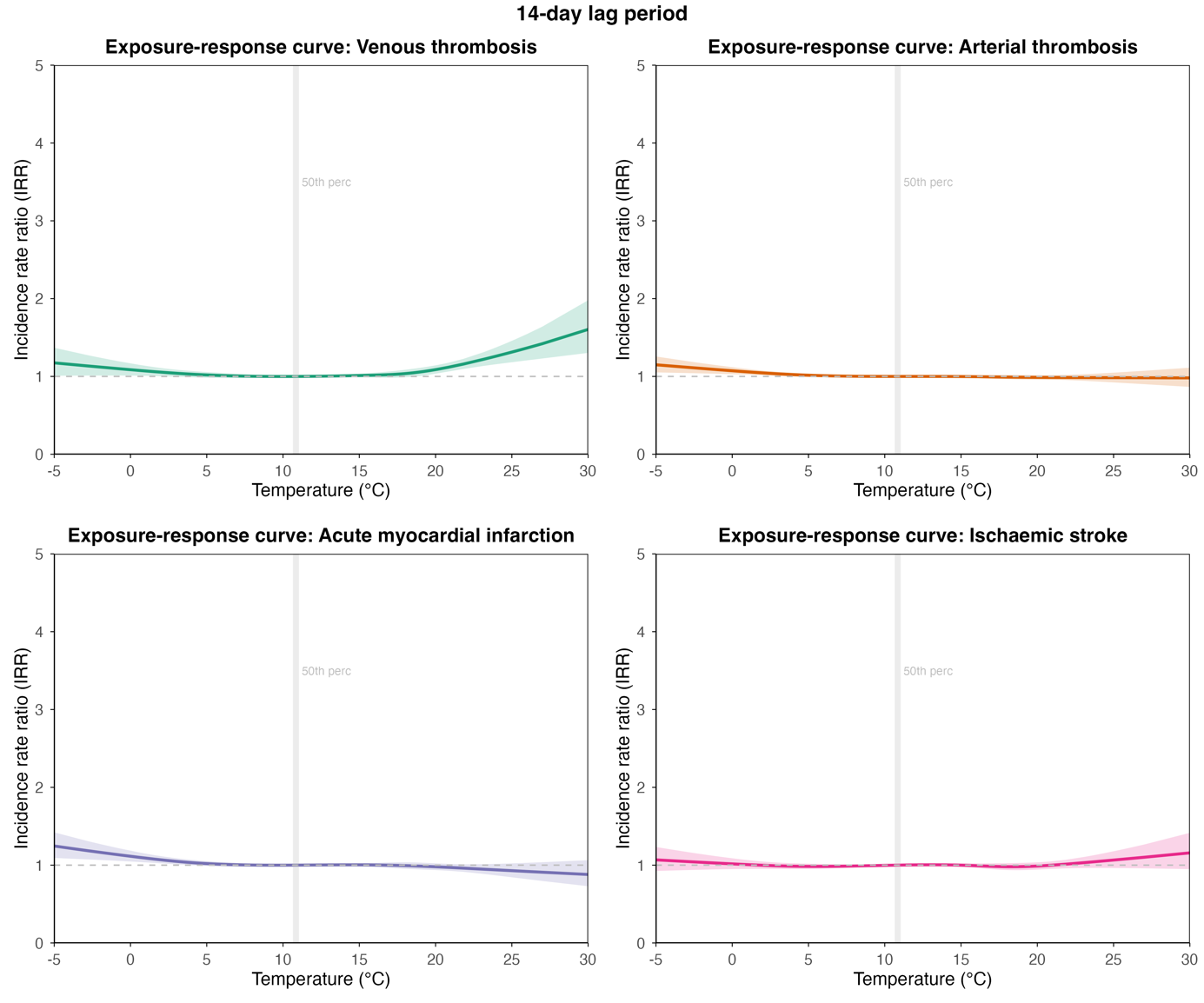

#### **Figure S30. Exposure-response curve acute myocardial infarction over 14-day lag period subgroups sex, age group, ethnic group, rural/urban area.**

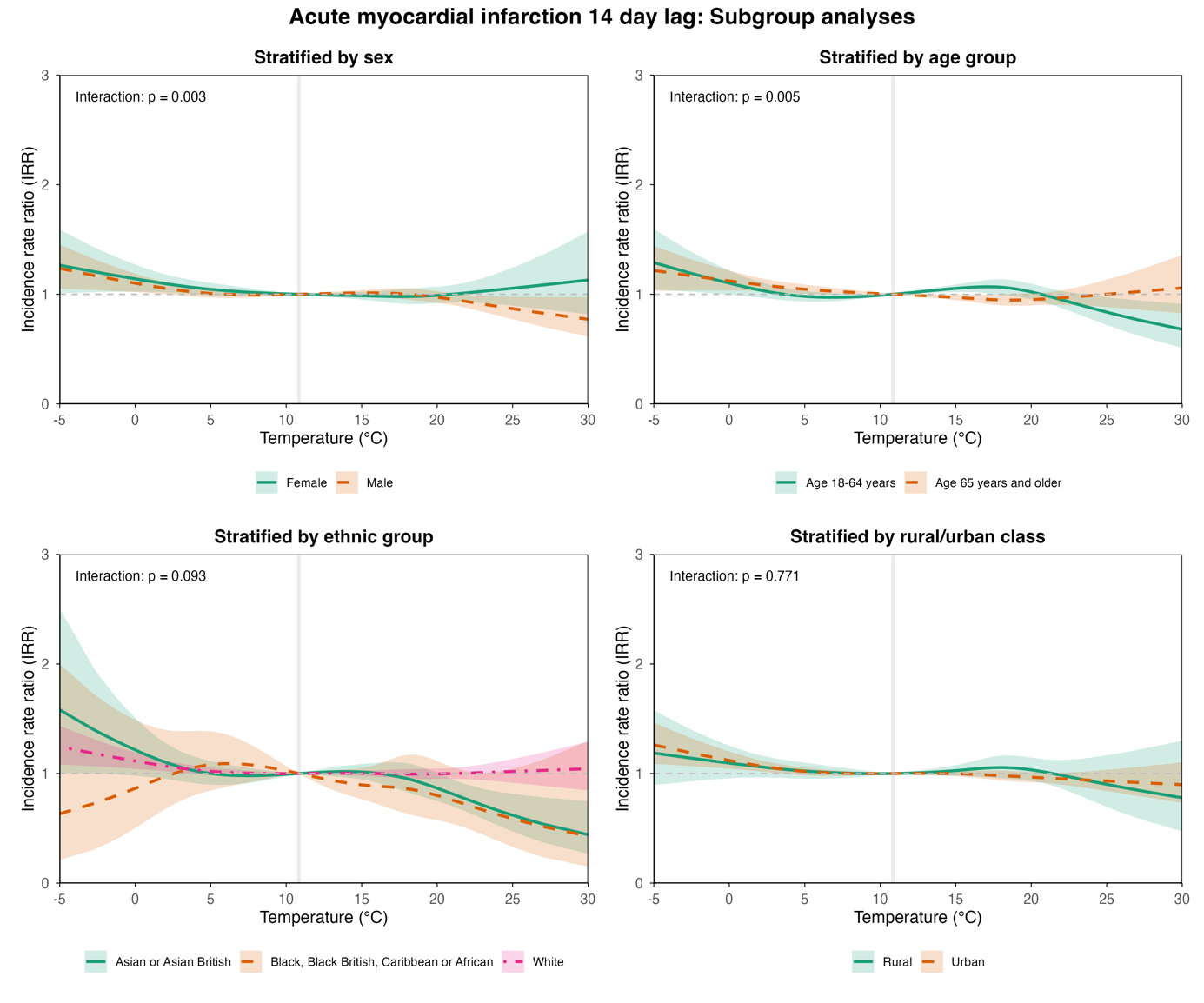

#### **Figure S31. Exposure-response curve acute myocardial infarction over 14-day lag period subgroups deprivation index, comorbidities, excess weight, smoking status.**

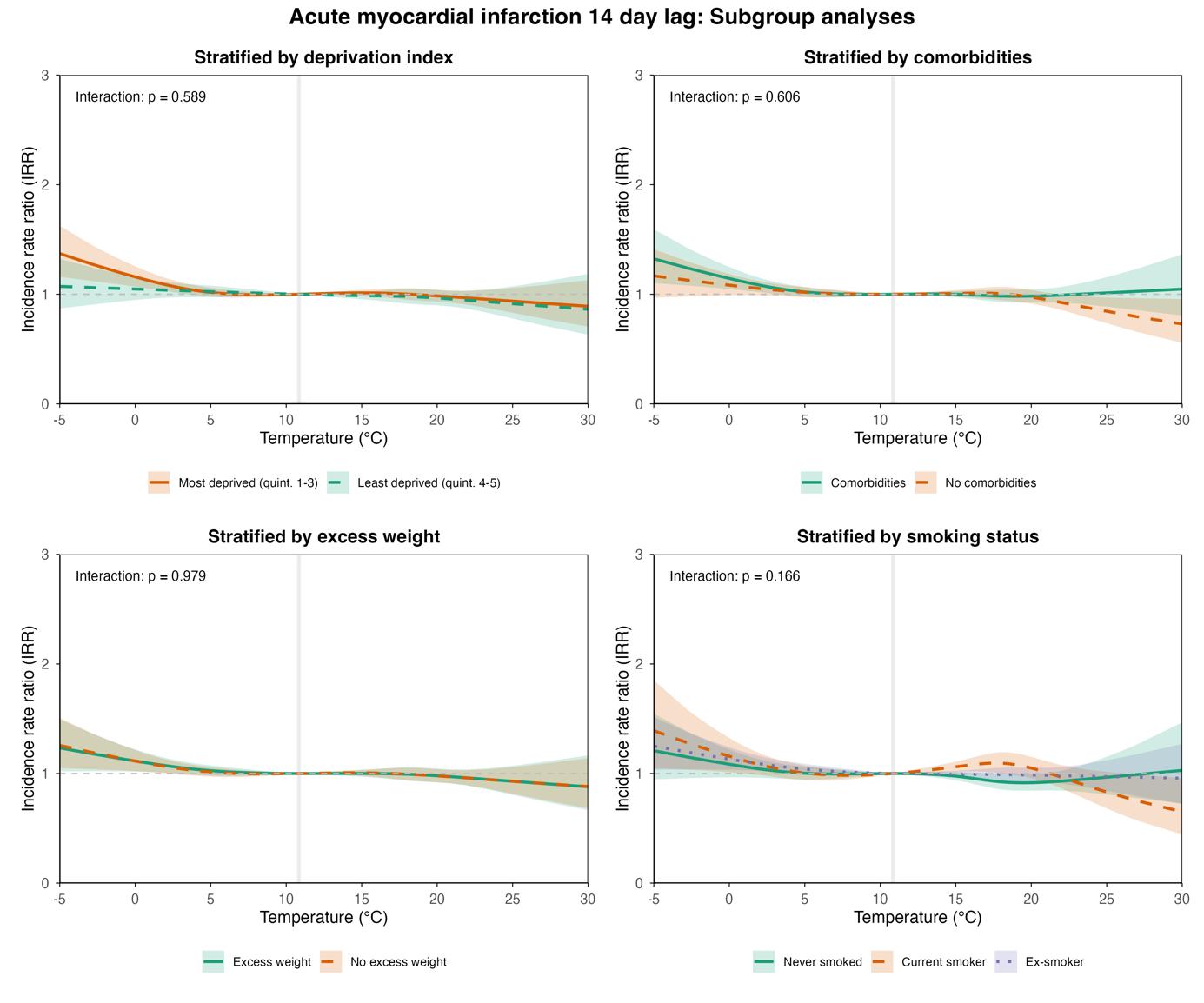

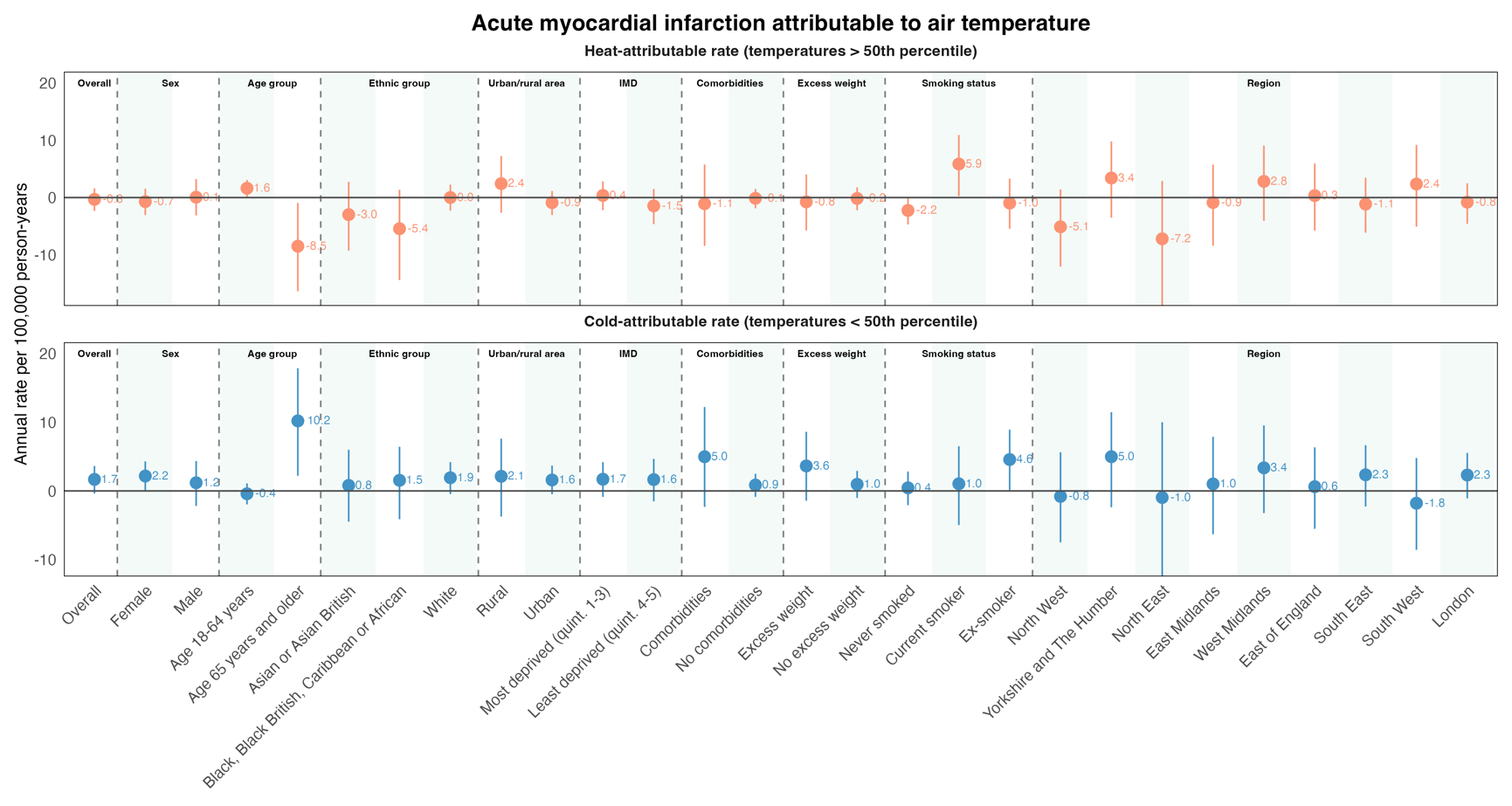

#### **Figure S32. Acute myocardial infarction attributable to heat and cold, stratified by demographic, socioeconomic, clinical, and regional subgroups, considering a 14-day lag period.**

### **Tables**

#### **Table S1. NHS digital data sources**

| **Data source** | **Description*** | **Data coverage** |
| --- | --- | --- |
| GPES Data for Pandemic Planning and Research (GDPPR) dataset | The GPES Data for Pandemic Planning and Research (GDPPR) dataset was established by NHS England to support the response to the coronavirus outbreak. The dataset is a central collection of GP patient data including the period since the start of the COVID-19 pandemic and a short period of time prior for comparison purposes. | 2018-2025 |
| Hospital Episode Statistics (HES)  Admitted Patient Care | The Hospital Episode Statistics (HES) datasets contain records of all admissions, appointments and attendances at NHS hospitals in England. Records in the HES Admitted Patient Care (HES APC) database are called ‘hospital episodes’, and each hospital episode relates to a period of care for a patient under a single consultant within one hospital provider. A stay in hospital from admission to discharge is called a ‘spell’ and can be made up of one or more episodes of care. | 1997-2025 |
| HES Outpatients (HES OP) | The Hospital Episode Statistics (HES) datasets contain records of all admissions, appointments and attendances at NHS hospitals in England. HES Outpatients (HES OP) is a record-level patient dataset of patients attending outpatient clinics at NHS hospitals in England. A record represents one appointment. | 2003-2025 |
| HES Accident and Emergency (HES AE) | The Hospital Episode Statistics (HES) datasets contain records of all admissions, appointments and attendances at NHS hospitals in England. HES Accident and Emergency (HES AE) is a record-level dataset of patients attending Accident and Emergency Departments (including minor injury units and walk-in centres) in England. A record represents one attendance. | 2007-2025 |
| Civil Registration of Deaths | The Civil Registration of Deaths dataset contains details of all registered deaths in England and Wales since 1993, as provided by the ONS. The dataset contains basic demographics of the deceased person, cause of death, records of other diagnoses, which may – or may not – have directly contributed to the death and information about registration itself. | 1992-2025 |
| COVID-19 UK non-hospital antigen testing results (Pillar 2) | The COVID-19 UK non-hospital antigen testing results (Pillar 2) dataset includes data on antigen swab COVID-19 testing, conducted via drive-through test centres, mobile testing units, satellite test centres, home testing and care home testing. It consists of records from the entire UK, but only English data is provided by default. This dataset is no longer updated | 2020-2024 |
| COVID-19 UK non-hospital antibody testing results (Pillar 3) | The COVID-19 UK non-hospital antibody testing results (Pillar 3) dataset, also referred to as iElisa, documents individuals that have undergone a finger prick test for antibodies from having had COVID-19. The dataset is UK-wide and contains positive, negative and void results. It also contains demographic data. Data available is in relation to specified cohorts which differ across geography and time. Data does not include the NHS Antibody tests as NHS Digital does not hold this data. This dataset is no longer updated. | 2020-2023 |
| Secondary Uses Service (SUS) | The Secondary Uses Service (SUS) is the single, comprehensive repository for healthcare data in England which enables a range of reporting and analyses to support the NHS in the delivery of healthcare services. SUS is a collection of health care data required by hospitals and used for planning healthcare, supporting payments, commissioning policy development and research. This dataset is no longer updated. | 2019-2022 |
| COVID-19 SARI-Watch dataset (formerly CHESS) | Data forming the COVID-19 SARI-Watch dataset (formerly CHESS) relates to demographic, risk factor, treatment and outcome information for patients admitted to hospital with a confirmed COVID-19 diagnosis, as recorded in the COVID-19 SARI-Watch surveillance. Note that SARI-Watch replaced CHESS which was initiated across all NHS Trusts in England on 15 March 2020. SARI-Watch replaced CHESS in summer 2020 and collects the same data items as CHESS, but includes infections other than COVID-19. | 2019-2024 |
| COVID-19 Second Generation Surveillance System (SGSS) | The COVID-19 Second Generation Surveillance System (SGSS) includes demographic and diagnostic information concerning antigen test reports for COVID-19 in England. This dataset currently includes the first positive results from Pillar 1 (swab testing in PHE and NHS hospital labs), and Pillar 2 (swab testing in the wider population at drive through test centres, walk-in centres, home kits returned by post etc). Future flows are anticipated to include all positive, negative, void and inconclusive test results. This dataset is no longer updated. | 2020-2024 |
| HES Critical Care (HES CC) | The Hospital Episode Statistics (HES) datasets contain records of all admissions, appointments and attendances at NHS hospitals in England. The HES Critical Care (HES CC) dataset is a subset of HES Admitted Patient Care (HES APC) data. It includes record-level patient data of patients admitted for treatment and receiving Critical Care (intensive care or high dependency care) at NHS hospitals in England. A record represents one episode of Critical Care. | 2007-2025 |
| COVID-19 vaccination status dataset | The COVID-19 vaccination status dataset records individual vaccination events, details of the patients, and batch information on the vaccine for anyone vaccinated within England, or those vaccinated in a devolved administration where this information was passed to England. | 2020-2025 |
| *Descriptions are extracted from the [British Heart Foundation Dataset Summary dashboard.](https://bhfdatasciencecentre.org/dashboard/) | | |

#### **Table S2. STROBE statement**

|  | Item No | Recommendation | **Page No.** |
| --- | --- | --- | --- |
| **Title and abstract** | 1 | (*a*) Indicate the study’s design with a commonly used term in the title or the abstract | Main text p. 1, 6 |
|  |  | (*b*) Provide in the abstract an informative and balanced summary of what was done and what was found | Main text p. 4 |
| Introduction | | |  |
| Background/rationale | 2 | Explain the scientific background and rationale for the investigation being reported | Main text p. 5  Supplement p. 9, 25 |
| Objectives | 3 | State specific objectives, including any prespecified hypotheses | Main text: p. 5-6 |
| Methods | | |  |
| Study design | 4 | Present key elements of study design early in the paper | Main text: p. 6-7  Supplement: p. 81, 84 |
| Setting | 5 | Describe the setting, locations, and relevant dates, including periods of recruitment, exposure, follow-up, and data collection | Main text: p.6-7  Supplement: p. 4-5 |
| Participants | 6 | (*a*) *Cohort study*—Give the eligibility criteria, and the sources and methods of selection of participants. Describe methods of follow-up  *Case-control study*—Give the eligibility criteria, and the sources and methods of case ascertainment and control selection. Give the rationale for the choice of cases and controls  *Cross-sectional study*—Give the eligibility criteria, and the sources and methods of selection of participants | Main text: p. 6-7  Supplement: p. 84 |
|  |  | (*b*) *Cohort study*—For matched studies, give matching criteria and number of exposed and unexposed  *Case-control study*—For matched studies, give matching criteria and the number of controls per case | NA. |
| Variables | 7 | Clearly define all outcomes, exposures, predictors, potential confounders, and effect modifiers. Give diagnostic criteria, if applicable | Main text: p. 6-7  Supplement: p. 4-5 |
| Data sources/ measurement | 8* | For each variable of interest, give sources of data and details of methods of assessment (measurement). Describe comparability of assessment methods if there is more than one group | Main text: p. 6-7  Supplement: p. 4-5, 71-72 |
| Bias | 9 | Describe any efforts to address potential sources of bias | Main text: p. 7  Supplement: p. 84 |
| Study size | 10 | Explain how the study size was arrived at | Supplement: p. 40 |
| Quantitative variables | 11 | Explain how quantitative variables were handled in the analyses. If applicable, describe which groupings were chosen and why | Main text: p. 6-7  Supplement: p. 4-5 |
| Statistical methods | 12 | (*a*) Describe all statistical methods, including those used to control for confounding | Main text: p. 7  Supplement: p. 84 |
|  |  | (*b*) Describe any methods used to examine subgroups and interactions | Main text: p. 7 |
|  |  | (*c*) Explain how missing data were addressed | Covariates and outcomes were derived from routinely collected healthcare records using diagnostic codes. Inclusion required a valid LSOA within England, and the environmental dataset had complete coverage. For diagnostic variables, the absence of a recorded code was assumed to indicate absence of the condition, acknowledging the potential for under-ascertainment. There were therefore no missing data for the variables used in the analysis, except for ethnic group and smoking status, for which missingness was retained as a separate category in interaction analyses. |
|  |  | (*d*) *Cohort study*—If applicable, explain how loss to follow-up was addressed  *Case-control study*—If applicable, explain how matching of cases and controls was addressed  *Cross-sectional study*—If applicable, describe analytical methods taking account of sampling strategy | Main text: p. 7  Supplement: p. 84 |
|  |  | (*e*) Describe any sensitivity analyses | Main text: p. 7  Supplement: p. 84 |

Continued on next page

| Results | | |  |
| --- | --- | --- | --- |
| Participants | 13* | (a) Report numbers of individuals at each stage of study—eg numbers potentially eligible, examined for eligibility, confirmed eligible, included in the study, completing follow-up, and analysed | Supplement: p. 40 |
|  |  | (b) Give reasons for non-participation at each stage | Supplement: p. 40 |
|  |  | (c) Consider use of a flow diagram | Supplement: p. 40 |
| Descriptive data | 14* | (a) Give characteristics of study participants (eg demographic, clinical, social) and information on exposures and potential confounders | Main text: p. 8-11 |
|  |  | (b) Indicate number of participants with missing data for each variable of interest | Main text: p.9-11 |
|  |  | (c) *Cohort study*—Summarise follow-up time (eg, average and total amount) | Supplement: p. 86 |
| Outcome data | 15* | *Cohort study*—Report numbers of outcome events or summary measures over time | Main text: p. 8-11  Supplement: p. 86-89, Excel Table S11 |
|  |  | *Case-control study—*Report numbers in each exposure category, or summary measures of exposure | NA. |
|  |  | *Cross-sectional study—*Report numbers of outcome events or summary measures | NA. |
| Main results | 16 | (*a*) Give unadjusted estimates and, if applicable, confounder-adjusted estimates and their precision (eg, 95% confidence interval). Make clear which confounders were adjusted for and why they were included | Main text: p.7, 8-9 |
|  |  | (*b*) Report category boundaries when continuous variables were categorized | Main text: p. 6  Supplement: p. 3-4 |
|  |  | (*c*) If relevant, consider translating estimates of relative risk into absolute risk for a meaningful time period | Main text: p. 9  Supplement: Excel Table S16 |
| Other analyses | 17 | Report other analyses done—eg analyses of subgroups and interactions, and sensitivity analyses | Main text: p. 8-9  Supplement: p. 84 |
| Discussion | | |  |
| Key results | 18 | Summarise key results with reference to study objectives | Main text: p.17-18 |
| Limitations | 19 | Discuss limitations of the study, taking into account sources of potential bias or imprecision. Discuss both direction and magnitude of any potential bias | Main text: p. 18 |
| Interpretation | 20 | Give a cautious overall interpretation of results considering objectives, limitations, multiplicity of analyses, results from similar studies, and other relevant evidence | Main text: p. 17-18 |
| Generalisability | 21 | Discuss the generalisability (external validity) of the study results | These findings, derived from a whole-population cohort in England during the COVID-19 pandemic, may not be directly generalisable to settings with different climatic conditions, healthcare systems, or population structures, although they are likely relevant to similar temperate, high-income contexts. |
| Other information | | |  |
| Funding | 22 | Give the source of funding and the role of the funders for the present study and, if applicable, for the original study on which the present article is based | Main text: p. 2-3, 18 |

#### **Table S3. Codelist covariates**

See excel file "S2_codelist_covariates.xlsx"

#### **Table S4. Codelist outcomes**

| **name** | **terminology** | **code** | **term** |
| --- | --- | --- | --- |
| ADISS | ICD10 | I670 | Dissection of cerebral arteries, nonruptured |
| ADISS | ICD10 | I71 | Aortic aneurysm and dissection |
| ADISS | ICD10 | I710 | Dissection of aorta [any part] |
| ADISS | ICD10 | I711 | Thoracic aortic aneurysm, ruptured |
| ADISS | ICD10 | I713 | Abdominal aortic aneurysm, ruptured |
| ADISS | ICD10 | I715 | Thoracoabdominal aortic aneurysm, ruptured |
| ADISS | ICD10 | I718 | Aortic aneurysm of unspecified site, ruptured |
| ADISS | ICD10 | I72 | Other aneurysm and dissection |
| ADISS | ICD10 | I720 | Aneurysm and dissection of carotid artery |
| ADISS | ICD10 | I721 | Aneurysm and dissection of artery of upper extremity |
| ADISS | ICD10 | I722 | Aneurysm and dissection of renal artery |
| ADISS | ICD10 | I723 | Aneurysm and dissection of iliac artery |
| ADISS | ICD10 | I724 | Aneurysm and dissection of artery of lower extremity |
| ADISS | ICD10 | I725 | Aneurysm and dissection of other precerebral arteries |
| ADISS | ICD10 | I728 | Aneurysm and dissection of other specified arteries |
| ADISS | ICD10 | I729 | Aneurysm and dissection of unspecified site |
| AT | ICD10 | I74 | Arterial embolism and thrombosis |
| AT | ICD10 | I740 | Embolism and thrombosis of abdominal aorta |
| AT | ICD10 | I741 | Embolism and thrombosis of other and unspecified parts of aorta |
| AT | ICD10 | I742 | Embolism and thrombosis of arteries of upper extremities |
| AT | ICD10 | I743 | Embolism and thrombosis of arteries of lower extremities |
| AT | ICD10 | I744 | Embolism and thrombosis of arteries of extremities, unspecified |
| AT | ICD10 | I745 | Embolism and thrombosis of iliac artery |
| AT | ICD10 | I748 | Embolism and thrombosis of other arteries |
| AT | ICD10 | I749 | Embolism and thrombosis of unspecified artery |
| AT | ICD10 | N280 | Ischaemia and infarction of kidney |
| ICVT | ICD10 | G08 | Intracranial and intraspinal phlebitis and thrombophlebitis |
| ICVT | ICD10 | I636 | Cerebral infarction due to cerebral venous thrombosis, nonpyogenic |
| ICVT | ICD10 | I676 | Nonpyogenic thrombosis of intracranial venous system |
| ICVT | ICD10 | O225 | Cerebral venous thrombosis in pregnancy |
| ICVT | ICD10 | O873 | Cerebral venous thrombosis in the puerperium |
| MI | ICD10 | I21 | Acute myocardial infarction |
| MI | ICD10 | I210 | Acute transmural myocardial infarction of anterior wall |
| MI | ICD10 | I211 | Acute transmural myocardial infarction of inferior wall |
| MI | ICD10 | I212 | Acute transmural myocardial infarction of other sites |
| MI | ICD10 | I213 | Acute transmural myocardial infarction of unspecified site |
| MI | ICD10 | I214 | Acute subendocardial myocardial infarction |
| MI | ICD10 | I219 | Acute myocardial infarction, unspecified |
| MI | ICD10 | I22 | Subsequent myocardial infarction |
| MI | ICD10 | I220 | Subsequent myocardial infarction of anterior wall |
| MI | ICD10 | I221 | Subsequent myocardial infarction of inferior wall |
| MI | ICD10 | I228 | Subsequent myocardial infarction of other sites |
| MI | ICD10 | I229 | Subsequent myocardial infarction of unspecified site |
| MI | ICD10 | I23 | Certain current complications following acute myocardial infarction |
| MI | ICD10 | I230 | Haemopericardium as current complication following acute myocardial infarction |
| MI | ICD10 | I231 | Atrial septal defect as current complication following acute myocardial infarction |
| MI | ICD10 | I232 | Ventricular septal defect as current complication following acute myocardial infarction |
| MI | ICD10 | I233 | Rupture of cardiac wall without haemopericardium as current complication following acute myocardial infarction |
| MI | ICD10 | I234 | Rupture of chordae tendineae as current complication following acute myocardial infarction |
| MI | ICD10 | I235 | Rupture of papillary muscle as current complication following acute myocardial infarction |
| MI | ICD10 | I236 | Thrombosis of atrium, auricular appendage, and ventricle as current complications following acute myocardial infarction |
| MI | ICD10 | I238 | Other current complications following acute myocardial infarction |
| PE | ICD10 | I26 | Pulmonary embolism |
| PE | ICD10 | I260 | Pulmonary embolism with mention of acute cor pulmonale |
| PE | ICD10 | I269 | Pulmonary embolism without mention of acute cor pulmonale |
| RI | ICD10 | H341 | Central retinal artery occlusion |
| RI | ICD10 | H342 | Other retinal artery occlusions |
| VT | ICD10 | I80 | Phlebitis and thrombophlebitis |
| VT | ICD10 | I800 | Phlebitis and thrombophlebitis of superficial vessels of lower extremities |
| VT | ICD10 | I801 | Phlebitis and thrombophlebitis of femoral vein |
| VT | ICD10 | I802 | Phlebitis and thrombophlebitis of other deep vessels of lower extremities |
| VT | ICD10 | I803 | Phlebitis and thrombophlebitis of lower extremities, unspecified |
| VT | ICD10 | I808 | Phlebitis and thrombophlebitis of other sites |
| VT | ICD10 | I809 | Phlebitis and thrombophlebitis of unspecified site |
| VT | ICD10 | I81 | Portal vein thrombosis |
| VT | ICD10 | I82 | Other venous embolism and thrombosis |
| VT | ICD10 | I820 | Budd-Chiari syndrome |
| VT | ICD10 | I821 | Thrombophlebitis migrans |
| VT | ICD10 | I822 | Embolism and thrombosis of vena cava |
| VT | ICD10 | I823 | Embolism and thrombosis of renal vein |
| VT | ICD10 | I828 | Embolism and thrombosis of other specified veins |
| VT | ICD10 | I829 | Embolism and thrombosis of unspecified vein |
| VT | ICD10 | O082 | Embolism following abortion and ectopic and molar pregnancy |
| VT | ICD10 | O222 | Superficial thrombophlebitis in pregnancy |
| VT | ICD10 | O223 | Deep phlebothrombosis in pregnancy |
| VT | ICD10 | O870 | Superficial thrombophlebitis in the puerperium |
| VT | ICD10 | O871 | Deep phlebothrombosis in the puerperium |
| VT | ICD10 | O879 | Venous complication in the puerperium, unspecified |
| VT | ICD10 | O882 | Obstetric blood-clot embolism |
| stroke_IS | ICD10 | I63 | Cerebral infarction |
| stroke_IS | ICD10 | I630 | Cerebral infarction due to thrombosis of precerebral arteries |
| stroke_IS | ICD10 | I631 | Cerebral infarction due to embolism of precerebral arteries |
| stroke_IS | ICD10 | I632 | Cerebral infarction due to unspecified occlusion or stenosis of precerebral arteries |
| stroke_IS | ICD10 | I633 | Cerebral infarction due to thrombosis of cerebral arteries |
| stroke_IS | ICD10 | I634 | Cerebral infarction due to embolism of cerebral arteries |
| stroke_IS | ICD10 | I635 | Cerebral infarction due to unspecified occlusion or stenosis of cerebral arteries |
| stroke_IS | ICD10 | I636 | Cerebral infarction due to cerebral venous thrombosis, nonpyogenic |
| stroke_IS | ICD10 | I638 | Other cerebral infarction |
| stroke_IS | ICD10 | I639 | Cerebral infarction, unspecified |
| stroke_NOS | ICD10 | G46 | Vascular syndromes of brain in cerebrovascular diseases |
| stroke_NOS | ICD10 | G460 | Middle cerebral artery syndrome |
| stroke_NOS | ICD10 | G461 | Anterior cerebral artery syndrome |
| stroke_NOS | ICD10 | G463 | Brain stem stroke syndrome |
| stroke_NOS | ICD10 | G464 | Cerebellar stroke syndrome |
| stroke_NOS | ICD10 | G465 | Pure motor lacunar syndrome |
| stroke_NOS | ICD10 | G466 | Pure sensory lacunar syndrome |
| stroke_NOS | ICD10 | G467 | Other lacunar syndromes |
| stroke_NOS | ICD10 | G468 | Other vascular syndromes of brain in cerebrovascular diseases |
| stroke_NOS | ICD10 | I64 | Stroke, not specified as haemorrhage or infarction |

#### **Table S5. Example time series dataset for one case with complete follow-up.**

| **PERSON_ID** | **DATE** | **LSOA** | **OUTCOME_INDICATOR** | **TMEAN (°C)** | | **PM2.5 (µg/m³)** | **WEEKDAY** | **MONTH** | **YEAR** |
| --- | --- | --- | --- | --- | --- | --- | --- | --- | --- |
| 100001 | 2020-01-01 | E01018223 | 0 | 4.8 | 12.3 | | 3 | 1 | 2020 |
| 100001 | 2020-01-02 | E01018223 | 0 | 5.1 | 10.9 | | 4 | 1 | 2020 |
| 100001 | 2020-01-03 | E01018223 | 0 | 5.6 | 11.7 | | 5 | 1 | 2020 |
| 100001 | 2020-01-04 | E01018223 | 0 | 4.9 | 9.5 | | 6 | 1 | 2020 |
| 100001 | 2020-01-05 | E01018223 | 0 | 5.3 | 8.8 | | 7 | 1 | 2020 |
| 100001 | 2020-01-06 | E01018223 | 1 | 4.7 | 10.4 | | 1 | 1 | 2020 |
| 100001 | 2020-01-07 | E01018223 | 0 | 3.9 | 11.2 | | 2 | 1 | 2020 |
| 100001 | 2020-01-08 | E01018223 | 0 | 4.1 | 10.1 | | 3 | 1 | 2020 |
| 100001 | 2020-01-09 | E01018223 | 0 | 4.5 | 9.9 | | 4 | 1 | 2020 |
| 100001 | 2020-01-10 | E01018223 | 0 | 5 | 11 | | 5 | 1 | 2020 |
| 100001 | [REST OF FOLLOW-UP] | | | | | | | | |
| 100001 | 2022-12-22 | E01000001 | 0 | 6.1 | * | | 4 | 12 | 2022 |
| 100001 | 2022-12-23 | E01000001 | 0 | 5.4 |  | | 5 | 12 | 2022 |
| 100001 | 2022-12-24 | E01000001 | 0 | 4.8 |  | | 6 | 12 | 2022 |
| 100001 | 2022-12-25 | E01000001 | 0 | 5.2 |  | | 7 | 12 | 2022 |
| 100001 | 2022-12-26 | E01000001 | 1 | 4.9 |  | | 1 | 12 | 2022 |
| 100001 | 2022-12-27 | E01000001 | 0 | 4.6 |  | | 2 | 12 | 2022 |
| 100001 | 2022-12-28 | E01000001 | 0 | 4.4 |  | | 3 | 12 | 2022 |
| 100001 | 2022-12-29 | E01000001 | 0 | 4.2 |  | | 4 | 12 | 2022 |
| 100001 | 2022-12-30 | E01000001 | 0 | 3.9 |  | | 5 | 12 | 2022 |
| 100001 | 2022-12-31 | E01000001 | 0 | 3.7 |  | | 6 | 12 | 2022 |
| * Air pollution data is available in the SDE until 2021-12-31. | | | | | | | | | |

#### **Table S6. Spline parameterisation results.**

| **Outcome** | **Lag days** | **Model max batch size** | **Spline parameterisation** | **Average AIC** |
| --- | --- | --- | --- | --- |
| Venous thrombotic events | 21 | 202,790 individuals | **Daily mean temperature:** natural spline, 4 knots at 5^th^ 35^th^ 65^th^ 95^th^ percentile, boundary knots at 1^st^ and 99^th^ percentile.  **Lag days:**  natural spline, 2 knots on log scale. | 2312983.3 |
| Venous thrombotic events | 21 | 202,790 individuals | **Daily mean temperature:** natural spline, 4 knots at 10^th^ 40^th^ 70^th^ 90^th^ percentile, boundary knots at 1^st^ and 99^th^ percentile.  **Lag days:**  natural spline, 2 knots on log scale. | 2312986.8 |
| Venous thrombotic events | 21 | 202,790 individuals | **Daily mean temperature:** natural spline, 4 knots at 5^th^ 35^th^ 65^th^ 95^th^ percentile, boundary knots at 1^st^ and 99^th^ percentile.  **Lag days:**  natural spline, 3 knots on log scale. | 2312986.0 |
| Venous thrombotic events | 21 | 202,790 individuals | **Daily mean temperature:** natural spline, 4 knots at 10^th^ 40^th^ 70^th^ 90^th^ percentile, boundary knots at 1^st^ and 99^th^ percentile.  **Lag days:**  natural spline, 3 knots on log scale. | 2312989.2 |
| Venous thrombotic events | 21 | 202,790 individuals | **Daily mean temperature:** natural spline, 4 knots at 5^th^ 35^th^ 65^th^ 95^th^ percentile, boundary knots at 1^st^ and 99^th^ percentile.  **Lag days:**  natural spline, 4 knots on log scale. | 2312985.0 |
| Venous thrombotic events | 21 | 202,790 individuals | **Daily mean temperature:** natural spline, 4 knots at 10^th^ 40^th^ 70^th^ 90^th^ percentile, boundary knots at 1^st^ and 99^th^ percentile.  **Lag days:**  natural spline, 4 knots on log scale. | 2312988.4 |
| Venous thrombotic events | 21 | 202,790 individuals | **Daily mean temperature:** natural spline, 2 knots at 10^th,^ 90^th^ percentile, boundary knots at 1^st^ and 99^th^ percentile.  **Lag days:**  natural spline, 2 knots on log scale. | 2313037.8 |
| Venous thrombotic events | 21 | 202,790 individuals | **Daily mean temperature:** natural spline, 2 knots at 25^th,^ 75^th^ percentile, boundary knots at 1^st^ and 99^th^ percentile.  **Lag days:**  natural spline, 2 knots on log scale. | 2313042.5 |
| Venous thrombotic events | 21 | 202,790 individuals | **Daily mean temperature:** natural spline, 2 knots at 10^th,^ 90^th^ percentile, boundary knots at 1^st^ and 99^th^ percentile.  **Lag days:**  natural spline, 3 knots on log scale. | 2313037.0 |
| Venous thrombotic events | 21 | 202,790 individuals | **Daily mean temperature:** natural spline, 2 knots at 25^th,^ 75^th^ percentile, boundary knots at 1^st^ and 99^th^ percentile.  **Lag days:**  natural spline, 3 knots on log scale. | 2313041.3 |
| Venous thrombotic events | 21 | 202,790 individuals | **Daily mean temperature:** natural spline, 2 knots at 10^th^, 90th percentile, boundary knots at 1^st^ and 99^th^ percentile.  **Lag days:**  natural spline, 4 knots on log scale. | 2313037.7 |
| Venous thrombotic events | 21 | 202,790 individuals | **Daily mean temperature:** natural spline, 2 knots at 25^th,^ 75^th^ percentile, boundary knots at 1^st^ and 99^th^ percentile.  **Lag days:**  natural spline, 4 knots on log scale. | 2313041.5 |
| Venous thrombotic events | 21 | 202,790 individuals | **Daily mean temperature:** natural spline, 3 knots at 10^th^, 75^th^, 90^th^ percentile, boundary knots at 1^st^ and 99^th^ percentile.  **Lag days:**  natural spline, 2 knots on log scale. | 2313034.6 |
| Venous thrombotic events | 21 | 202,790 individuals | **Daily mean temperature:** natural spline, 3 knots at 5^th^, 50^th^, 95^th^ percentile, boundary knots at 1^st^ and 99^th^ percentile.  **Lag days:**  natural spline, 2 knots on log scale. | 2313043.1 |
| Venous thrombotic events | 21 | 202,790 individuals | **Daily mean temperature:** natural spline, 3 knots at 10^th^, 75^th^, 90^th^ percentile, boundary knots at 1^st^ and 99^th^ percentile.  **Lag days:**  natural spline, 3 knots on log scale. | 2313033.7 |
| Venous thrombotic events | 21 | 202,790 individuals | **Daily mean temperature:** natural spline, 3 knots at 5^th^, 50^th^, 95^th^ percentile, boundary knots at 1^st^ and 99^th^ percentile.  **Lag days:**  natural spline, 3 knots on log scale. | 2313043.2 |
| Venous thrombotic events | 21 | 202,790 individuals | **Daily mean temperature:** natural spline, 3 knots at 10^th^, 75^th^, 90^th^ percentile, boundary knots at 1^st^ and 99^th^ percentile.  **Lag days:**  natural spline, 4 knots on log scale. | 2313031.8 |
| Venous thrombotic events | 21 | 202,790 individuals s | **Daily mean temperature:** natural spline, 3 knots at 5^th^, 50^th^, 95^th^ percentile, boundary knots at 1^st^ and 99^th^ percentile.  **Lag days:**  natural spline, 4 knots on log scale. | 2313043.5 |

#### **Table S7. Main and sensitivity analyses elaboration.**

| **Analysis** | **Description** | **Aim** | **Case time series assumption^25^** |
| --- | --- | --- | --- |
| Main analysis | **Time series:** One row per day of follow-up per person. Follow-up continues until either the study end date (31-12-2022) or the last day of the month in which the person died.  **Crossbasis:**  • argvar: Natural splines with temperature knots at the 10th, 75th, and 90th percentiles of the distribution. Boundary knots at the 1st and 99th percentiles.  • arglag: Natural splines with 3 knots logarithmically spaced (higher resolution at early lags).  **Model:** Conditional Poisson regression  **Formula:** Outcome ~ Temperature crossbasis + Day of week  **Strata:** By each unique combination of person, month, and year. | Assess the association between air temperature and outcomes of interest. | Censoring is not done at actual date of death but at the last day of the month of death, such that if this is an event month, the assumption that outcomes cannot affect the duration of observation periods, is met.^25^ |
| Crossbasis spline parameterisation | Trying out different definitions of argvar and arglag in crossbasis() and testing AIC value. Based on venous events outcome. | Check for large differences in fit; if negligible, retain specification based on literature. | NA. |
| Restricted study period | Study start date at 01-04-2020 instead of 01-01-2020. | Assess if the reduction in all-cause hospitalisations in the beginning of the COVID-19 pandemic, may have impacted results, by excluding those months. | NA. |
| Different lag periods | Test associations across lag periods (4, 7, 14, 21 days) commonly described in literature. | To compare IRR at each temperature percentile, integrated over different lag periods. | NA. |
| Non-fatal cases only | Exclude events with fatal outcomes (different definitions used: death on or within 7, 14, 30 days of outcome event) | Compare non-fatal results to the main analysis and to the fatal analysis. Since follow-up in this analysis, like in the main analysis, is extended to the end of the death month, no outcome-dependent follow-up bias should exist. Differences may reflect different risk associations for severe (fatal) cases. | NA. |
| Fatal cases only | Exclude events with non-fatal outcomes (different definitions used: no death on or within 7, 14, 30 days of outcome event) | Compare fatal analysis results to the main analysis and to the non-fatal analysis. Since follow-up in this analysis, like in the main analysis, is extended to the end of the death month, no outcome-dependent follow-up bias should exist. Differences may reflect different risk associations for severe (fatal) cases. | NA |
| Censoring at death | Follow-up per person is till study end date (31-12-2022) or the actual date of death. | Compare results to the main analysis. Estimates may appear more pronounced due to selection bias (e.g., both temperature and CVD influence censoring via death risk). | Violated: Outcomes cannot affect the duration of observation periods. |
| First outcome event | Include only the first outcome event per person in the time series. | Compare results to the main analysis. Expect similar findings if the outcome events are independent. | The outcome must represent conditionally independent observations. |

#### **Table S8. Incidence rates of outcomes over study period.**

| **Outcome** | **Person years cohort*** | **Number of individuals*** | **Number of events*** | **Incidence rate** | **Subgroup** |
| --- | --- | --- | --- | --- | --- |
| Acute myocardial infarction | 144758730 | 49080575 | 275390 | 190.241374 | Overall cohort |
| Arterial thrombotic event | 144758730 | 49080575 | 604655 | 417.699847 | Overall cohort |
| Ischaemic stroke | 144758730 | 49080575 | 231485 | 159.909525 | Overall cohort |
| Venous thrombotic event | 144758730 | 49080575 | 215090 | 148.583783 | Overall cohort |
| Acute myocardial infarction | 5721985 | 1922355 | 4250 | 74.2400002 | Black, Black British, Caribbean or African |
| Arterial thrombotic event | 5721985 | 1922355 | 13045 | 227.962938 | Black, Black British, Caribbean or African |
| Ischaemic stroke | 5721985 | 1922355 | 6700 | 117.057326 | Black, Black British, Caribbean or African |
| Venous thrombotic event | 5721985 | 1922355 | 6520 | 113.946516 | Black, Black British, Caribbean or African |
| Acute myocardial infarction | 15204315 | 5099510 | 23220 | 152.706653 | Asian or Asian British |
| Arterial thrombotic event | 15204315 | 5099510 | 38110 | 250.665689 | Asian or Asian British |
| Ischaemic stroke | 15204315 | 5099510 | 11815 | 77.7147823 | Asian or Asian British |
| Venous thrombotic event | 15204315 | 5099510 | 6730 | 44.2769054 | Asian or Asian British |
| Acute myocardial infarction | 2447915 | 820875 | 1850 | 75.6562404 | Mixed or multiple ethnic groups |
| Arterial thrombotic event | 2447915 | 820875 | 4245 | 173.331225 | Mixed or multiple ethnic groups |
| Ischaemic stroke | 2447915 | 820875 | 1705 | 69.7328307 | Mixed or multiple ethnic groups |
| Venous thrombotic event | 2447915 | 820875 | 2130 | 87.0945489 | Mixed or multiple ethnic groups |
| Acute myocardial infarction | 113880610 | 38721535 | 241850 | 212.372417 | White |
| Arterial thrombotic event | 113880610 | 38721535 | 540935 | 475.001853 | White |
| Ischaemic stroke | 113880610 | 38721535 | 208445 | 183.038186 | White |
| Venous thrombotic event | 113880610 | 38721535 | 197020 | 173.005749 | White |
| Acute myocardial infarction | 3463170 | 1161040 | 2870 | 82.9297832 | Other ethnic group |
| Arterial thrombotic event | 3463170 | 1161040 | 5715 | 164.964433 | Other ethnic group |
| Ischaemic stroke | 3463170 | 1161040 | 2030 | 58.6168036 | Other ethnic group |
| Venous thrombotic event | 3463170 | 1161040 | 1840 | 53.159377 | Other ethnic group |
| Acute myocardial infarction | 14038980 | 4769400 | 31055 | 221.219738 | South West |
| Arterial thrombotic event | 14038980 | 4769400 | 68590 | 488.575299 | South West |
| Ischaemic stroke | 14038980 | 4769400 | 25685 | 182.954856 | South West |
| Venous thrombotic event | 14038980 | 4769400 | 21805 | 155.317525 | South West |
| Acute myocardial infarction | 18456835 | 6271990 | 42265 | 229.004598 | North West |
| Arterial thrombotic event | 18456835 | 6271990 | 89775 | 486.405181 | North West |
| Ischaemic stroke | 18456835 | 6271990 | 33130 | 179.489065 | North West |
| Venous thrombotic event | 18456835 | 6271990 | 31760 | 172.088013 | North West |
| Acute myocardial infarction | 23199540 | 7868425 | 40680 | 175.33969 | South East |
| Arterial thrombotic event | 23199540 | 7868425 | 93530 | 403.163182 | South East |
| Ischaemic stroke | 23199540 | 7868425 | 36660 | 158.024691 | South East |
| Venous thrombotic event | 23199540 | 7868425 | 37040 | 159.658344 | South East |
| Acute myocardial infarction | 14776740 | 5017760 | 30910 | 209.186872 | West Midlands |
| Arterial thrombotic event | 14776740 | 5017760 | 65725 | 444.786876 | West Midlands |
| Ischaemic stroke | 14776740 | 5017760 | 24725 | 167.317014 | West Midlands |
| Venous thrombotic event | 14776740 | 5017760 | 24805 | 167.858405 | West Midlands |
| Acute myocardial infarction | 25986145 | 8751175 | 29115 | 112.036632 | London |
| Arterial thrombotic event | 25986145 | 8751175 | 66585 | 256.232711 | London |
| Ischaemic stroke | 25986145 | 8751175 | 25815 | 99.3337083 | London |
| Venous thrombotic event | 25986145 | 8751175 | 23240 | 89.4284294 | London |
| Acute myocardial infarction | 6689580 | 2276150 | 16005 | 239.237654 | North East |
| Arterial thrombotic event | 6689580 | 2276150 | 35455 | 529.973291 | North East |
| Ischaemic stroke | 6689580 | 2276150 | 14395 | 215.200279 | North East |
| Venous thrombotic event | 6689580 | 2276150 | 13200 | 197.306786 | North East |
| Acute myocardial infarction | 12196190 | 4140795 | 24940 | 204.473682 | East Midlands |
| Arterial thrombotic event | 12196190 | 4140795 | 53770 | 440.891765 | East Midlands |
| Ischaemic stroke | 12196190 | 4140795 | 21165 | 173.554192 | East Midlands |
| Venous thrombotic event | 12196190 | 4140795 | 19370 | 158.828282 | East Midlands |
| Acute myocardial infarction | 15612430 | 5297230 | 28925 | 185.262633 | East of England |
| Arterial thrombotic event | 15612430 | 5297230 | 64880 | 415.5663 | East of England |
| Ischaemic stroke | 15612430 | 5297230 | 24840 | 159.09118 | East of England |
| Venous thrombotic event | 15612430 | 5297230 | 21955 | 140.61872 | East of England |
| Acute myocardial infarction | 13802280 | 4687645 | 31500 | 228.208684 | Yorkshire and The Humber |
| Arterial thrombotic event | 13802280 | 4687645 | 66345 | 480.674231 | Yorkshire and The Humber |
| Ischaemic stroke | 13802280 | 4687645 | 25070 | 181.643911 | Yorkshire and The Humber |
| Venous thrombotic event | 13802280 | 4687645 | 21915 | 158.770878 | Yorkshire and The Humber |
| Acute myocardial infarction | 72835475 | 24687730 | 92275 | 126.686895 | Female |
| Arterial thrombotic event | 72835475 | 24687730 | 240870 | 330.701495 | Female |
| Ischaemic stroke | 72835475 | 24687730 | 108965 | 149.605672 | Female |
| Venous thrombotic event | 72835475 | 24687730 | 110870 | 152.221156 | Female |
| Acute myocardial infarction | 71923260 | 24392845 | 183120 | 254.601926 | Male |
| Arterial thrombotic event | 71923260 | 24392845 | 363790 | 505.801615 | Male |
| Ischaemic stroke | 71923260 | 24392845 | 122515 | 170.344063 | Male |
| Venous thrombotic event | 71923260 | 24392845 | 104215 | 144.900277 | Male |
| *Person years, number of people, number of events are rounded to fives to comply with privacy regulations. | | | | | |

#### **Table S9. Breakdown of arterial and venous thrombotic event composite cases datasets.**

| **Composite Outcome** | **Component Outcome** | **Count** | **% of Composite*** |
| --- | --- | --- | --- |
| **Arterial thrombotic event** | **Total individuals** | 546,185 | 100% |
|  | Myocardial infarction | 260,915 | 47.77% |
|  | Ischaemic stroke | 208,965 | 38.26% |
|  | Stroke not otherwise specified | 42,760 | 7.82% |
|  | Retinal infarction | 1,625 | 0.30% |
|  | Other arterial thrombosis | 19,420 | 3.56% |
|  | Arterial dissection/ruptured aneurysm | 42,095 | 7.71% |
| **Venous thrombotic event** | **Total individuals** | 202,790 | 100% |
|  | Venous thrombosis | 92,955 | 45.84% |
|  | Pulmonary embolism | 110,975 | 54.72% |
|  | Intracranial venous thrombosis | 2,505 | 1.24% |
| * Components do not add up to 100%, as some individuals had multiple types of events. | | | |

#### **Table S10. Daily mean air temperature and PM2.5 distributions in England and regions, 2020-2021 and 2020-2022.**

See excel file "S9_tmean_pm2p5_distribution.xlsx"

#### **Table S11. Model coefficients, number of individuals, number of events.**

See excel file "S10_first_stage_models.xlsx"

#### **Table S12. Venous thrombotic event cumulative incidence rate ratio per temperature percentile compared to the median, over 0-21 day lag period. Overall column for the main analysis, and additional columns for the subgroups.**

See excel file "cumulative_IRR_by_percentile_ve.xlsx"

#### **Table S13. Arterial thrombotic event cumulative incidence rate ratio per temperature percentile compared to the median, over 0-21 day lag period. Overall column for the main analysis, and additional columns for the subgroups.**

See excel file "cumulative_IRR_by_percentile_ae.xlsx"

#### **Table S14. Acute myocardial infarction cumulative incidence rate ratio per temperature percentile compared to the median, over 0-21 day lag period. Overall column for the main analysis, and additional columns for the subgroups.**

See excel file "cumulative_IRR_by_percentile_mi.xlsx"

#### **Table S15. Ischaemic stroke cumulative incidence rate ratio per temperature percentile compared to the median, over 0-21 day lag period. Overall column for the main analysis, and additional columns for the subgroups.**

See excel file "cumulative_IRR_by_percentile_stroke.xlsx"

#### **Table S16. Excess events, attributable incidence rate, and attributable fraction per outcome and subgroup.**

See excel file "S15_impact_analysis.xlsx"

#### **Table S17. Venous thrombotic event cumulative incidence rate ratio per temperature percentile compared to the median, over 0-21 day lag period. Overall column for the main analysis, and additional columns for the sensitivity analyses.**

See excel file "cumulative_IRR_by_percentile_sens_ve.xlsx"

#### **Table S18. Arterial thrombotic event cumulative incidence rate ratio per temperature percentile compared to the median, over 0-21 day lag period. Overall column for the main analysis, and additional columns for the sensitivity analyses.**

See excel file "cumulative_IRR_by_percentile_sens_ae.xlsx"

#### **Table S19. Acute myocardial infarction cumulative incidence rate ratio per temperature percentile compared to the median, over 0-21 day lag period. Overall column for the main analysis, and additional columns for the sensitivity analyses.**

See excel file "cumulative_IRR_by_percentile_sens_mi.xlsx"

#### **Table S20. Ischaemic stroke cumulative incidence rate ratio per temperature percentile compared to the median, over 0-21 day lag period. Overall column for the main analysis, and additional columns for the sensitivity analyses.**

See excel file "cumulative_IRR_by_percentile_sens_stroke.xlsx"

### **References**

1.	Knight R, Walker V, Ip S, et al. Association of COVID-19 With Major Arterial and Venous Thrombotic Diseases: A Population-Wide Cohort Study of 48 Million Adults in England and Wales. *Circulation* 2022; 146(12): 892-906.

2.	Institute for Government. Timeline of UK government coronavirus lockdowns and restrictions. INSTITUTE FOR GOVERNMENT, 2022.

3.	Baker RE, Mahmud AS, Miller IF, et al. Infectious disease in an era of global change. *Nature Reviews Microbiology* 2022; 20(4): 193-205.

4.	NOAA National Centers for Environmental Information. Monthly Global Climate Report for Annual 2023. 2024. <https://www.ncei.noaa.gov/access/monitoring/monthly-report/global/202313>.

5.	Romanello M, Napoli Cd, Green C, et al. The 2023 report of the <em>Lancet</em> Countdown on health and climate change: the imperative for a health-centred response in a world facing irreversible harms. *The Lancet* 2023; 402(10419): 2346-94.

6.	Vicedo-Cabrera AM, Scovronick N, Sera F, et al. The burden of heat-related mortality attributable to recent human-induced climate change. *Nat Clim Chang* 2021; 11(6): 492-500.

7.	van Oldenborgh GJ, Mitchell-Larson E, Vecchi GA, de Vries H, Vautard R, Otto F. Cold waves are getting milder in the northern midlatitudes. *Environmental Research Letters* 2019; 14(11): 114004.

8.	Lo YTE, Mitchell DM, Watson PAG, Screen JA. Changes in Winter Temperature Extremes From Future Arctic Sea-Ice Loss and Ocean Warming. *Geophysical Research Letters* 2023; 50(3): e2022GL102542.

9.	NASA. Shifting Distribution of Land Temperature Anomalies, 1951-2020. https://svs.gsfc.nasa.gov/48912023).

10.	Alahmad B, Khraishah H, Royé D, et al. Associations Between Extreme Temperatures and	Cardiovascular Cause-Specific Mortality: Results From 27 Countries. *Circulation* 2023; 147(1): 35-46.

11.	Al-Kindi SG, Brook RD, Biswal S, Rajagopalan S. Environmental determinants of cardiovascular disease: lessons learned from air pollution. *Nature Reviews Cardiology* 2020; 17(10): 656-72.

12.	Raisi-Estabragh Z, Cooper J, Salih A, et al. Cardiovascular disease and mortality sequelae of COVID-19 in the UK Biobank. *Heart* 2023; 109(2): 119.

13.	Wood A, Denholm R, Hollings S, et al. Linked electronic health records for research on a nationwide cohort of more than 54 million people in England: data resource. *BMJ* 2021; 373: n826.

14.	Vanderweele TJ. Invited commentary: assessing mechanistic interaction between coinfecting pathogens for diarrheal disease. *Am J Epidemiol* 2012; 176(5): 396-9.

15.	Wang J, Tang K, Feng K, et al. Impact of temperature and relative humidity on the transmission of COVID-19: a modelling study in China and the United States. *BMJ Open* 2021; 11(2): e043863.

16.	Kang D, Ellgen C, Kulstad E. Possible effects of air temperature on COVID-19 disease severity and transmission rates. *J Med Virol* 2021; 93(9): 5358-66.

17.	Christophi CA, Sotos-Prieto M, Lan F-Y, et al. Ambient temperature and subsequent COVID-19 mortality in the OECD countries and individual United States. *Scientific Reports* 2021; 11(1): 8710.

18.	Quilodrán CS, Currat M, Montoya-Burgos JI. Air temperature influences early Covid-19 outbreak as indicated by worldwide mortality. *Science of The Total Environment* 2021; 792: 148312.

19.	Usmani M, Jamal Y, Gangwar M, et al. Asymmetric Relationship between Ambient Air Temperature and Incidence of COVID-19 in the Human Population. *Am J Trop Med Hyg* 2022; 106(3): 877-85.

20.	Hernandez Carballo I, Bakola M, Stuckler D. The impact of air pollution on COVID-19 incidence, severity, and mortality: A systematic review of studies in Europe and North America. *Environ Res* 2022; 215(Pt 1): 114155.

21.	Lolli S, Chen Y-C, Wang S-H, Vivone G. Impact of meteorological conditions and air pollution on COVID-19 pandemic transmission in Italy. *Scientific Reports* 2020; 10(1): 16213.

22.	Sun Z, Chen C, Xu D, Li T. Effects of ambient temperature on myocardial infarction: A systematic review and meta-analysis. *Environmental Pollution* 2018; 241: 1106-14.

23.	Mustafić H, Jabre P, Caussin C, et al. Main Air Pollutants and Myocardial Infarction: A Systematic Review and Meta-analysis. *JAMA* 2012; 307(7): 713-21.

24.	Johansen NR, Kjaerbye-Thygesen A, Jonsson S, Westh H, Nilas L, Rorbye C. Prevalence and treatment of group B streptococcus colonization based on risk factors versus intrapartum culture screening. *European journal of obstetrics, gynecology, and reproductive biology* 2019; 240: 178-81.

25.	Gasparrini A. The Case Time Series Design. *Epidemiology* 2021; 32(6): 829-37.

26.	Huang J, Wang J, Yu W. The Lag Effects and Vulnerabilities of Temperature Effects on Cardiovascular Disease Mortality in a Subtropical Climate Zone in China. *International Journal of Environmental Research and Public Health* 2014; 11(4): 3982-94.

27.	Dahlquist M, Frykman V, Hollenberg J, et al. Short‐Term Ambient Air Pollution Exposure and Risk of Out‐of‐Hospital Cardiac Arrest in Sweden: A Nationwide Case‐Crossover Study. *Journal of the American Heart Association* 2023; 12(21): e030456.

1. Approvals are being extended to the end of 2024, although we envisage extending beyond this date in the future.
2. Word counts are only a guide and can be exceeded, if necessary.
3. Your plain English summary will appear on the [CVD-COVID-UK / COVID-IMPACT webpage](https://bhfdatasciencecentre.org/areas/cvd-covid-uk-covid-impact/). Please use the sort of language you might use to describe your project to a non-specialist friend, relative or journalist. Please avoid using technical jargon and aim to keep sentences short for ease and clarity of reading.
4. **COVID-IMPACT:** COVID-related research projects **not** directly linked to cardiovascular disease and/or its risk factors.
5. Includes patients with active, current registrations at participating practices and deceased patients with a date of death on or after 1 November 2019. Note: prescriptions and numeric values (e.g. BP, laboratory test results) only go back two years.
6. Pillar 1 and 2 positive tests
7. Data provided comprises a single cut of the data as at June 2020 with no current updates. Based on data used in the EAVEII project.
8. Contains the first positive test result per person or earliest test result if they have never tested positive (dataset not updated after August 2021 – replaced by Covid Tests)
9. Contains all test results (positive and negative) and replaced the ECOSS dataset from August 2021.
10. Additional approval process required for this dataset.
11. Additional approval process required for this dataset.
12. Contains data to identify patients receiving hospital based renal replacement therapy – haemodialysis – only (from January 2019).
13. Additional approval process required for this dataset
14. Additional approval process required for this dataset (4-6 week lead time). Analysts requiring access must be ONS Safe Researcher Training certified and have a valid Accredited Researcher (AR) number.
15. Additional approval process required for this dataset (4-6 week lead time). Analysts requiring access must be ONS Safe Researcher Training certified and have a valid Accredited Researcher (AR) number.
